# Single-cell gene programs define subtype identity and metastatic trajectories in renal cell carcinoma

**DOI:** 10.64898/2026.07.14.26357682

**Authors:** Ariel Madrigal, Minjun Kim, Zohreh Mehrjoo, Tamiko Nishimura, Ozge Saatci, Adrien Osakwe, Evelyn Zavacky, Elham Moslemi, Kate I Glennon, Matthew Dankner, Sarah M. Maritan, Hellen Kuasne, Virginie Pilon, Anie Monast, Mustafa Soytas, Madeleine Arseneault, Spyridon Oikonomopoulos, Ashot Harutyunyan, Tianyuan Lu, Roni Rayes, Larisa M. Soto, Aldo Hernandez-Corchado, Jonathan D. Spicer, Kevin Petrecca, Peter Siegel, Morag Park, Jiannis Ragoussis, Ozgur Sahin, Fadi Brimo, Simon Tanguay, Yasser Riazalhosseini, Hamed S. Najafabadi

## Abstract

While extensive cellular heterogeneity in renal cell carcinomas (RCC) is linked to diverse clinical outcomes, our understanding of this diversity is limited to those driven by clonal patterns or activity of canonical pathways. Here, we present a compendium of over 85,000 single-cell gene expression profiles from primary and metastatic tumors as well as patient-derived models across four RCC subtypes, including the rare clear cell papillary renal cell tumors, which we show are often misclassified and for which we identify CASP14 as a highly sensitive and specific biomarker. We dissect malignant cell variation within and across tumors using a generative modeling framework that accounts for clonal and copy number-driven expression shifts, defining 59 gene expression programs that deconstruct canonical pathways into functional submodules with divergent activity patterns, distinct regulators, and differential association with clinical outcomes. Despite the canonical view that VHL-deficient clear cell RCC exists in a constitutive pseudohypoxic state, we show strong intra-tumor variability of a hypoxia inducible factor 2 (HIF2)-driven program linked to poor outcome. We also identify early, spatially organized activation of a complete epithelial-to-mesenchymal transition (EMT) program, loss of epithelial identity, and upregulation of protein translation programs as key characteristics of metastatic progression. Finally, a metastatic signature capturing cellular de-differentiation and translational activity identifies primary tumors associated with adverse clinical outcomes. Together, this resource establishes a framework for dissecting malignant cell heterogeneity, refines RCC subtype classification, and defines transcriptional programs underlying metastasis progression.

## Introduction

Renal cell carcinoma (RCC) comprises a heterogeneous group of malignancies originating from renal tubular epithelial cells and includes multiple histological and molecular subtypes^1^. Clear cell RCC (ccRCC) is the most common, representing about 80% of cases, followed by papillary RCC (pRCC) and chromophobe RCC (chRCC)^2^. These subtypes differ in their cells of origin: ccRCC and pRCC arise from the proximal tubule, whereas chRCC originates from the distal tubule^3^. These origins contribute to differences in immunohistochemical and molecular profiles and potentially to clinical outcomes^2^, making accurate subtype classification essential for prognosis and treatment selection. However, with the recent and continuing expansion in renal tumor subtype classifications, determining the accurate diagnosis can be challenging^4,5^, particularly in rare tumors with mixed and overlapping histological features such as clear cell papillary renal cell tumors (ccpRCT).

In addition to differences between subtypes, RCC exhibits extensive intra-tumor heterogeneity. Subclonal genetic evolution is an important contributor to this variability, as demonstrated by multi-region sequencing studies^6,7^. However, intra-tumor heterogeneity also arises from non-genetic mechanisms occurring in both malignant and non-malignant cell populations^8^, which may include transcriptional plasticity, epigenetic regulation and tumor microenvironment (TME) effects. Understanding this variability is important, as it can generate diverse cellular states that enable rapid adaptation to perturbations and promote therapy resistance^8,9^.

The advent of single-cell technologies has enabled a much deeper characterization of intra-tumor heterogeneity. Given the immunogenicity of ccRCC, substantial effort has focused on dissecting heterogeneity within the immune compartment. These efforts include profiling of immune cell landscapes and their association with survival outcomes^10^, characterization of the transcriptional changes across disease stages^11^, and the identification of immune cell states associated with responses to immunotherapy^12,13^.

Despite these advancements, malignant cell heterogeneity (MCH) in RCC remains comparatively under-characterized by single-cell studies, in part due to substantial analytical challenges. A major obstacle is the strong patient-specific signal in malignant cells^14^, which complicates the application of traditional integration methods. To address this challenge, recent efforts have begun to examine malignant cell diversity through the identification of gene programs^15–17^. Although informative, these strategies have several drawbacks: they often overlook the impact of intra-clonal heterogeneity on shifting gene-expression baselines; they typically reduce heterogeneity to a limited number of meta-programs that may aggregate biologically distinct processes; and they are generally restricted to a single RCC subtype, most commonly ccRCC.

Another aspect that remains poorly understood is how the spectrum of MCH evolves during metastatic progression. Metastasis is the leading cause of death in kidney cancer, reducing the 5-year relative survival rate from 93% in localized disease to 19% in patients with distant metastases^18^. Characterization of gene expression differences between primary tumors and metastatic lesions, particularly in lymph nodes and bone^19–21^, have highlighted broad shifts in malignant cell states, including increased activation of epithelial-to-mesenchymal (EMT) processes^19^. However, substantial gaps remain, including a more comprehensive description of the transcriptional programs that emerge early in primary tumors and are associated with invasive potential, as well as those that arise in established distant metastases.

Here, we present a comprehensive cellular atlas of primary and metastatic RCCs spanning four distinct subtypes. Through the integration of single-cell transcriptomic profiles, spatial RNA maps, and large-scale bulk RNA-seq datasets, we (i) refine the characterization of histological subtypes, including resolving cases suggestive of misclassification in rare tumors, and identify a novel biomarker for the ccpRCT subtype; (ii) provide a panoramic view of MCH by identifying gene programs that reveal functional submodules within canonical pathways; and (iii) delineate gene programs, associated with early transient states, including invasiveness, as well as early-stable and late cellular changes during metastasis.

## Results

### A cellular atlas of primary and metastatic RCCs

We assembled a diverse cohort of RCC samples, encompassing 21 specimens spanning four RCC subtypes, including 16 ccRCC, 2 pRCC, and 1 chRCC samples, in addition to one sample from the rarer ccpRCT subtype and one not-otherwise-specified (NOS) RCC tumor. The cohort comprises 12 primary and 9 metastatic tumor samples, including 16 patient tumors and 5 patient-derived xenograft (PDX) models derived from primary tumors or metastases in the brain and lung (**Figure 1a** and **Supplementary Data Table 1**; primary and metastatic tumors are not patient-matched). Using the droplet-based 10x Genomics Chromium platform (single-cell 3′), we generated a cellular map of transcriptional heterogeneity in RCC, comprising 85,563 single-cell expression profiles passing QC criteria (see **Methods** for details). This dataset captures diverse cell populations within the epithelial, immune, and stromal compartments, each characterized by distinct gene markers (**Figure 1b-c**; see Methods for details on cell type identification and annotation). We observed marked heterogeneity in cellular composition across tumors, with some samples enriched for malignant epithelial cells, while others were dominated by immune cell populations (**Supplementary Figure 1**). Overall, we captured 45,494 malignant cells, including 16,212 primary and 29,282 metastatic cancer cells.

**Figure 1.**
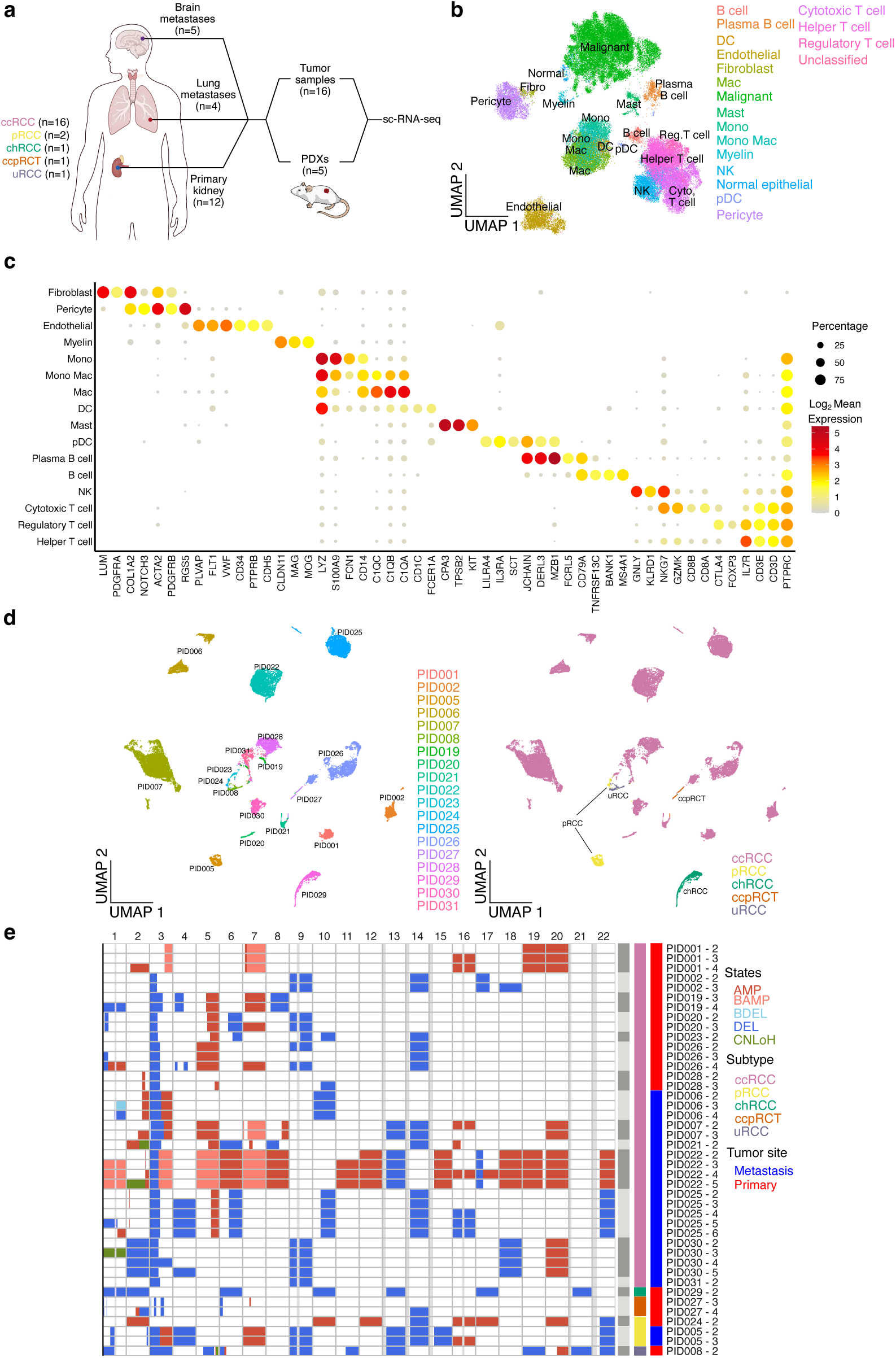
Overview of the RCC single-cell compendium. (**a**) Schematic of the experimental design for single-cell transcriptomic profiling of renal cell carcinoma samples from primary and metastatic tumors. (**b**) UMAP embedding of RCC patient tumor cells after integration with GEDI^34^ (see Methods). Each dot represents a single cell, colored by cell type labels. (**c**) Dot plot of representative marker genes for each cell type. Dot color reflects log_2_ mean expression, and dot size indicates the percentage of cells expressing a given gene. Only gene-cell type pairs where at least 5% of cells within that cell type express the gene are shown. (**d**) UMAP embedding of malignant RCC cells from tumor samples and PDXs. For generating this embedding, no integration was enforced. Only samples with more than 100 cancer cells, resulting in analysis of 44,739 malignant cancer cells across 19 samples were included in this analysis. Labels indicate the patient donor (left) or the subtype of each sample (right). (**e**) CNV analysis across malignant sub-clones. Each heatmap row represents a subclone, with colors denoting CNV states identified by Numbat^80^: amplification, deletion, balanced amplification, balanced deletion and copy neutral loss of heterozygosity. Gray vertical bars indicate gap regions.

Consistent with previous observations^14^, transcriptomic analysis of malignant RCC cells exhibited pronounced inter-tumor heterogeneity, evidenced by sample-specific clustering (**Figure 1d**). Among the contributing factors, patient-specific copy number variations (CNVs) play a significant role in driving these inter-tumor heterogeneity effects, evident from expression-based inference of single-cell CNV profiles (**Figure 1e**). The inferred CNVs also revealed recurrent alterations within each subtype. Loss of the short arm of chromosome 3 (chr3p) was observed in 13 out of 14 ccRCC samples, with the remaining tumor harboring a *VHL* mutation (**Supplementary Data Table 1**) and overall downregulation of genes located on chr3p (**Supplementary Figure 2**). Additional CNV hallmarks^22^ observed in the ccRCC samples included loss of the long arm of chromosome 14, which was present in 6 samples, and gain of the long arm of chromosome 5, observed in 8 samples. The chRCC sample exhibited deletions of chromosomes 1,6,10,13,17 and 21, which are hallmarks of this subtype^1^. Among the pRCCs, one showed amplification of chromosome 7, while the other displayed gain of chromosome 17, both characteristic rearrangements^22^. Bulk genomic analyses, either through whole-exome or an RCC-specific gene panel^23^ sequencing of samples confirmed the presence of characteristic gene mutations, and broadly confirmed these single-cell-based CNV profiles (**Supplementary Figure 3, Supplementary Data Table 1**).

### The transcriptomic profiles of RCCs reveal misclassification of uncommon subtypes

Subtype gene expression signatures are an important tool for classification and characterization of RCCs^24^. Signatures obtained from bulk-tissue data, however, can be contaminated by genes expressed in tumor-infiltrating cells^25^. We used our single-cell compendium to create cancer cell-intrinsic gene signatures for ccRCC, chRCC, pRCC, and ccpRCT subtypes, free of confounding effects from stromal and immune contributions (see **Methods** for details). When applied to three RCC cohorts from The Cancer Genome Atlas (TCGA) project, including the KIRC (ccRCC), KICH (chRCC), and KIRP (pRCC) cohorts^26,27^, our gene signatures showed overall high agreement with the recorded sample annotations (**Supplementary Figure 4a-c**). Nonetheless, we also observed discrepancies, such as samples from KICH and KIRP cohorts obtaining high ccRCC signature scores, or samples with high chRCC signature scores in the KIRC and KIRP cohorts. Most surprisingly, we identified several samples with unexpectedly high ccpRCT signature scores, even though no ccpRCTs have been reported in the TCGA cohorts (**Supplementary Figure 4d**).

These observations prompted us to examine the possibility of clinical misclassification of RCCs and the potential of transcriptomic signatures to better resolve RCC subtypes. We started with an unbiased clustering of 1,017 TCGA samples based on their transcriptomic profiles, limiting our analysis to genes that, based on our single-cell data, were not markers of any tumor-infiltrating cells, to limit the potential confounding effect of tumor impurity on clustering (see **Methods**). We used shared nearest neighbour clustering^28^ to identify 23 transcriptomic clusters, which were further merged into six major groups by hierarchical clustering (**Figure 2a** and **Supplementary Data Table 2**). Although these groups largely aligned with TCGA-reported diagnosis (**Figure 2b**), we also observed numerous inconsistencies. For example, 19 out of 65 samples from the TCGA KIRC cohort were grouped together with samples from KICH (chRCC) cohort based on their RNA profiles. At the protein level, these samples also showed high expression of c-KIT (**Figure 2c**), which is a biomarker of chRCC, consistent with the transcriptomic-based clustering. Conversely, six samples from the KICH cohort clustered together with those of other subtypes—these samples did not show c-KIT expression at the protein level (**Figure 2d**), again consistent with our transcriptomic-based clustering rather than the original annotation.

**Figure 2.**
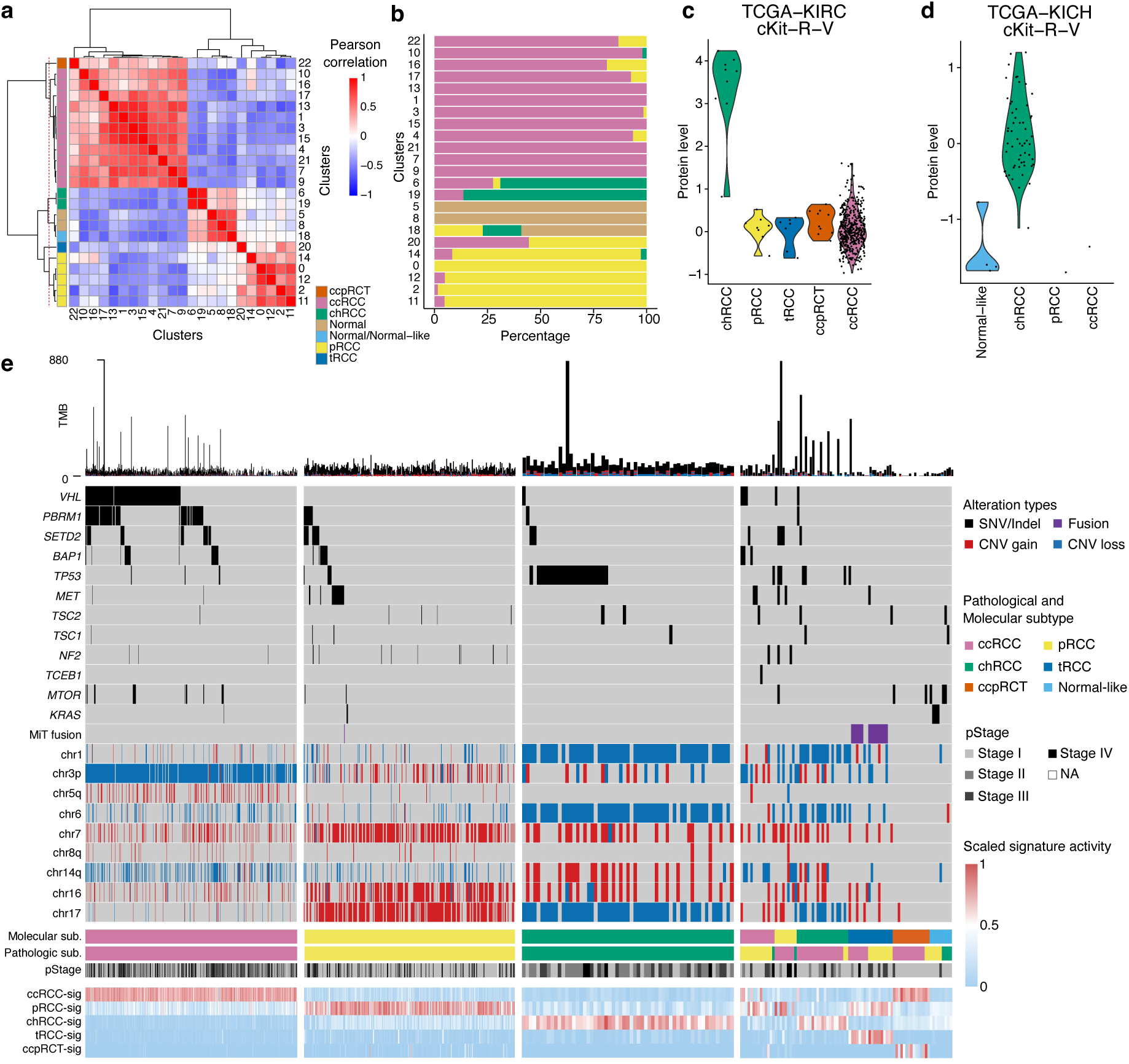
Misclassification of RCCs in TCGA. (**a**) Transcriptomic reclassification of RCC tumors using bulk-RNA-seq profiles from TCGA. Combining transcriptomic profiles of KIRC, KICH, KIRP and normal kidney tissue samples, 23 clusters were derived using shared-nearest neighbor clustering (see Methods). Heatmap shows Pearson correlation between centered mean expression profiles of the 23 clusters. Left bar plot indicates the six major RCC groups identified via hierarchical clustering that define our molecular subtype. (**b**) Comparison between original diagnosis and molecular subtype of RCC tumors. Bar plot shows the percentage distribution of original diagnoses within each transcriptomic cluster. (**c**) Violin plot showing the distribution of c-KIT protein levels in TCGA KIRC samples, grouped by molecular subtypes defined by transcriptomic clustering. (**d**) Same as (c) but showing the distribution of c-KIT in TCGA KICH samples. (**e**) Oncoplot of RCC tumors in TCGA showing selected molecular alterations, original diagnosis, molecular subtype and subtype-specific transcriptomic signatures. Signatures are normalized using min-max normalization. Top bar indicates tumor mutation burden.

We further investigated genomic landscapes and potential linkages between known subtype-specific genomic alterations and RCC subtypes assigned by transcriptome profiles using TCGA data available for the same cohorts described above. These analyses substantiated our preceding observations by revealing that many tumors with discordant histological and transcriptomic diagnoses exhibit CNVs and/or somatic mutations that are consistent with transcriptome-based diagnosis (**Figure 2e**). For example, among the TCGA-KIRC cohort, genomic hallmarks of ccRCC such as *VHL* mutation or loss of chr3p are significantly more frequent in tumors classified as ccRCC by transcriptomic clustering than in those that are not (94.3% vs 18.8%; 459 of 487 vs 9 of 48, Fisher’s exact test *p* = 2 ×10^−32^). Similarly, in both TCGA-KIRP and KICH cohorts, canonical CNV hallmarks are significantly enriched in tumors clustered as consensus subtypes. For the KIRP cohort, simultaneous gain of chromosomes 7 and 17 is observed in 58% in pRCC clusters, but only 9.4% in those clustered as non-pRCC (149 of 257 vs 3 of 32, p = 8.1 x 10^-8^). Concomitant loss of chromosomes 1, 6, and 17 is only found in chRCC clusters for the KICH cohort (79.7% vs 0%; 47 of 59 vs 0 of 6, p = 2.2×10^−4^) ^22^.

In addition to sporadic reassignment of samples by transcriptome signatures, we also observed clusters that were highly distinct from those of major RCC subtypes. For example, 18 samples from the TCGA KIRC and KICH cohorts grouped together into one cluster, with a transcriptomic profile that showed little similarity to other clusters containing KIRC or KICH samples (**Figure 2a**, cluster 20). For a large majority of the samples from this cluster, we were able to detect, based on RNA-seq reads, the fusion of *TFE3* gene with other genes, with *SFPQ* being the most prevalent fusion partner (**Supplementary Figure 5**), suggesting that the samples of this cluster in fact correspond to MiT-family translocation RCC (tRCC)^29^. To further verify the tRCC identity of these samples, we used external data^30–33^ to build a tRCC gene signature and applied it to the TCGA data (see **Methods**); indeed, we observed high tRCC signature scores for this cluster (**Figure 2e**). Among the 18 tumors in the tRCC cluster, the majority (n=13) show both the MiT/TFE fusions and tRCC-specific signature. However, we observed that the remaining 5 tumors only show high expression of the signature without previously identified MiT/TFE gene fusions^30^. This observation is consistent with a previous report on tRCCs cases showing TFE3-positivity in IHC staining without fusion detection^32^, which could be due to the technical shortcomings of short-read sequencing for fusion detection. As another example, we identified a transcriptomic cluster that, in addition to several normal kidney samples, included tumors from the KICH (chRCC) and KIRP (pRCC) cohorts (**Figure 2a**), suggesting the presence of a distinct group of RCCs. Notably, samples of this group did not show CNVs or mutations of common RCC subtypes, but rather harbored mutations in *KRAS* (3/9, 33%), *MTOR* (3/9, 33%), *TSC1* (1/9, 11%) and *TSC2* (1/9, 11%), in a mutually exclusive manner (**Figure 2e**).

### CASP14 expression is a marker of ccpRCT

During our analyses, we observed that one of the transcriptionally distinct clusters (**Figure 2a**, cluster 22) encompassed the majority of samples with high ccpRCT signature scores (**Figure 3a**). Direct comparison of the transcriptomic profiles of the samples in this cluster and the single-cell profiles of our ccpRCT sample revealed that they are reciprocal best matches. Specifically, we integrated^34^ TCGA RNA-seq profiles with our single-cell data and measured their similarity in the resulting harmonized low-dimensional space. This revealed that the centroid of cluster 22 had the highest cosine similarity to our ccpRCT single cells (**Figure 3b**), and the centroid of our ccpRCT single cells had the highest cosine similarity with the cluster 22 samples (**Figure 3c**). Finally, to confirm that the samples from cluster 22 are ccpRCTs, we obtained images of H&E staining of these tumors from cBioPortal and had their morphological characteristics re-examined by a dedicated genitourinary pathologist (FB). Indeed, in 15 out of 15 samples, we were able to detect morphological features of ccpRCTs, with tubulopapillary architecture, bland-looking clear cells with apical nuclear arrangements (**Figure 3d**), supporting the transcriptomic-based classification. Notably, these tumors were devoid of common RCC-associated CNVs or mutations (**Figure 2e**), suggesting that their molecular footprints may be different from those of common RCC subtypes.

**Figure 3.**
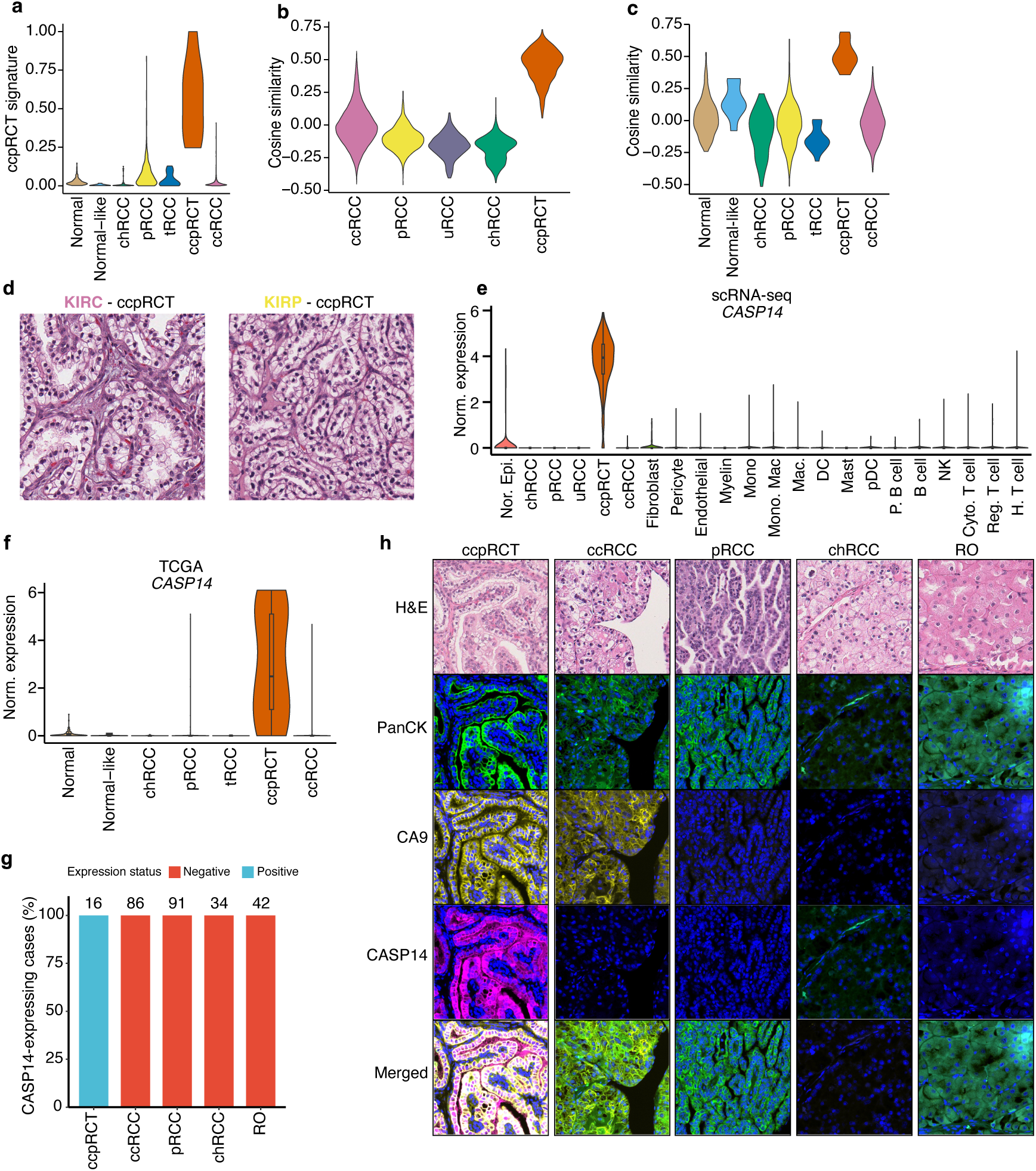
Identification of CASP14 as a molecular marker of ccpRCT. (**a**) Violin plot showing the distribution of ccpRCT signature activity across RCC tumors and normal kidney samples from TCGA. The x-axis indicates molecular subtypes. (**b**) Comparison of overall transcriptomic similarity between the ccpRCT cluster in TCGA and single-cell expression profiles of malignant cells across RCC subtypes. To obtain this plot, gene expression profiles from TCGA RCC samples and single-cell malignant RCC cells were integrated into a shared latent space using GEDI^34^. Then, a mean embedding vector of the ccpRCT cluster was computed and compared to single-cell embeddings using cosine similarity. (**c**) Comparison of overall transcriptomic similarity between malignant ccpRCT cells and molecular subtypes from TCGA. Cosine similarity was computed as in (b) but starting from the mean embedding vector of ccpRCT malignant cells. (**d**) Representative hematoxylin and eosin (H&E) staining of two TCGA tumors reclassified as ccpRCT based on molecular profiling. (**e**) Single-cell mRNA expression of *CASP14* in the tumor microenvironment. (**f**) mRNA expression of *CASP14* in RCC tumors from TCGA. (**g**) CASP14 expression determined by IF staining in a cohort of RCC tissue microarrays. Bar plot shows the percentage of samples with positive or negative CASP14 expression. (**h**) Examples of IF staining for selected markers, including CASP14, in tissue microarrays of various RCC subtypes.

Together, these results suggest that recently recognized and less common forms of renal tumors were previously misclassified by histopathological examinations, including in large well-characterized reference public tumor libraries: our analysis reclassified 9.7% of TCGA samples (86/889), with ∼17% (15/86) of these cases corresponding to ccpRCT samples that were originally misclassified as ccRCC (13 cases) or, less frequently, as pRCCs (2 cases) (**Figure 2b**). While ccpRCTs are currently known to show a different immunoprofile pattern than ccRCC (using CA9, GATA3, CK34BE12, CD10), which aid in their differential diagnosis, significant morphological overlap still exists, especially in biopsy specimens, as many ccRCCs were shown to harbor ccpRCT-like area morphologically^35^. In addition, no single ccpRCT-specific reliable immunomarker is described to date, in parallel to the lack of characteristic genetic alterations as seen in other tumor subtypes^36,37^. To explore the possibility of finding immunomarkers for ccpRCT, we used our single-cell data, in combination with the bulk expression profiles of reclassified ccpRCTs from TCGA, to identify genes that (i) are highly expressed in ccpRCT cancers cells, (ii) are not expressed in cancer cells of other subtypes, and (iii) are not expressed in tumor-infiltrating cells in any subtype. We identified three potential candidates (**Figure 3e,f** and **Supplementary Figure 6**), of which we were able to establish an antibody-based assay for the CASP14 protein. To evaluate the potential value of CASP14 expression as a marker of ccpRCT, we first examined its expression using immunostaining in a cohort of 16 well-characterized ccpRCT cases diagnosed at the McGill University Health Centre. All these tumors showed high cytoplasmic CASP14 expression in malignant cells (**Figure 3g**). Next, to validate the specificity of CASP14 as a ccpRCT marker, we applied the CASP14 immunostaining to an independent cohort of 255 RCC tumors and 27 normal kidney samples, including 86 ccRCC, 91 pRCC, 34 chRCC, 42 renal oncocytomas (RO), and two other cases (1 NOS tumor and 1 renal carcinoid). Immunofluorescence (IF) analysis using five tissue microarrays (TMAs) in this cohort confirmed lack of CASP14 expression in RCC subtypes other than ccpRCT (**Figure 3h**), allowing us to distinguish ccpRCTs from other RCCs with a sensitivity of 1.00 (16 out of 16 ccpRCTs; 95% confidence interval 0.81-1.00, Wilson method) and specificity of 1.00 (0 out of 282 non-ccpRCTs including 27 normal kidney samples; 95% confidence interval 0.99-1.00, Wilson method).

### Intra-tumor gene expression programs reveal clinically relevant functional submodules in RCCs

Having refined RCC subtype classification through molecular profiling, we sought to examine the cellular states that are common among and/or specific to RCC subtypes, by systematic analysis of single-cell gene programs (GP)—sets of co-expressed genes under shared regulatory influences with similar functional states^38^. A key challenge in identifying GPs within tumors is that, in addition to technical noise and unwanted sources of variation, gene program discovery can be confounded by genomic alterations (such as copy number loss or amplification). Such rearrangements can shift baseline gene expression in a sample- or subclone-specific manner, obscuring true patterns of coordinated expression (schematically shown in **Figure 4a**). This effect is particularly pronounced in our dataset, as we observed a total of 42 malignant sub-clones identified across the 21 RCC samples (**Figure 1e**)—in several samples, inter-clonal gene expression differences dominated the transcriptomic heterogeneity of the cells (**Figure 4b**). To define GPs in our RCC single-cell dataset, we devised a novel generative modeling framework, based on convex non-negative matrix factorization^39^, to identify sets of genes that covary across cells, while taking into account the statistical characteristics of single-cell data such as Poisson noise and overdispersion of counts (**Supplementary Figure 8**; see **Methods** for details). By explicitly modeling the clonal shifts in gene expression, we focus on GPs that contribute to intra-tumor (and intra-clonal) heterogeneity, without the confounding effect of CNVs and other clone-or sample-level sources of variation. Also, by expressing each gene program as a convex combination of multiple genes, our framework intrinsically provides a measure of gene program activity in each individual cell (**Figure 4c**).

**Figure 4.**
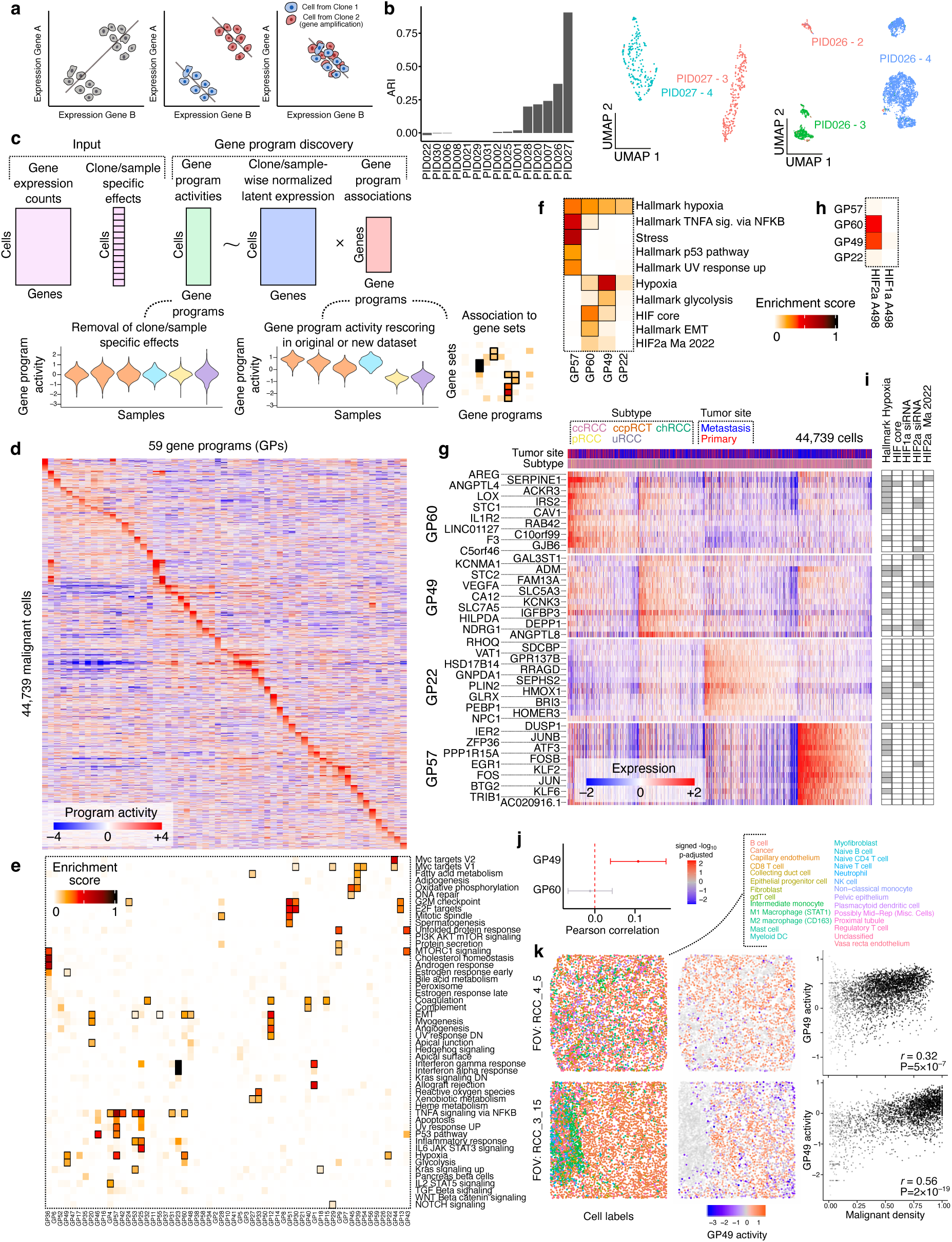
Gene expression programs driving intra-tumor heterogeneity in malignant RCC cells. (**a**) Schematic illustrating how genomic alterations can distort apparent co-expression relationships in a sample-or clone-specific manner. The diagram shows how a gene amplification event in a particular clone shifts baseline expression, and how accounting for these changes enables inference of correct co-expression relationships. (**b**) Bar plot shows Adjusted Rand Index scores comparing intra-tumor transcriptomic cluster assignments and clones identified by Numbat^80^ (left). Examples of intra-tumor clustering and clonal diversity for two samples (middle and right). Each plot displays a UMAP embedding computed from the log-normalized expression counts for that sample, with colors indicating clonal identity. Additional examples can be found in Supplementary Figure 7. (**c**) Schematic of the scConvexNMF model. The inferred gene programs are constructed as weighted combinations of gene-expression profiles and provide a measure of program activity for each individual cell. During the program discovery step, sample- or clone-specific are removed. Once the programs are inferred, they can be applied to compute activity scores in either the original or new datasets, while preserving sample or clone-specific effects present in the input data. (**d**) Heatmap of activities for 59 gene programs (columns) across malignant RCC cells (rows). (**e**) Association between gene programs (columns) and Hallmark pathways^40^ (rows). Heatmap shows enrichment scores (see Methods for details). Significant associations (FDR < 0.05) are outlined with a black box. (**f**) Same as in (e) but showing selected gene programs associated with the Hallmark hypoxia pathway. Additional gene sets include: Hypoxia and Stress^46^; HIF core^41^ and HIF2a Ma 2022^42^. (**g**) Intra-tumor heterogeneity of expression for four hypoxia-related gene programs. Heatmap shows the normalized latent-log expression from the scConvexNMF framework. Shown are the top 15 member genes (rows) across individual cells (columns). Top bar indicates metastasis status and RCC subtype for each single cell. (**h**) Association of hypoxia-related programs to gene-sets of HIF1a or HIF2a specific signatures in A498 cells. (**i**) Membership of top genes from hypoxia-related programs as shown in (g) within selected hypoxia gene sets. Grey indicates gene inclusion in the set. (**j**) Weighted correlation analysis between local malignant cell density and activities of GP49 and GP60 in a single-cell spatial transcriptomic dataset^44^. Pearson correlation coefficients and 95% confidence intervals were derived via meta-analysis across 21 FOVs. Color represents signed -log_10_ FDR values. Spatial analysis used a neighborhood-step of *k*=3 (see Supplementary Figure 15 for additional *k* values). (**k**) Example FOVs showing positive association between malignant density and GP49 activity. Left: spatial coordinates of cell centroids, colored by the cell type annotation from the original authors^44^. Middle: spatial coordinates showing GP49 activity in malignant cells; non-malignant cells are transparent. Right: scatterplots comparing GP49 activity and malignant density; dot intensity reflects weights representing local neighborhood support for both signals.

Using this framework, we found that the intra-tumor variability of gene expression in malignant RCC cells can be explained based on the variable activities of 59 GPs (**Figure 4d**, **Supplementary Figure 9** and **Supplementary Data Table 3**). As expected, our framework associates genes to GPs even in the presence of CNVs in a subset of samples and/or clones (**Supplementary Figure 10**). These GPs were overall concordant with known biological pathways; for example, we found 76 significant program-pathway associations (FDR<0.05) between 35 GPs and 38 Hallmark pathways^40^ (**Figure 4e**; see **Supplementary Figure 11** for additional associations based on other pathway annotations). Interestingly, we found 18 instances where the same pathway was associated with more than one GP, effectively deconvolving that pathway into multiple submodules with divergent activity patterns across the cells. These pathways and their submodules are shown in detail in **Supplementary Figure 12**.

A notable example of the multi-module pathways that we found is the hypoxia signaling pathway—pseudohypoxia due to loss of VHL is a hallmark of ccRCCs; however, we found substantial cell-to-cell variability in the expression of Hallmark hypoxia signature^40^ genes even within VHL-deficient ccRCC tumors (**Figure 4f,g**). These genes significantly overlap four distinct GPs: GP60, whose genes are associated with EMT (*SERPINE1*, *AREG*, *LOX*), GP49, which is associated with glycolysis (*GAL3ST1*, *STC2*, *IGFBP3*), GP57, which shows enrichment for stress and TNFa signaling pathways (*DUSP1*, *IER2*, *JUNB*) and contains many immediate-early response genes (e.g., *FOS*, *JUN*, *EGR1*) (**Figure 4g**), and GP22, which does not significantly overlap other Hallmark pathways, but contains several genes involved in oxidative and redox stress response (*HMOX1*, *GLRX*, *SEPHS2*, *HSD17B14*). We refer to these four programs as the hypoxia-EMT program, hypoxia-glycolysis program, immediate-early response (IER) program, and oxidative stress response program, respectively.

Among the four programs, only hypoxia-EMT and hypoxia-glycolysis programs overlapped a previously defined core set of HIF targets^41^ (“HIF core” in **Figure 4f**). The same two gene programs also overlapped a previously defined HIF2a signature^42^ (“HIF2a Ma 2022” in **Figure 4f**), as well as HIF2a ChIP-seq targets^43^ (**Supplementary Figure 13**). These observations suggest that the previously reported hypoxia signature consists of four functionally specialized submodules with different regulatory influences, of which only two are potentially driven by HIFs. To validate this hypothesis, we performed siRNA knockdown experiments targeting HIF1a, HIF2a, or both (in A498 cells) to derive HIF-specific gene signatures (**Supplementary Figure 14**). Comparison of the four gene programs with these HIF-specific signatures revealed that both hypoxia-EMT and hypoxia-glycolysis programs significantly overlapped the set of HIF2a-responsive genes (**Figure 4h**). In contrast, neither program showed statistically significant overlap with HIF1a-specific signature (FDR > 0.05), which was further reflected by the absence of overlap between their top gene members and the HIF1a-signature (**Figure 4i**). The IER and oxidative stress response programs did not significantly overlap the signatures of any of the two HIFs from this work or previous works.

These results confirm that hypoxia-EMT and hypoxia-glycolysis programs are controlled by HIF2a in the ccRCC cell line model A498. The cell-to-cell variability of these programs in VHL-deficient ccRCCs, which are expected to have constitutively active HIFs, points to additional layers of regulation beyond canonical oxygen sensing^41^. To investigate whether the TME contributes to this heterogeneity, we analyzed a published single-cell-resolution spatial transcriptomics dataset of ccRCC tumors^44^. We quantified local malignant cell density, defined as the fraction of neighboring cells that are malignant (see **Methods**), and examined its relationship to gene program activity. Meta-analysis across 21 fields of view (FOV) (**Methods**) revealed a significant association between high malignant cell density and activation of the hypoxia-glycolysis program (FDR < 0.05; **Figure 4j,k**). Thus, this program is preferentially induced in malignant cells embedded within densely packed tumor regions, consistent with a model in which metabolic plasticity is dynamically tuned by microenvironmental context even in the absence of canonical oxygen sensing. In contrast, the hypoxia-EMT program did not exhibit a clear spatial pattern (**Figure 4j**), suggesting that its variability may be governed by distinct, non-microenvironmental regulatory mechanisms.

### A common set of gene programs underlie inter- and intra-tumor heterogeneity in renal cell carcinomas

Gene programs that underlie cellular diversity often mirror inter-tumor heterogeneity, highlighting recurrent transcriptional states that define cellular function at the micro level and tumor phenotypes at the macro level^45^. We observed a similar relationship in RCC, where a substantial portion of the inter-tumor heterogeneity across 889 RCCs spanning five molecular subtypes and 128 normal samples (from TCGA) could be explained by differential activity of the programs identified through analysis of cellular variability in cancer cells (program activities could be used to reconstruct genome-wide expression patterns, Pearson r = 0.96); **Supplementary Figure 16**). Furthermore, 55 gene programs had statistically significant subtype-specific activities (FDR < 0.05) (**Figure 5a**). For instance, hypoxia-EMT (GP60) and hypoxia-glycolysis (GP49) showed higher activity in ccRCCs, consistent with their HIF-driven activity (**Figure 5b**). In contrast, the IER (GP57) and oxidative stress response (GP22) programs, despite their overlap with existing hypoxia signaling signatures, did not show elevated activity in ccRCCs, in agreement with our siRNA experiments suggesting that these two programs are not driven by HIFs. In fact, the oxidative stress response program was more active in tRCC and tumors that co-cluster with normal tissues (**Figure 5b**). Other subtype-specific gene programs and their associated genes are explored in **Supplementary Figure 17**.

**Figure 5.**
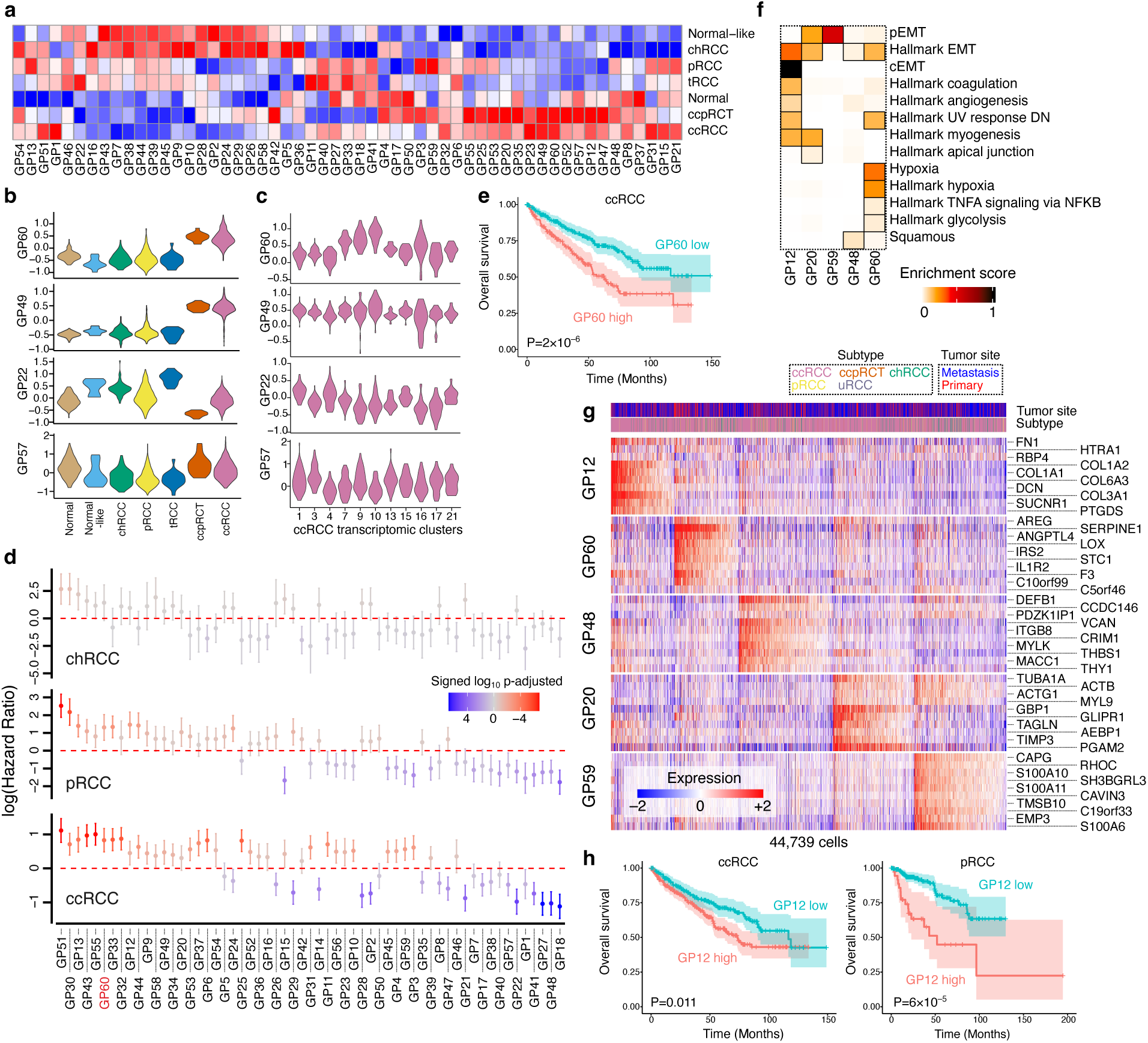
Inter-tumor heterogeneity in RCCs. (**a**) Heatmap of gene program activities across RCC subtypes and normal kidney samples from TCGA bulk RNA-seq profiles. Values represent column-wise z-scores of mean gene program activities (columns) across RCC subtypes (rows). (**b**) Violin plots showing the distribution of hypoxia-related gene program activities across RCC subtypes and normal samples from TCGA. (**c**) Same as (b) but showing hypoxia-related gene programs activities distributions across ccRCC transcriptomic clusters. (**d**) Overall survival analysis of RCC gene programs using TCGA data across the 3 major RCC subtypes. Plots show log hazard ratios with 95% confidence intervals from a Cox-regression model that included purity and sex as covariates. Color represents signed -log_10_ FDR values. See Supplementary Figure 18 for disease-free survival analysis. (**e**) Kaplan-Meier survival curve comparing overall survival of ccRCC tumors with high versus low GP60 activity in TCGA. Patients were stratified into two groups based on the optimal cutoff determined during testing. (**f**) Association between selected gene programs associated with EMT pathways. Gene sets include Hallmark pathways from MSigDB^40^ and cancer cell state signatures^46^. Heatmap shows enrichment scores; significant associations (FDR < 0.05) are outlined with a black box. (**g**) Intra-tumor heterogeneity of expression for five EMT-related gene programs. Heatmap shows normalized latent-log expression values from the scConvexNMF framework. Rows represent the top 10 member genes; columns represent individual cells. Top bar indicates metastasis status and RCC subtype for each cell. (**h**) Kaplan-Meier survival curves comparing overall survival of ccRCC (left) and pRCC (right) tumors with high versus low GP12 activity in TCGA.

Beyond subtype specificity, we also observed marked inter-patient heterogeneity in gene program activity within individual RCC subtypes. This intra-subtype variability often correlated with clinical outcome. For example, in contrast to the hypoxia-glycolysis program, which showed stable activity across ccRCC subclusters, the hypoxia-EMT program exhibits high variability across primary ccRCC tumors (**Figure 5c**) and is associated with poor prognosis (based on analysis of overall and disease-free survival; **Figure 5d-e** and **Supplementary Figure 18**). The oxidative stress response program showed an inverse trend and was associated with a favorable prognosis, further underlining the notion that previously reported hypoxia signature genes consist of multiple submodules with distinct biology, and clinical significance. Overall, we identified 52 programs that were significantly associated with outcome (FDR < 0.05) in at least one RCC subtype (**Figure 5d**).

We note that hypoxia-EMT is only one of five EMT-associated programs that we identified in our analysis of single-cell transcriptomes (**Figure 5f**,**g**). Consistent with findings in other cancer types^46^, one program (GP12) showed strong enrichment for members of a complete mesenchymal module (*FN1*, *COL1A1*, *COL1A2*), which we refer to as the complete EMT (cEMT) program, while another one (GP59) significantly overlapped with a partial mesenchymal module (*RHOC*, *S100A10*, *TMSB10*), designated as the partial EMT (pEMT) program. Notably, the cEMT program was associated with poor prognosis in both ccRCC and pRCC subtypes (**Figure 5h**), whereas the pEMT program was linked to adverse outcomes exclusively in ccRCC (**Supplementary Figure 19**), suggesting subtype-specific roles in disease progression. In contrast, another EMT-enriched program, GP48, was associated with improved prognosis in both ccRCC and pRCC (**Figure 5d**). Interestingly, this program showed high activity in normal tissue samples (**Figure 5a**). Despite current annotation of many of its genes as EMT signatures, GP48 predominantly comprises genes involved in epithelial tissue integrity and extra-cellular matrix organization (*DEFB1*, *ITGB8*, *THBS1*) (**Figure 5g**). These features suggest that GP48 represents an epithelial barrier integrity program, and that tumors with elevated activity of this program may retain more differentiated, epithelial-like characteristics. Other notable examples, including programs whose association with disease outcome depends on patient sex, are explored in **Supplementary Figure 19**.

### Transcriptional reprogramming marks early emergence of metastatic cell states in primary ccRCC

With a balanced representation of primary and metastatic tumors among our ccRCC samples (n=7+7), we set out to define cancer cell-intrinsic gene expression programs linked to metastasis. Principal component analysis (PCA) of pseudo-bulk malignant cell profiles revealed that the first principal component (PC1), explaining 20% of inter-tumor variation, was associated with metastasis status (Wilcoxon rank-sum test, P=0.053; **Figure 6a**). This suggests that metastatic transition is a major driver of transcriptional heterogeneity in ccRCC, with widespread remodeling of malignant cell states. Notably, this separation was independent of biosample type and could be seen in both patient tumors and PDXs (**Supplementary Figure 20a**), suggesting that observed differences between primary and metastatic tumors are driven by cancer cell-intrinsic mechanisms rather than tumor site.

**Figure 6.**
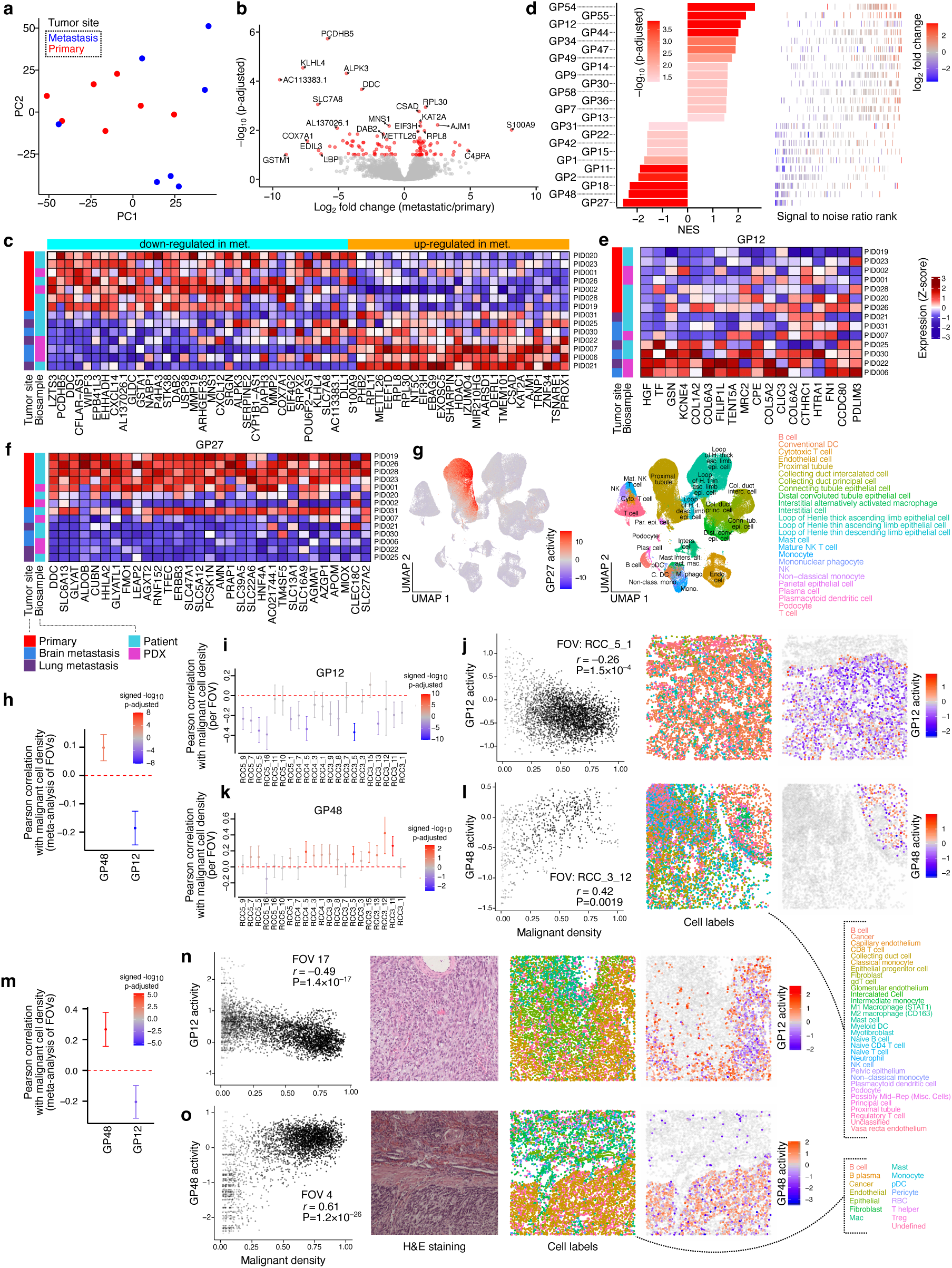
Gene programs characterizing transcriptomic differences between primary and metastatic tumors. (**a**) PCA of pseudo-bulk mRNA profiles from malignant ccRCC cells. Each dot represents one sample; color indicates metastatic status. See Supplementary Figure 20 for plot showing biosample type. (**b**) Volcano plot of differentially expressed genes between primary and metastatic ccRCC samples. Red dots indicate genes with FDR < 0.1. (**c**) Heatmap of differentially expressed genes between primary and metastatic ccRCC samples. Values represent column-wise z-scores of log_2_-normalized pseudo-bulk RNA-seq expression profiles for 51 genes (FDR < 0.05). Left annotation bar indicates metastatic status and bio type for each sample; top annotation bar indicates whether the gene was up- or downregulated. See Supplementary Figure 20 for visualization of 111 genes (FDR < 0.1). (**d**) Metastasis-associated gene expression programs. Left: bar plot showing normalized enrichment scores from GSEA for 24 programs with significant enrichment (FDR < 0.05). Right: signal-to noise-ratio ranking for genes within each program. For each gene program, member genes are shown as bars colored by log_2_ fold-change from differential expression analysis. (**e**) Heatmap of leading-edge genes for GP12, showing column-wise z-scores of log_2_-normalized pseudo-bulk RNA-seq ccRCC expression profiles. The left annotation bar indicates metastatic status and biosample type. (**f**) Same as (e), but for GP27. (**g**) Activity of leading-edge genes from GP27 in an atlas of single-cell expression profiles of kidney cells from the Kidney Precision Medicine Project^96^. UMAP embedding shows gene-set activity estimated using the ulm method from decoupleR^95^ (left) and cell type labels from the original study (right). (**h**) Meta-analysis of weighted correlation between local malignant cell density and activities of GP12 and GP48 in a single-cell spatial transcriptomic dataset^44^. Pearson correlation coefficients and 95% confidence intervals were derived via meta-analysis across FOVs. Color represents signed -log_10_ FDR values. Spatial analysis used a neighborhood-step of *k*=3 (see Supplementary Figure 15 for additional *k* values). (**i**) Weighted correlation analysis between malignant density and GP12 for individual FOVs. (**j**) Example of negative association between malignant density and GP12 activity. Left: scatterplot comparing GP12 activity and malignant density; dot intensity reflects weights representing local neighborhood support for both signals. Middle: spatial coordinates of cell centroids colored by cell type annotation from the original authors^44^. Right: spatial coordinates showing GP12 activity in malignant cells; non-malignant cells are transparent. (**k**) Weighted correlation analysis between malignant density and GP48 for individual FOVs. (**l**) Same as (j) but showing an example of positive association between malignant density and GP48 activity. (**m**) Meta-analysis of weighted correlation between local malignant cell density and activities of GP12 and GP48 in a single-cell resolution spatial transcriptomic map of a MTSCRCC tumor. Statistics are as in (h). (**n**) Example of negative association between malignant density and GP12 activity in a MTSCRCC tumor. Panel format follows (j), except that hematoxylin and eosin staining is also shown. (**o**) Same as (n) but showing an example of positive association between malignant density and GP48 activity.

Differential expression analysis of pseudobulk malignant cell profiles identified 111 genes significantly altered between primary and metastatic malignant cells (FDR < 0.1; **Figure 6b-c**, **Supplementary Figure 20b** and **Supplementary Data Table 4**). Many of these genes showed consistent up- or down-regulation across both lung and brain metastases, further reinforcing the notion that their modulation reflects cancer cell-intrinsic mechanisms rather than tissue-specific influences (**Supplementary Figure 20b**). To place these gene-level differences in a functional context, we performed gene set enrichment analysis (GSEA) across the 59 single-cell-derived GPs, which we described above. Notably, 24 out of 59 programs were significantly up- or down-regulated in metastatic tumors (FDR < 0.05), revealing extensive reorganization of malignant cell states associated with metastatic progression (**Figure 6d**).

In total, we identified 14 metastasis-associated programs, including a protein translation program (GP54), a cell-adhesion program (GP55), and the cEMT program (GP12, **Figure 6e**), but not the other four programs enriched for annotated EMT genes (**Supplementary Figure 21**). These changes point to enhanced protein biosynthesis, altered adhesion dynamics, and sustained cEMT activity as hallmarks of metastatic ccRCC. In contrast, primary tumors were enriched for renal lineage-associated programs: among the 10 primary-associated programs were GP27, which contains multiple solute transport genes (**Figure 6f**), GP18, a detoxification-related program, and GP48, an epithelial barrier integrity program (**Supplementary Figure 21**). Interestingly, in addition to primary tumors, these GPs are characteristically active in kidney proximal tubule cells, the cell of origin of ccRCC^3^, with GP27 in particular representing a highly specific marker of proximal tubule identity (**Figure 6g**), while GP18 and GP48 are more broadly expressed in multiple cell types of renal lineage (**Supplementary Figure 22**). The recurrent loss of these programs in metastatic tumors indicates that disease progression in ccRCC is accompanied by loss of proximal tubule identity and a transition toward a de-differentiated state.

Strikingly, key transcriptional changes linked to metastasis, including activation of the cEMT program (GP12) and loss of epithelial barrier integrity program (GP48), arise early and are already detectable in disseminating malignant cells at the invasive tumor front. Spatial analysis of ccRCC tumors^44^ revealed a significant negative correlation between cEMT program activity and local malignant cell density (**Figure 6h**,**i**), with elevated activity specifically in malignant cells, located at low-density regions near the tumor periphery (**Figure 6j**). This spatial enrichment suggests that cEMT activation marks early invasive states and may contribute to neoplastic cell dissemination, consistent with prior observations of EMT-related gene expression at tumor margins^16^. In contrast, GP48 activity showed a positive association with malignant cell density (**Figure 6k**) and was markedly reduced in disseminating cells (**Figure 6l**), indicating that the loss of epithelial gene programs, including epithelial barrier integrity program, is an early event in invasive transition.

To examine whether this spatial organization extends beyond ccRCC, we generated a single-cell-resolution spatial transcriptomics map from a mucinous tubular and spindle cell RCC (MTSCRCC) tumor (CosMx 1K-panel; see **Methods**), comprising 22 FOVs. Meta-analysis across FOVs confirmed the same spatial patterns observed in ccRCC, with cEMT activation enriched at the invasive front and GP48 expression diminished in disseminating malignant cells (**Figure 6m-o**). These findings demonstrate that early emergence of transcriptional hallmarks of metastasis, including cEMT activation and epithelial de-differentiation, and their spatial organization are not limited to ccRCC.

### Loss of proximal tubule identity defines an early metastatic transcriptional signature predictive of ccRCC outcomes

The erosion of proximal tubule identity in ccRCC metastatic cells was further evident when we derived a metastasis classifier directly from our single-cell data. Using significant differentially expressed (DE) genes between primary and metastatic tumors as features, we trained a logistic regression model to predict the metastatic status of individual malignant cells. Internal validation by leave-one-sample-out cross-validation demonstrated high sensitivity and specificity, with an area under the receiver operating characteristic curve (AUROC) of 0.912 (**Supplementary Figure 23**). Across iterations, a subset of 16 genes consistently contributed most strongly to model predictions (absolute mean Pearson > 0.1, **Figure 7a**), defining a compact metastasis signature. This gene panel effectively distinguished metastatic cells within our single-cell cohort (**Figure 7b**). External validation on bulk RNA-seq data from the TRACERx renal cohort^8^ confirmed the predictive power of this signature, with an AUROC of 0.709 (P = 0.01) for the 16-gene panel, demonstrating its robustness across independent datasets (**Supplementary Figure 24**).

**Figure 7.**
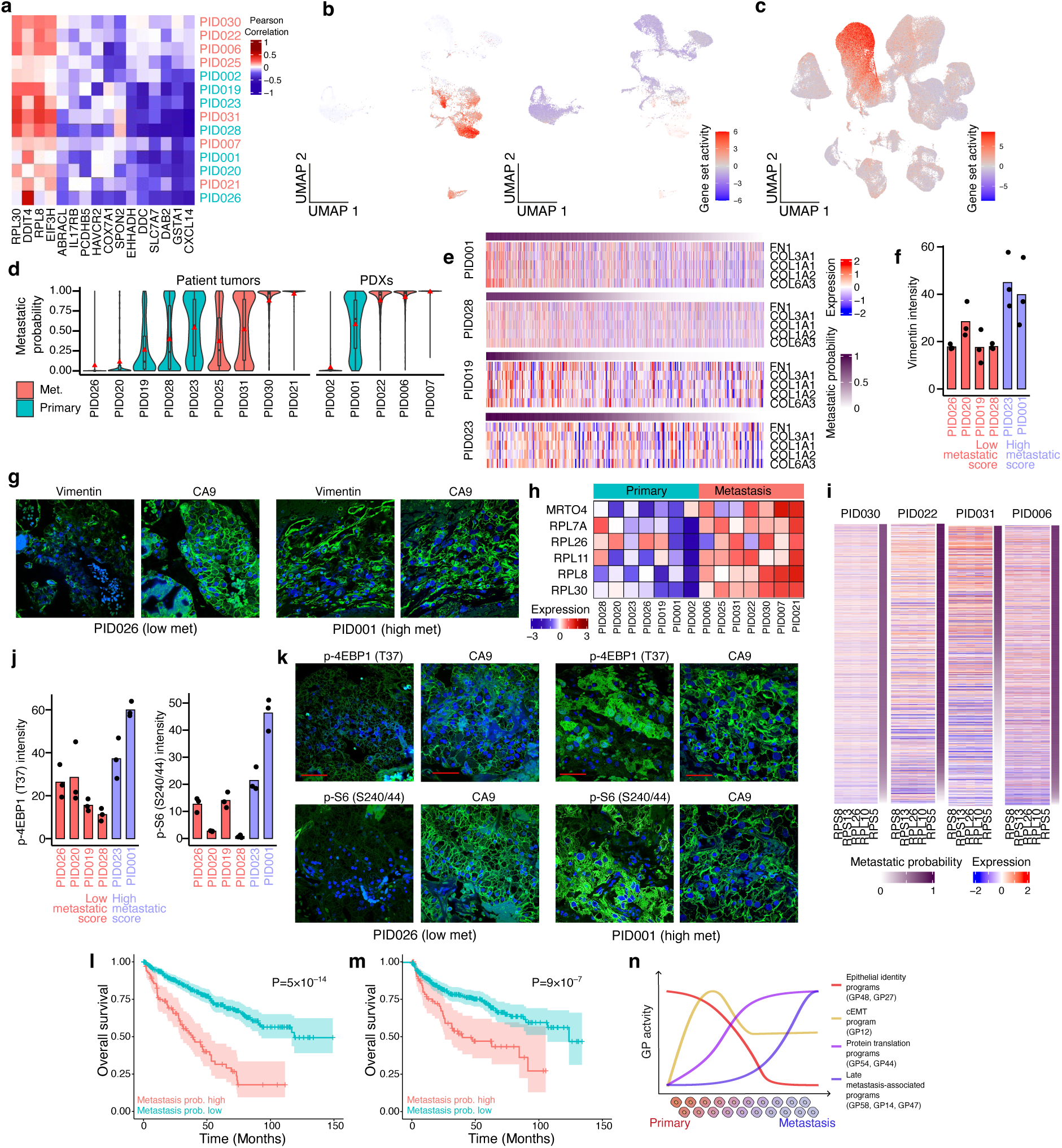
Metastasis signature and its relation to survival outcomes in ccRCC. (**a**) Heatmap showing the compact metastatic signature composed of genes contributing most strongly to model predictions (absolute mean Pearson > 0.1). Values represent Pearson correlation between log-normalized expression in malignant ccRCC cells and the full metastatic signature per donor. (**b**) Activity of proximal tubule identity genes distinguishes metastatic ccRCC cells. This gene set includes 12 members of the compact metastatic signature negatively associated with metastatic classification. UMAP embeddings were computed from rescored gene program activities in malignant ccRCC cells, based on the 24 programs differentially active between primary and metastatic tumors (Figure 6d). Gene set activity was estimated using the ulm method from decoupleR^95^. (**c**) Activity of proximal tubule identity genes in an atlas of single-cell expression profiles of kidney cells from the Kidney Precision Medicine Project^96^. UMAP embedding shows gene-set activity estimated using the ulm method from decoupleR^95^. (**d**) Distribution of metastasis signature scores per sample. Scores represent probabilities from an elastic net logistic regression model using leave-one-out sample cross-validation. Violin plots and boxplots show the distribution across malignant ccRCC cells; red triangle: mean, center line: median; box limits: upper and lower quartiles; whiskers: 1.5x the interquartile range. (**e**) Correlation between metastasis signature scores and the top five gene members of the cEMT program in ccRCC primary tumor samples showing heterogeneity in the metastasis signature. Each heatmap shows normalized latent-log expression values from the scConvexNMF framework. Top bar shows metastasis signature scores for each cell. (**f**) Quantification of immunostaining intensities for vimentin in ccRCC tumors with low and high metastatic probability score, together with CA9 staining at the corresponding regions. (**g**) Representative images of vimentin staining from a ccRCC tumor with high metastatic probability (PID001) and a tumor with low metastatic probability (PID026). Scale bar = 50 µm. (**h**) Heatmap of genes from the metastatic signature that overlap members of translation gene programs (GP54 and GP44), showing row-wise z-scores of log_2_-normalized pseudo-bulk RNA-seq ccRCC expression profiles. The top annotation bar indicates metastatic status. (**i**) Correlation between metastasis signature scores and the top five gene members of the protein translation program (GP54) in selected ccRCC metastatic samples. Each heatmap shows normalized latent-log expression values from the scConvexNMF framework. Right bar in each subpanel shows metastasis signature scores for each cell. (**j-k**) Same as (f-g) but showing immunostaining intensities for p-4EBP1 (T37) and p-S6 (S240/244). (**l**) Kaplan-Meier survival curves comparing overall survival of ccRCC tumors with high versus low full metastasis signature scores in TCGA. (**m**) Same as (l) but comparing survival using the compact signature score. (**n**) Schematic of temporal progression of gene programs during the primary to metastatic ccRCC transition.

Strikingly, 12 of these 16 genes were negatively associated with metastatic classification (**Figure 7a**) and showed highly specific expression in proximal tubule cells of the normal kidney (**Figure 7c**), reinforcing that loss of proximal tubule identity is a defining hallmark of metastatic ccRCC. These negative markers included lineage-specific genes such as *GSTA1*, a glutathione S-transferase critical for detoxification and defense against oxidative damage^47^, and *DDC*, an enzyme involved in dopamine biosynthesis with established roles in renal blood pressure and salt balance^48^. This signature also captured genes previously implicated in tumor progression, including *DAB2*, a phosphoprotein involved in endocytosis and in the protection of kidney tubular epithelial cells from TGF-β–induced EMT^49^, and *CXCL14*, a chemokine whose expression in ccRCC has been associated with improved survival after immunotherapy, suggesting a role in shaping the TME^50^. Together, these findings further strengthen the notion that metastatic progression in ccRCC is tightly coupled to the loss of proximal tubule-specific gene expression programs and the emergence of a distinct transcriptional state.

Consistent with our spatial analyses of gene programs, we found that metastasis-associated signatures were detectable even within primary tumors: in several primary ccRCC samples, malignant cells exhibited a broad distribution of metastasis signature scores, with a subset displaying high metastatic scores (**Figure 7d**)—this subset also express high levels of the cEMT program genes (**Figure 7e**). To assess whether this heterogeneity was also evident at the protein level, we performed immunostaining on primary tumors with low (n=4) and high (n=2) metastatic scores for vimentin, a marker of EMT and metastatic progression. As predicted, samples with high metastatic scores displayed elevated levels of vimentin (**Figure 7f**,**g**).

In addition to loss of proximal tubule identity, our metastatic signature captured genes belonging to protein translation programs that were upregulated in metastatic samples (**Figure 7h**). Notably, GP54, a protein translation program, showed a strong correlation with the metastatic signature across multiple metastatic samples (**Figure 7i**). Thus, we also examined the levels of p-4EBP1 (T37) and p-S6 (S240/244), downstream markers of mTOR signaling activation, which indicate increased levels of protein translation^51^, by immunostaining across primary tissues covering a range of metastatic potential scores. Indeed, both markers were significantly elevated in the metastasis-high subset of primary tumours (**Figure 7j-k**). These findings indicate that a subset of transcriptomic hallmarks of metastasis, including loss of epithelial identity and elevated protein translation, can emerge within subpopulations of primary tumor cells well before dissemination occurs, and are predictive of metastatic potential. Consistent with this, a primary tumor-derived PDX model with pronounced heterogeneity in metastasis scores (PID001) readily produced distant metastases in mice (**Supplementary Figure 25**), whereas another PDX model dominated by cells with low scores (PID002) did not, suggesting that the presence of high-scoring cells may forecast metastatic competence.

To test whether the detection of metastatic transcriptional states in primary tumors correlates with clinical outcomes, we applied our signature to bulk RNA-seq data from primary ccRCC tumors in TCGA^27^. High metastasis signature scores was strongly associated with worse overall survival (111-gene signature, Cox regression P < 10^−14^; **Figure 7l**), and this association remained highly significant even with the compact 16-gene panel (P < 10^−7^; **Figure 7m**). The prognostic value of the signature was independently validated in primary tumors from the TRACERx renal cohort (**Supplementary Figure 24**). Together, these findings demonstrate that advanced loss of proximal tubule identity, a defining feature of metastatic transcriptional states, can readily be detected in subsets of primary tumor cells and is strongly associated with adverse patient outcomes.

## Discussion

In this study, we characterize cellular diversity in RCC through the lens of transcriptional gene programs as the building blocks underlying malignant cell heterogeneity. While intra-tumor heterogeneity has traditionally been attributed to genetic evolution and microenvironmental influences^6,52^, our results demonstrate that a common set of transcriptional programs, discovered from within-clonal variability of cells, can explain both cell-to-cell variability and inter-tumor diversity across RCC subtypes. This convergence suggests that intra- and inter-tumor heterogeneity are not independent phenomena but instead reflect shared regulatory axes that govern malignant cell states. By explicitly accounting for clonal and copy number-driven shifts in gene expression, our framework provides a unified representation of tumor heterogeneity across scales, including both intra- and inter-tumor variability. In fact, we demonstrate that transcriptomic features can more accurately reflect inter-tumor variability than traditional histopathological annotations in RCC, as exemplified by the finding that many ccpRCTs are frequently misclassified, likely due to previous under-recognition, lack of specific biomarkers, as well as the overlapping morphological features with ccRCC^53^. We present CASP14 as a highly specific and sensitive biomarker with potential clinical utility for distinguishing ccpRCT cases from other RCC subtypes. Our findings suggest that incorporating CASP14 into diagnostic workflows can significantly improve ccpRCT classification and support better clinical management, especially in the biopsy setting in which distinguishing ccpRCT from ccRCC tumors is key.

While not explored in our main results, our data also shed light on the cell of origin of ccpRCT in addition to its diagnosis. Despite sharing substantial transcriptomic similarity with ccRCC, including activation of hypoxia-associated programs, we found that ccpRCT tumors exhibit low activity of proximal tubule-associated gene programs (GP27, GP18, GP21), indicating that this subtype is unlikely to originate from proximal tubule cells (**Supplementary Figure 26a**). Instead, ccpRCT tumors display strong similarity to distal tubule cell populations, particularly principal cells (PC), with cells from the inner medullary collecting duct (IMCD) and collecting duct (CCD-PC) showing the highest similarity (**Supplementary Figure 26b**). This interpretation is further supported by the expression of GATA3^54^ in ccpRCT tumors, a marker of the distal nephron and collecting system, collectively supporting a distal nephron origin for ccpRCT. Our data also reinforces the consensus on cell of origin of other RCC subtypes^55–57^, with proximal tubule cells and distal tubule cells as the most likely cell of origin for ccRCC and chRCC, respectively (**Supplementary Figure 26c**). Notably, among proximal tubule populations, proximal tubule segment 3 (PT-S3) showed the highest transcriptional similarity to ccRCC cells. PT-S3 cells are known to be particularly susceptible to ischemic injury^58^ and have been implicated in regenerative responses^59^, suggesting that this population may be especially prone to malignant transformation.

Our systematic analysis of gene programs provide evidence that canonical cancer pathways are not monolithic, but instead comprise multiple functionally distinct submodules with divergent activities and clinical associations. This is exemplified by the hypoxia signaling pathway, which we resolve into four separate gene programs with distinct functional and regulatory properties. Pseudo-hypoxic state is a hallmark of ccRCC, emerging from early VHL inactivation and the resulting accumulation of HIF2^60^. However, only two of the hypoxia submodules that we identified, including a hypoxia-EMT program, are driven by HIF2 activity, while the others reflect immediate-early and oxidative stress responses, underscoring the composite nature of widely used pathway signatures. Moreover, the substantial variability of HIF2-driven programs across VHL-deficient cancer cells is unexpected under the conventional view that constitutive VHL inactivation produces a uniform pseudohypoxic state, and suggests the existence of additional regulatory mechanisms that modulate HIF2 activity independently of VHL. Supporting this notion, in a separate work^61^, we show that lysyl oxidase (LOX) is a previously unrecognized regulator of HIF2α protein stability. Notably, *LOX* itself is a component of the hypoxia-EMT program (**Figure 4g**), indicating that variation in *LOX* expression is coupled to the activity of other genes within this program and implicating LOX as a likely regulator of its heterogeneous activity. Importantly, pharmacological inhibition of LOX attenuates the HIF2 program and reduces metastatic burden^61^, highlighting the therapeutic potential of targeting gene program regulators. More broadly, this modular view provides a framework for dissecting complex transcriptional phenotypes and identifying candidate regulators with therapeutic potential.

Our analysis of metastatic progression in ccRCC further reveals that transcriptional reprogramming involves multiple gene programs and follows a temporally ordered trajectory. We identify early activation of a cEMT program and concurrent loss of epithelial identity at the invasive tumor front, indicating that invasive potential emerges within primary tumors prior to dissemination. The progressive loss of proximal tubule identity, on the other hand, likely facilitates metastatic progression by promoting a more plastic, stem-like transcriptional state and relaxing lineage constraints that may otherwise limit dissemination and colonization, features that have been broadly associated with metastatic competence in cancer^62–64^. These early changes are followed by broader transcriptional remodeling in metastatic lesions, including activation of additional programs that are not associated with survival or spatial organization in primary tumors, suggesting a distinction between early invasive states and later metastasis-specific adaptations (**Figure 7n**). The observation that the spatial organization of early metastatic states are conserved across RCC subtypes further supports a shared regulatory logic governing invasive behavior.

In addition to these early transitions, we observed increased activation of protein translation programs in metastatic ccRCC (**Figure 6d**). While enhanced translation is often interpreted as a consequence of increased proliferative demand^65^, our findings suggest a more active role in metastatic reprogramming, as translation programs are activated concurrently with EMT (**Figure 7**), with immunostaining analysis confirming their early activation specifically in primary tumor models with high metastatic potential (**Figure 7j-k**). Importantly, we observe that metastasis-associated translation programs (GP54 and GP44) are enriched for targets of MYC (**Figure 4e**), a key driver of ribosome biogenesis^66^ whose activity is known to be augmented downstream of HIF2α^67^, supporting a model in which hypoxia-MYC signaling contributes to the upregulation of specific submodules of the translation machinery during metastatic reprogramming. This model is consistent with emerging evidence linking EMT initiation and increased transcription of ribosomal RNAs (rRNA), whose inhibition reduces metastasis^68^. Furthermore, accumulating evidence suggests that ribosomes exhibit compositional heterogeneity in ribosomal proteins and rRNAs^69^ with pro-metastatic functions^70^. These observations nominate protein translation as a key axis of metastatic reprogramming.

Although our study focuses on mapping transcriptomic heterogeneity in malignant cells, our data and gene modeling framework are also applicable to profiling non-malignant cell populations in RCC tumors. As an example, we examined the transcriptional heterogeneity of tumor-associated macrophages (TAMs) to assess how immune GPs shape disease progression and tumor-immune interactions. Our analysis identified GPs that capture transcriptomic variations among TAMs (**Supplementary Figure 27** and **Supplementary Data Table 5**), with several prognostic programs and programs enriched or depleted in metastasis (**Supplementary Figure 28**). Notably, we observe spatial organization of TAM states within primary RCC tumors (**Supplementary Figure 29**). For example, an M2-like *SPP1* program (GP31) is positively correlated with proximity to malignant cells in the tumor core, indicative of tumor infiltration and consistent with observations that *SPP1*-expressing macrophages localize to hypoxic tumor regions^71,72^. Of note, *SPP1*-expressing macrophages are reported to be enriched in a “suppressed mixed immune” microenvironment associated with poor outcome^73^. Interestingly, in our data, this M2-like *SPP1* program shows significant enrichment in metastatic tumors, suggesting that metastatic tumors enrich for suppressive immune programs. Other TAM gene programs, their spatial organization, and association with metastasis or disease outcome are explored in **Supplementary Figure 27-29.**

In summary, our study establishes a gene program-centric framework for interpreting malignant cell heterogeneity in RCC, demonstrating that a shared set of transcriptional programs underlies tumor identity, evolution, and clinical behavior. By linking cell-intrinsic variability to inter-tumor diversity, refining subtype classification, and defining transcriptional trajectories associated with metastasis, this work provides a generalizable framework for studying tumor heterogeneity across RCCs. More broadly, these findings suggest that resolving complex transcriptional phenotypes into modular regulatory programs may enable more precise characterization of tumor states and facilitate the identification of actionable vulnerabilities in cancer.

## Methods

### Patient samples

Tumor tissues were available through the McGill University Health Centre (MUHC)/McGill RCC biobank. Lung metastases were obtained from the Thoracic Oncology Clinical Database and Biobank (MUHC REB# 2014-1119). Tumors were collected from patients undergoing nephrectomy, after obtaining informed written consent. Pathology evaluations of the tumor tissues and diagnosis of RCC subtypes were completed by a Genitourinary pathologist at the MUHC. Ethical approval was obtained from the McGill University Health Centre Research Ethics Board (MUHC REB). No blinding or statistical power calculations were performed in this study.

### Generation of Patient derived xenograft models

Patient-Derived xenograft models were generated by implanting a tumor fragment (approximately 1mm x 1mm, up to 3mm x 3mm) obtained from resected human renal carcinoma specimens into the right flank of 8–12-week-old male Non-Obese Diabetic severe combined immunodeficient gamma (NSG) mice [Cg-Prkdc scid Il2rg tm1Wjl /SzJ; The Jackson Laboratory, stock no. 005557]. Mice were monitored twice a week, and tumor size was measured once a week. Mice were euthanized when tumor volume reached 20,000 mm^3^ or upon meeting clinical or experimental endpoints. Euthanasia was performed under isoflurane anesthesia followed by CO_2_ inhalation and cervical dislocation. All animal experiments were conducted in accordance with institutional and national guidelines for animal care and use and were approved by the McGill University Animal Care Committee (protocols #7514 and #4830).

### Single-cell preparation

Tumors were dissociated to single cells through the following steps: the tumor was washed with 5-10 ml Dulbecco’s Phosphate-Buffered Saline (DPBS; Gibco, Cat# 14190-136) 2-3 times and placed on a petri dish. 1ml DPBS was added on the tumor tissue to avoid drying. The tumor was cut into small 1-2 mm pieces with scalpel No. 10 (Andwin Scientific, Cat# NC9999403). 9 ml DPBS was added into the petri dish and the pieces were collected in a 50 ml centrifuge tube with a 5 ml serological pipet and centrifuged at 300 x g for 5 min. The pieces were washed with 5-10 ml DPBS 1 or 2 times. The human tumor dissociation kit (Miltenyi Biotec, Cat# 130-095-929, Bergisch Gladbach, Germany) enzymes were prepared according to the provided instructions. Enzyme solution was prepared by adding Enzymes H (200 µl), R (100 µl), and A (25 µl) to 4.7 ml Roswell Park Memorial Institute (RPMI) Medium 1640 (1X) (Gibco, Cat# 11875093). After centrifugation, supernatant was discarded and tumor pieces were re-suspended in the enzyme solution, and transferred into the gentleMACS C tubes (Miltenyi Biotec, Cat# 130-093-237, Bergisch Gladbach, Germany). gentleMACS C tube were attached upside down onto the sleeve of the gentleMACS Octo Dissociator with Heaters (Miltenyi Biotec, Cat# 130-096-427, Bergisch Gladbach, Germany). Tumor tissue was dissociated using 37_h_TDK_1 program. When the program was completed, a short centrifuge was done to collect cells at the bottom of the gentleMACS C tube. All materials were collected by a 10 ml serological pipette, passed through a 70 µM strainer drop-by-drop (UltiDent, Cat# 229483) and transferred to a 15 ml centrifuge tube. 4-5 ml DPBS was added on the strainer to collect the remaining cells. Cells were centrifuged at 300 x g for 7 minutes and supernatant was discarded. To remove red blood cells, the cell pellet was re-suspended in 3 ml ACK lysing buffer (Gibco, Cat# A10492-01) and incubated at room temperature for 3 minutes. DPBS was then added to the cell suspension to fill the tube to a final volume of 15 ml and centrifuged at 300 x g for 5 minutes. Supernatant was discarded and cells were resuspended in 1-2 ml 1% BSA in DPBS. Cells were filtered through 40 µM Flowmi^TM^ cell strainer (Bel-Art, Cat# 136800040) to remove large particles. Then the cells were counted using TC20^TM^ automated cell counter (Bio-Rad, Cat# 1450102, USA) following mixing 10-100 µl of cell suspensions with the same amount (10-100 µl) of Trypan Blue Stain (0.4%) (Gibco, Cat# 15250-061, Thermo Fisher Scientific, USA). After 3 minutes, 10 µl of mixture was loaded into cell counting slides (Bio-Rad, Cat# 145-0011, USA) and inserted into TC20^TM^.

In the case of PDX models, mouse cells were removed using Mouse Cell Depletion Kit (Miltenyi Biotec, Cat# 130-104-694, Bergisch Gladbach, Germany) according to the manufacturer protocol. Cells were centrifuged at 30 x g for 10 minutes. For 2×10^6^ cancer cells (10^7^ total cells) or less, the cell pellet was resuspended in 80 µL 0.5% BSA in DPBS buffer, and 20 µL of mouse cell depletion beads was added and incubated in refrigerator (2-8 C) for 15 minutes. Then 400 µL 0.5% BSA in DPBS buffer was added to the cell suspension. For more cells, the amount of each reagent was scaled up. LS column (Miltenyi Biotec, Cat# 130-042-401, Bergisch Gladbach, Germany) were attached to QuadroMACS™ Separator (Miltenyi Biotec, Cat#130-091-051, Bergisch Gladbach, Germany) and rinsed with 3ml buffer. The collecting tube was changed after rinsing, and cell suspension was applied to the column and collected. 1 ml 0.5% BSA in DPBS buffer was added to the column twice and collected in the same tube. After removing mouse cells, cells were counted with TC20^TM^ automated cell counter.

### 10x library preparation and sequencing

Quality control of cell suspension and generation sequence ready libraries were performed according to the manufacturers’ user guides by experienced lab personnel at the McGill Applied Genomics Innovation Core, as described below.

An aliquot was taken from each cell suspension and incubated in live-dead staining with working concentration of 2µM calcein-AM and 4µM Ethidium-Homodimer1 (Thermo Fisher Scientific, Cat# L3224). After 10 minutes of room temperature incubation, the stained samples were loaded onto hemocytometer (Incyto, Cat# DHC-N01-5) and imaged on bright field, GFP (for Calcein-AM) and RFP (for Ethidium homodimer-1) channels using an EVOS FL Auto Imaging System (Thermo Fisher Scientific, Waltham, MA, USA). Sample viability, concentration, segregation and absence of large debris were determined from the resulting microscopy images.

The single cell cDNA libraries were generated following Chromium Single Cell 3’ Reagent Kits User Guide (10X Genomics). Briefly, samples that passed quality control were suspended into Reverse Transcription (RT) Master Mix then pipetted into 10X genomics chip Well-1, followed by Gel Beads into Well-2 and Partition oil into output well. The chip assembly was run on a Chromium Controller which generated Gel Bead-In-EMulsions (GEMs). GEMs were pipetted out from the Chip and into a 200µL PCR tube (Eppendorf, Hamburg, Germany, Cat# 951010022) and ran on T100^TM^ Thermal Cycler (Bio-Rad Laboratories, Hercules, CA, USA, Cat# 1861096) with RT protocol [45min at 53°C, 5min at 85°C, hold at 4°C]. After RT, the GEMs content was released using Recovery Reagent and cDNA were isolated using Buffer Sample Clean Up 1 (10X Genomics) containing Silane Dynabeads (Thermo Fisher Scientific, Cat# 2000048). The purified cDNA was PCR amplified followed by purification using 0.6X volume SPRIselect beads (Beckman Coulter, Cat# B23318). The cDNA quality (size distribution) and concentration were assessed using the LabChip (Perkin Elmer Cat# 760517, CLS760672). A one-step Fragmentation, End Repair & A-tailing mix (10X genomics) were added to the cDNA in a 200µL PCR tube and ran on a thermocycler with the protocol [hold at 4°C, 5min at 32°C, 30min at 65°C, hold at 4°C]. The fragmented cDNA was subjected to double-sided size selection using SPRIselect bead, by first suspending the fragmented cDNA in 0.6X volume of SPRIselect for 5 minutes, using 10x Magnetic Separator to pull down the beads, moving the suspension into another PCR tube and topped with 0.8X volume of SPRIselect. Following two rounds of 80% ethanol washes, the desired sized fragmented cDNA was eluted into adaptor ligation mix (10X genomics) and incubated for 15 minutes at 20°C. The ligated product was cleaned with 0.8X volume of SPRIselect, added to Sample Index PCR Mix (10X genomics) and amplified via 12-14 PCR cycles depending on the amount of full-length cDNA as input. The final PCR product (or sequence ready library) was purified as done with fragmented cDNA and quality controlled using Lab Chip, as described earlier. The libraries were sequenced on an Illumina NovaSeq 6000 or MGI platforms to a targeted number of 20,000 reads per cell.

### Pre-processing and quality control of scRNA-seq data

Raw sequencing reads were aligned to reference genomes using the 10x Genomics CellRanger software^74^ (v3.0.2). For tumor samples, reads were aligned to the human reference genome (hg38). For PDX samples, a combined human (hg38) and mouse (mm10) reference was used. Unique molecular identifier (UMI) counts were generated by collapsing alignments using CellRanger. For PDX samples, multiplets and mouse cells were removed as identified by the CellRanger pipeline. Only cells with >95% of counts mapped to the human genome were retained. Subsequently, only counts assigned to the human reference were used for downstream analysis.

Quality control metrics were computed using the scuttle^75^ (v.1.0.4) package. Cells were retained if they met the following criteria: log_10_ total counts >3, log_10_ total features >3, and percentage of mitochondrial reads per cell <20%. Doublet detection was performed using DoubletFinder^76^ (v.2.0.3). The doublet rate was estimated based on the number of cells recovered per sample, following the multiplet rate guidelines from the 10x Genomics v3 user guide. Other parameters remained as default, except that the top 50 principal components were used for dimensional reduction. After QC, the RCC Atlas data set contained 85,563 cells. Log_2_-normalized expression values were obtained using the multiBatchNorm function from the batchelor^77^ package (v1.6.3), and unless otherwise specified, these log_2_-normalized counts were used for downstream analyses.

### Copy number variations inference and clonal analysis

Malignant and non-malignant cells were identified using inferCNV^78^ (v.1.2.1). As a reference for normal expression, we included healthy kidney from patient 1 from Young et al 2018^55^. We ran inferCNV separately for each sample, using the following parameters: cutoff = 0.1, cluster_by_groups=F, denoise=T and HMM=T, with all other parameters set to default.

To classify malignant cells, we used the residual expression matrix from the inferCNV object as input for clustering. For each sample, dimensional reduction was performed using the first 30 principal components, followed by construction of a nearest-neighbor graph. Clustering was performed using the ‘cluster_walktrap’ and ‘cut_at’ functions from the igraph^79^ package (v.1.3.4). Residual expression was then summarized at the chromosome-arm level for each cell by calculating the median expression across genes. Malignant and non-malignant clusters were annotated based on visualization of chromosome-arm level profiles across multiple arms.

Copy number variations and tumor sub-clones were inferred using Numbat^80^ (v.1.4.0) with default parameters. Allele counting and SNP phasing were performed using the 1000 Genome reference. The same normal gene expression reference that we used for InferCNV was used.

### Cell type annotation

Clustering analysis was performed using the Seurat^81^ (v.5.03) pipeline. After normalization and scaling steps, highly variable genes were identified using the FindVariableFeatures function with the ‘vst’ method. Mitochondrial and ribosomal genes (matching the patterns (‘^MT-|^RP[SL]’) were excluded from downstream analyses. PCA was conducted on the filtered set of highly variable genes, and the top 40 principal components were selected for dimensionality reduction. Clustering was performed on a shared nearest neighbor (SNN) graph using the Louvain algorithm, as implemented in the FindClusters function.

Based on the resulting clusters, major cellular compartments, including epithelial, immune and stromal populations, were manually annotated by examining the expression of canonical marker genes (**Supplementary Data Table 6**). Immune and stromal compartments were subsequently subsetted and re-clustered using an additional round of SNN clustering to further resolve these subpopulations, leading to the identification of broad immune and stromal clusters (**Figure 1b,c**). Clusters with ambiguous or mixed marker expression profiles were labelled as *Unclassified*.

### Data set integration

Integration across samples was performed using GEDI^34^ (v.1.0.1) on the raw count matrices, specifying the number of latent variables as K=20, and leaving all other hyperparameters as default unless otherwise noted. For the integration of RCC patient tumor cells, sample-level information about metastasis status was incorporated into the model via the H parameter. In this analysis, the integrated embedding was generated by performing UMAP to the projection of the transcriptomic vector field of metastasis learned by GEDI, with cell positions representing the manifold embedding for the primary condition. GEDI was also used to generate an embedding of malignant cells across all tumors and PDX models. For this analysis, no integration was enforced, setting the distortion parameters oi_shrinkage and Qi_shrinkage to 100.

### Transcriptomic reclassification of bulk RNA sequencing data of different RCC subtypes from The Cancer Genome Atlas

The count matrix and metadata for KIRC, KIRP, and KICH from TCGA were downloaded via cBioPortal. Genes were annotated using GENCODE v22. Genes with identical symbols were merged, and those with a mean count less than 1 in each tissue type were excluded from further analysis. Based on transcriptomic patterns and the expression of clinical markers, we identified and manually corrected one case in which the tumor and matched normal sample labels were swapped (TCGA-CW-5591-01A & -11A).

The filtered count data were log-normalized using Seurat’s NormalizeData function with a scaling factor of 100,000. The top 12,000 variable genes were identified using the ‘vst’ method, of which 10,778 were then used to calculate principal components after filtering out 1,222 genes that (a) were markers of normal kidney cells based on data from Su *et al* 2021^82^ and McEvoy *et al* 2022^83^ or (b) were markers of at least one immune or stromal cell type in our single-cell data (see **Supplementary Data Table 7** for the set of genes that were retained). To perform this filtering, normal kidney data from McEvoy *et al* 2022^83^ and one paired chRCC and normal sample from Su *et al* 2021^82^ were analyzed jointly with the RCC dataset. Briefly, the merged object was processed using the Seurat pipeline, clustering was performed using the FindClusters function, and markers associated with non-epithelial populations were identified using FindMarkers. The filtered data were subsequently scaled using the ScaleData function. The top 36 principal components were selected based on elbow plots and JackStraw analysis and used to cluster samples by SNN graph construction followed the Louvain clustering with multilevel refinement with resolution 4, followed by hierarchical grouping of Louvain clusters as described in the main text.

Tumor samples were reclassified at the cluster level by generating cancer cell-intrinsic gene signatures for the ccRCC, chRCC and ccpRCT subtypes. To derive these signatures, we combined our single-cell expression data with datasets from Su *et al* 2021^82^ and McEvoy *et al* 2022^83^. The merged object was processed using the Seurat pipeline, including NormalizeData, FindVariableFeatures, ScaleData, RunPCA, FindNeighbors and FindClusters steps, with the objective of obtaining clusters that grouped malignant cells according to subtype. Marker genes for each subtype-associated cluster were then identified using the FindMarkers function. Candidate signature genes were selected by filtering for markers that met the following criteria: adjusted *P* value < 0.05, log_2_ fold change > 1 and percentage of expressing samples > 70%. Final gene selection for each subtype was refined by visual inspection of violin plots. The resulting subtype-specific signature gene lists are provided in **Supplementary Data Table 8**.

For pRCC and tRCC, we first focused on identifying and evaluating markers from the literature, given the heterogeneity of pRCC and that our cohort did not include tRCC. In the latest WHO classification (5^th^ edition, 2022), pRCC is considered as a single entity, whereas previous classifications distinguished type 1 and type 2 pRCCs based on different histopathological features. To better reflect this heterogeneity, we collected candidate mRNA markers from previous studies (PMID: 23887297^84^, PMID: 32150988^85^, PMID: 38703764^86^) and selected the most representative markers that were positive in pRCC and negative in most of the other subtypes/cell types. Similarly, the tRCC signature was generated by reviewing markers reported in previous publications (PMID: 34986355^30^, PMID: 33854184^31^, PMID: 39085357^32^, PMID: 38104891^33^) and refining them based on single-cell expression patterns.

Finally, cluster 20 contained nine tumor samples originally diagnosed as pRCC or chRCC, along with normal tissue samples. These tumors did not show high activity of other subtype-specific signatures and exhibited few genomic alterations; therefore, they were annotated as normal-like.

### Genomic analysis

Somatic mutation and copy number alteration data for the ccRCC, pRCC, and chRCC cohorts from TCGA were downloaded from cBioPortal and processed in R using the maftools^87^ (v.2.16.0) package. For SNV analysis, mutation annotation format (MAF) files of each cohort were merged and samples with matched RNA-seq data were used for the analysis. Variant classifications were binarized for the purpose of visualization excluding mutations classified as ‘Intron’ and ‘Silent’.

For CNV analysis, gene-level CNV data were aggregated to infer chromosome arm- or whole chromosome-level alterations. CNV status for each gene (-2 to 2 for deletion to amplification) was averaged across each arm or chromosome, and frequencies of arm- or chromosome-level CNVs were classified as deletion, loss, neutral, gain, and amplification based on the mean scores by the following thresholds: -1.3, -0.3, 0, 0.3, and 1.3.

To identify microphthalmia-associated transcription factor family (MiT) fusion gene status, StarFusion^88^ (v.1.13.0) was used. Raw sequencing data in FASTQ format were aligned to the hg38 reference genome, and the following parameters were included in the STAR-Fusion command: --FusionInspector validate, --examine_coding_effect and – denovo_reconstruct. From the fusion analysis, samples harboring fusion genes involving *TFE3*, *TFEB*, *TFEC*, or *MITF* were annotated as ‘MiT fusion’. In addition to our fusion analysis, samples that have previously been annotated as MiT fusion by Bakouny *et al*^30^ were also included. Oncoplots were generated using the ‘oncoplot’ function from maftools to display genomic alterations in the genes that are frequently altered in major RCC subtypes as well as most frequently mutated genes in other subtypes re-classified by our transcriptomic analysis.

### Development of HIF1A and HIF2A activity signatures using gene perturbation in ccRCC

A498 cell line was purchased from ATCC and maintained in RPMI 1640 supplemented with 10% fetal bovine serum and 1% penicillin-streptomycin at 37°C in a humidified incubator with 5% CO₂. For knockdown of *HIF1A* and *EPAS1*, reverse siRNA transfection was performed using Lipofectamine RNAiMAX (Thermo Fisher Scientific) according to the manufacturer’s instructions. Cells were transfected with siRNA targeting HIF1A, HIF2A (EPAS1), or a non-targeting control siRNA (Qiagen) at a final concentration of 10 nM. Transfection was carried out in 6-well plates with 2.5 × 10⁵ cells per well. Total RNA was extracted 48 hours post-transfection using the RNeasy Mini Kit (Qiagen) following the manufacturer’s protocol. RNA quality and integrity were assessed using a Bioanalyzer. Samples with RNA Integrity Number (RIN) ≥ 8.0 were used for library preparation. RNA-seq libraries were prepared using the TruSeq Stranded mRNA Library Prep Kit (Illumina) and sequenced on an Illumina NovaSeq 6000 platform to generate 150 bp paired-end reads (need to be confirmed). Raw sequencing reads were aligned to the human reference genome (GRCh38) using HISAT2^89^ (v.2.2.1). Gene-level read counts were obtained using featureCounts^90^ (v.2.0.3). Differential gene expression analysis was performed using the DESeq2 package. To model the effects of HIF1A and EPAS1(HIF2A) knockdown on gene expression, the design formula ∼ HIF1_status + HIF2_status was applied. The analysis included four experimental conditions: control, HIF1A-KD, EPAS1-KD, and double-KD, each with three biological replicates. To derive HIF1A and HIF2a gene signatures, significantly differentially expressed genes (p-adj < 0.05 & FC > 2) were selected, excluding *HIF1A* and *EPAS1* themselves. Additionally, 5 genes that overlapped between the two signatures (*IGFBP3*, *CDH3*, *BX640514.2*, *RTL9* and *PADI1*) were excluded from the final signature sets.

### Single-cell convex non-negative matrix factorization

We use convex non-negative matrix factorization (convex NMF^39^) to decompose single-cell gene expression profiles into a set of gene programs, each of which representing a convex combination of multiple tightly co-expressed genes. Convex NMF has properties that make it suitable for gene program analysis, including (a) sparse decomposition^39^, (b) the ability to model both positive and negative values, which, in contrast to NMF, enables analysis of log-scaled expression profiles, and (c) decomposition of the expression matrix into low-rank factors that represent a convex (non-negative weighted) combination of genes, enabling direct interpretation of gene programs and computation of their activities from the expression of their underlying genes. We implemented convex NMF decomposition as a generative model with data-generating distributions tailored toward sparse single-cell data analysis^34^, while taking into account the effects of clonal gene expression shifts to remove the confounding effect of inter-tumour and inter-clonal heterogeneity. Thus, the gene programs that are identified represent gene sets that show coordinated intra-tumour and intra-clonal variability.

Consider the single-cell gene expression dataset with measurements for *G* genes in *N* cells. Each cell *i* (*i*∈{1,…,*N*}), with the (latent, log-scaled) mRNA abundance profile **y***_i_*, belongs to one of *K* clonal groups (which may come from the same or different samples). We denote the clonal group of cell *i* as *k*(*i*). For each gene *j* (*j*∈{1,…,*J*}) in each cell *i*, the observed UMI count *m_ij_* is modeled as a Poisson function of the underlying mRNA abundance *y_ij_* in addition to a cell-specific size factor *l_i_*:

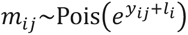

We denote the clone-specific contributions to the expression of gene *j* in cell *i* as *θ_j_*_,*k*(*i*)_, representing the average shift in expression of gene *j* in the clonal group to which cell *i* belongs (*θ_j_*_,*k*(*i*)_∈ℝ). Thus, the expression of gene *j* in cell *i* after subtracting clonal gene-level effects is:

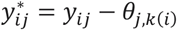

These clone-wise normalized expression values form the matrix **Y**\*∈ℝ^N×J^. We approximate this matrix with the lower-rank matrix **Ŷ** (rank *P*) through convex NMF as:

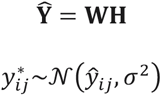

where **H** is a non-negative matrix (**H**∈**ℝ**_+_^P×J^), **G** is the pseudoinverse of **H,** and **W** is a matrix whose columns are restricted to be convex combinations of the columns of **Y**\* (**W**∈**ℝ**^N×P^):

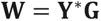

Together, these equations form a generative model, in which the latent, log-scaled, clone-wise normalized expression profiles of cells are approximated with convex NMF on one hand, and determine the parameters of the generative distribution of UMI counts on the other hand (**Supplementary Figure 8**).

The parameters of this model are fitted to the observed data using an expectation-maximization (EM) approach previously described^34^. Briefly, in each iteration of the EM algorithm, we estimate the mean of the posterior distribution of each element of **Y**\*, given the observed UMI counts and the current model parameters, using Laplace approximation as described previously^34^. Then, *l_i_* is updated for each individual cell, and *θ_j_*_,*k*_ for each individual gene in each clonal group *k*, as:

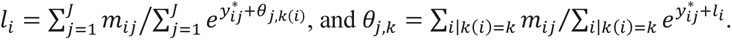

This is followed by updating **G** as the pseudoinverse of **H**^39^, updating **W** based on the new **Y**\* and **G**, and updating **H** based on the new **Y**\* and **W** using non-negative least-squares regression. To enforce a unique solution, we also restrict **Y**\* to have row- and column-wise means of zero, and **G** to have column-wise sums of 1. The latter also forces each column of W to be the weighted mean of the expression profiles of the set of genes that make up the corresponding gene program; this weighted mean is interpreted as the “activity” of that gene program in each cell.

After fitting the model to our single-cell data, to calculate the activity of the gene programs in a new dataset, we follow the exact same procedure above, except that **G** is not updated and is kept the same as the **G** fitted to the single-cell data; thus, **W** columns remain the weighted average of the normalized expression profiles of genes involved in each gene program, with weights learned from our single-cell data.

This model was fitted to each cell type in our dataset separately. For malignant cells, we considered the intersection of cells that were identified as malignant by InferCNV and those identified as malignant and assigned to a tumor clone by Numbat. We also included only samples with more than 100 cancer cells, resulting in analysis of 44,739 malignant cancer cells across 19 samples. In non-malignant cells, each “clonal group” was simply defined as cells of the same cell type obtained from one sample.

To determine the rank *P* (i.e., the number of gene programs), we performed internal cross-validation by randomly withholding 1% of the entries in the original count matrix during model fitting using the maskingRatio parameter. Masked entries were excluded from parameter estimation and later used to quantify predictive performance. Candidate ranks from 10 to 100, in increments of 10, were evaluated according to two criteria: (i) goodness of fit and (ii) biological interpretability (**Supplementary Figure 9**).

For evaluating goodness of fit, we computed for each rank the Poisson log-likelihood of the observed counts under two models: 1) an offset-only model, consisting solely of the cell-specific and gene-specific intercept terms (*l_i_* and *θ_j_*_,*k*_); and 2) the full scConvexNMF model, which additionally includes the low-rank component **WH**. Because the intercept terms alone capture substantial variation, we quantified the added explanatory power of the gene-program structure by comparing the Poisson log-likelihood of the full model to that of the offset-only model on held-out entries.

To assess biological interpretability, we evaluated the enrichment of each gene program in a combined pathway annotation matrix that included MSigDB Hallmark gene sets^40^, a HIF core gene signature^41^, a HIF2α signature^42^, HIF-derived gene signatures and cancer cell states gene sets^46^. This analysis initially identified 60 gene programs for malignant cells and 40 for macrophages; one program in each case was subsequently excluded due to enrichment for markers specific to other components of the tumor microenvironment.

Enrichment between each gene program and a given gene set was computed as the product of the Weighted Jaccard Index and a -log_10_ empirical permutation-based p-value of association between gene program loadings and gene sets. For testing significance of association, p-values were calculated by fitting a Gamma distribution parameterized using permutation-derived null distributions of weighted Jaccard similarities. For each rank, we then counted the number of pathways with at least one significantly enriched gene program.

### TCGA Deconvolution Analysis

Cell type deconvolution of bulk RCC samples from the TCGA database was performed with a guided topic model (GTM) implemented in the SpaTM^91^(v.0.3) package. The model was trained using our RCC single cell atlas which was filtered to only include cells from primary RCC samples and cell types represented by at least 1000 cells. The malignant tumor cell population was also sub-sampled to match the distribution of other cell types. For training, we restricted the feature set to genes expressed in at least 2% and less than 75% of cells. These thresholds were determined using leave-one-out cross-validation, based on cell type prediction accuracy and pseudo-bulk deconvolution performance on held-out samples. We then applied the trained GTM on the bulk RCC data to infer cell type proportions. Finally, we validated the deconvolution by examining the correlation between the inferred cell type proportions and the expression of established cell type markers.

### Survival analysis

Survival analyses were conducted using the R packages survival (v.3.5-7) and survminer (0.5.0). Cox proportional hazards regression models were fitted using a binary predictor, incorporating tumor purity and sex as covariates. For analyses involving normal cells, a similar Cox model was applied, substituting tumor purity with the log_10_-transformed estimated cell proportion derived from the deconvolution analysis. To determine optimal cutpoints for continuous variables, we evaluated thresholds from the 15^th^ to the 85^th^ percentiles in 5-percentile increments, classifying samples into ‘high’ or ‘low’ groups at each threshold. All statistical tests across these binnings were pooled, and p-values were adjusted for multiple testing using the Benjamini–Hochberg FDR procedure. For each variable, the most significant association was reported.

### Differential expression analysis

Differential expression analysis was employed to characterize transcriptional differences between primary and metastatic malignant cell populations. To minimize confounding signals from distinct microenvironments, ambient RNA was removed using CellBender^92^ (v.0.3.0). Then, pseudo-bulk expression profiles were generated by aggregating raw counts per sample. Only samples with more than 100 malignant cells were retained for analysis. Differential expression analysis was conducted on the pseudo-bulk profiles using DESeq2^93^ (v.1.42.0). A model comparing primary versus metastatic status was fitted, including sample source (either tumor tissue or PDX) and sex as covariates.

### GSEA analysis

Pseudo-bulk mRNA expression profiles generated during the differential expression step were used as input to GSEA^94^ (v.4.4.0). For macrophage populations, pseudo-bulk profiles were constructed similarly to those of malignant cells, with the exception that only samples with more than 50 macrophage cells were included. GSEA was performed using the gene set permutation mode, with all other parameters set to default. To assess enrichment of gene programs, we used the top 50 genes most strongly associated with each program, as defined by the gene-program association matrix from the scConvexNMF framework.

### Gene set activity estimation

Gene set activity was estimated using the ‘run_ulm’ function from the decoupleR^95^ package (v.2.8.0), applied to log_2_-normalized counts. For the analysis of normal kidney cells, we utilized publicly available data from the Kidney Precision Medicine Project^96^.

### Defining a metastatic transcriptional signature

We used 111 differentially expressed genes (FDR < 0.1) between metastatic and primary tumor malignant ccRCC cells as input features to train a logistic regression model. The model was fitted using the ‘cv.glmnet’ function from the glmnet package (v.4.1.8), specifying ‘family=binomial’ and applying an elastic net penalty with ‘alpha=0.5’. The regularization parameter ‘lambda’ was selected using ‘s=lambda.1se’, corresponding to the most regularized model within one standard error of the minimum cross-validated error. Internal validation was performed using a leave-one-sample-out cross-validation strategy. The model was trained on mean-centered log_2_-normalized single-cell expression values. For prediction on external bulk-RNA seq datasets, log_2_ normalized expression values were obtained using the normalizeCounts from scuttle^75^ and subsequently mean-centered prior to prediction.

### Co-localization framework

For each field of view, we constructed a spatial graph using Delaunay triangulation based on distances between cell centroids, implemented using the ‘tri.mesh’ function from the interp package (v.1.1-5). In the resulting graph, nodes represented cell centroids and edges corresponded to the distances between neighboring cells. To eliminate long-range contacts unlikely to reflect meaningful spatial interactions, we applied a distance threshold of 150 pixels, determined by inspecting the distribution of inter cellular distances. The resulting filtered edges were used to assemble a symmetric adjacency matrix **G**, where nodes correspond to spatial units (cells or spots) and edges encode spatial proximity.

We quantified spatial colocalization between two sets of features **X** and **Y** (e.g., gene programs or binary cell-type indicators) using a graph-based diffusion and weighted correlation framework across multiple neighborhood scales *k*. The adjacency matrix **G**, was normalized to define a column-stochastic transition matrix **P**, enabling signal propagation across the graph. The initial diffusion kernel was configured to emphasize local interactions.

For each diffusion step *k*, the *k*-step transition matrix was applied to smooth **X** and **Y** across the graph. For each kernel center, we computed an effective kernel size based on the inverse squared sum of diffusion weights, along with a coverage statistic describing the effective number of spatial locations sampled by the ensemble of kernels. These quantities were combined to estimate an effective sample size that corrects for spatial dependence introduced by smoothing. Weighted Pearson correlations between features in **X** and **Y** were then computed using kernel-derived weights. Statistical significance was assessed using Fisher’s z-transformation with the estimated effective sample size, and p-values were adjusted for multiple testing using the Benjamini–Hochberg FDR procedure.

Gene program activity in the CosMX datasets was inferred using the scConvexNMF framework. Prior to co-localization analysis, the cell-by-program activity matrices were normalized by applying row-wise mean and variance scaling. Due to the limited number of genes profiled in the CosMX panel (∼1,000 genes), we anticipated that some gene programs might not be adequately represented. To address this, we restricted the analysis to gene programs that had a Pearson correlation > 0.2 with the original model when the single-cell expression data was downsampled to match the CosMX feature set.

For each neighborhood-step *k*, meta-analysis across samples or FOVs was performed using the ‘ma_r’ function from the psychmeta (v.2.7.0) package, incorporating the estimated effective sample size in the ‘n’ parameter. Adjusted p-values were calculated using the Benjamini-Hochberg procedure.

### Immunostaining and immunohistochemistry

Multiplex immunofluorescence was performed on 5 um sections of individual tumors or TMAs containing 1mm cores of primary and metastatic RCC tumors, as well as normal kidney controls. Following antigen retrieval (Tris pH 9 for 15 min at 98C), slides were blocked (4% BSA in PBS) and then incubated with a fluorophore-conjugated primary antibody cocktail targeting CASP14 (Novus Cat# NBP2-90006AF594, RRID:AB_3445730), CK8/18 (Novus Cat# NBP3-11504AF532, RRID:AB_3579423), and CA9 (Novus Cat# NB100-417AF647, RRID:AB_3157478), together with the DNA dye SYTO13 (S7575, Invitrogen). Images were acquired on the GeoMx™ DSP (NanoString Technologies) epi-fluorescent microscope.

### Immunostaining of protein-translation markers

Immunofluorescence staining of p-4EBP1, p-S6, vimentin, and CA9 in FFPE slides from ccRCC patient tumors and the lung tissue of the PID001 PDX were performed by deparaffinization at 60 °C for 30 min, followed by incubation in xylene for 15 min (2 times), 100% ethanol for 5 min (2 times), 70% ethanol for 5 min (1 time), and deionized water for 5 min (twice). Antigen retrieval was done with Tris-EDTA pH=9 at 96 °C for 20 min, followed by cooling down to room temperature for 30 min, and washing with deionized water for 5 min. Blocking was done at room temperature with 5% BSA for 30 min. Slides were incubated with primary antibodies (p-4EBP1 (CST Cat# 2855S, RRID:AB_560835, 1:300), p-S6 (CST Cat# 5364S, RRID:AB_10694233, 1:300), vimentin (CST Cat# 5741S, RRID:AB_10695459, 1:300), CA9 (Abcam Cat# ab15086, RRID:AB_2066533, 1:100) at +4 °C overnight, followed by 3 times washing in PBS for 5 min and incubation with the Alexa fluor-488 conjugated secondary antibody at a dilution of 1:100 for 2 hrs at 37 °C. DAPI was used for nuclear counterstaining. Images were taken from three different areas at 10X magnification using Zeiss LSM700 Confocal Microscopy. Regions of interests were selected at CA9 positive regions of the tumors and the corresponding intensities for p-4EBP1, p-S6, and vimentin on consecutive slides were averaged for each area and plotted for each donor.

### Generation of CosMx data

CosMx™ Spatial Molecular Imaging (SMI) was performed as per the supplier’s protocol using the Universal Cell Characterization RNA Panel (Nanostring Technologies). Briefly, 5 µm FFPE sections of ccRCC tumors were processed through deparaffinization, antigen retrieval (in Tris-EDTA ph9 for 15 min at 98°C), and Proteinase K digestion (3 ug/mL Proteinase K for 30 min at 40°C) prior to overnight incubation at 37°C with the human Universal Cell Characterization RNA Panel diluted in hybridization Buffer R. The next day, slides were washed twice in 2XSSC/formamide (25 min at 37°C), twice in 2XSSC, and incubated with a segmentation antibody cocktail targeting CD298, B2M, PanCK, CK8/18, CD45, and SYTO13 for 1 hr in a humidity chamber. Slides were washed 3 times in PBS and then loaded onto flowcells for cyclic imaging of defined FOVs (0.51 x 0.51 mm) on the CosMx™ Spatial Molecular Imager, resulting in single molecule read-out of detected mRNAs. Quality control, cell segmentation, and RNA normalization was performed on AtoMx™ prior to downstream analysis.

### Pre-processing of CosMx data

The count matrix from the MTSCRCC tumor was loaded into a Seurat object, filtering out cells with fewer than 10 detected features and genes expressed in fewer than 3 cells, and log-normalized using Seurat’s NormalizeData function with a scaling factor of 10,000. The data were then scaled using the ScaleData function. The top 40 principal components were selected based on elbow plots and JackStraw analysis and were used to construct a k-nearest neighbor graph and perform graph-based clustering using the Louvain algorithm at a resolution of 0.5 (FindNeighbors/FindClusters). The resulting shared nearest neighbor (SNN) clusters were characterized by cluster-specific marker genes identified using the FindAllMarkers function. Cells without distinct gene expression profiles, likely due to limited transcript coverage, were labeled as “Undefined”, while other clusters were annotated based on canonical marker genes identified in our CosMx 1K spatial transcriptomics dataset.

For the analysis of a previously published single-cell-resolution spatial transcriptomics dataset of ccRCC tumors^44^, CosMx spatial transcriptomics data were obtained from the original publication repository (Zenodo DOI: 10.5281/zenodo.12730226). Raw gene expression counts, metadata and spatial coordinates were extracted from the provided Seurat object. For downstream analyses, we included only cells and fields of view that passed the quality control criteria defined in the original study. The original cell type annotation from the authors was used with the following modifications for defining malignant cells. First, to refine the identification of malignant cells, we integrated the CosMx count data with our single-cell RNA-seq data using GEDI. Logistic regression models were trained on the single-cell data to predict the cell labels from coordinates in the integrated space, and where used to imputed cell type modality for the CosMx data set. Subsequently, we performed clustering on the normalized log-transformed CosMx expression data using PCA followed by Leiden clustering. For each cluster, we computed the mean cell type label modality for the malignant cell type. Clusters with a mean malignant cell type modality exceeding a threshold of 0.06 were designated as malignant. Finally, cells were labelled as malignant in our analysis only if they were annotated as malignant in the original metadata and belonged to one of the identified malignant clusters. This procedure was conducted separately for cells originating from tumor and stroma fields of view.

## Data availability

Processed single-cell gene expression data from RCC samples, raw and processed single cell spatial transcriptomics from a MTSCRCC sample, and raw and processed bulk RNA-seq gene expression data from A498 ccRCC cells are available in the Gene Expression Omnibus (GEO) under accession number GSE328692. Other datasets used in the article were downloaded from the publication access codes or from their online data repositories. Preprocessed data, embeddings, and models can be accessed via Zenodo (DOIs: 10.5281/zenodo.20058906 and 10.5281/zenodo.20086236). A description of the Zenodo files, including additional source data files, is available in **Supplementary Table 1**.

## Code availability

scConvexNMF is available via GitHub at https://github.com/csglab/scConvexNMF. Reproducible notebooks for the analyses presented in this work can be found at https://github.com/csglab/RCC_manuscript.

## Acknowledgements

This work was supported by research grants from the Canadian Institutes of Health Research (CIHR) [PJT-173317 to HSN], New Frontiers in Research Fund [NFRFE-2019-00975 to HSN and YR], Cancer Research Society and the Canadian VHL Alliance [23479 to HSN], the United States Department of Defense (DoD) Kidney Cancer Research Program (KCRP) [W81XWH2110945 and W81XWH-2110579 to YR], and in part by research funding from CIHR to PMS [CIHR PJT-175066], in addition to resources provided by Calcul Québec (calculquebec.ca) and the Digital Research Alliance of Canada (alliancecan.ca) to HSN. AM and ZM were supported by doctoral training awards from Fonds de Recherche du Québec Santé, and MK was a recipient of a doctoral scholarship from ASAN Foundation. HSN holds a CIHR Canada Research Chair.

## Author contributions

Conceptualization: A Madrigal, MK, YR, HSN. Methodology: A Madrigal, MK, ZM, TN, O Saatci, EM, KIG, HK, TL, JR, O Sahin, FB, ST, YR, HSN. Software: A Madrigal, HSN. Formal analysis: A Madrigal, MK, AO, EZ. Visualization: A Madrigal, MK, HSN. Investigation: A Madrigal, MK, ZM, TN, O Saatci, MD, SMM, HK, VP, A Monast, MS, MA, SO, AH, RR, LMS. Resources: JDS, KP, PS, MP, JR, O Sahin, FB, ST, YR, HSN. Data Curation: A Madrigal, MK, AHC. Writing - Original Draft: A Madrigal, MK, YR, HSN. Writing - Review and Editing: A Madrigal, MK, YR, HSN, with contributions from all authors. Supervision: YR, HSN. Project administration: YR, HSN. Funding acquisition: YR, HSN.

## List of Supplementary Figures and Tables

**Supplementary Figure 1.** Overall cellular composition across RCC tumors.

**Supplementary Figure 2.** CNV patterns in a ccRCC sample.

**Supplementary Figure 3.** Comparison between CNV patterns from bulk genomic analysis and single-cell inferred CNV profiles.

**Supplementary Figure 4.** Subtype gene-signature activities across pathological subtypes in TCGA.

**Supplementary Figure 5.** *TFE3* fusion events detected in TCGA samples.

**Supplementary Figure 6.** ccpRCT subtype markers.

**Supplementary Figure 7.** Additional examples comparing intra-tumor clustering and clonal heterogeneity.

**Supplementary Figure 8.** Schematic of the scConvexNMF model.

**Supplementary Figure 9.** Metrics used to select the rank *P* for the scConvexNMF model.

**Supplementary Figure 10.** Representative example of gene program identification in samples harboring CNVs.

**Supplementary Figure 11.** Additional associations of malignant gene programs with biological gene sets.

**Supplementary Figure 12.** Additional examples of pathways from the Hallmark MsigDB collection deconvolved into submodules.

**Supplementary Figure 13.** Association of hypoxia gene programs with regulons from DoRothEA.

**Supplementary Figure 14.** HIF-specific siRNA-derived gene signatures.

**Supplementary Figure 15.** Additional spatial analysis of malignant cells.

**Supplementary Figure 16.** Correlation of predicted program activity and observed counts.

**Supplementary Figure 17.** Subtype-specific gene programs across molecular subtypes in TCGA.

**Supplementary Figure 18.** Disease-free survival analysis of gene programs in TCGA.

**Supplementary Figure 19.** Additional examples of gene programs associated with survival outcomes in TCGA.

**Supplementary Figure 20.** Differential expression analysis between primary and metastatic ccRCCs.

**Supplementary Figure 21.** Additional gene programs differentially active between metastatic or primary ccRCC.

**Supplementary Figure 22.** Activity of primary-associated gene programs in an atlas of kidney cells.

**Supplementary Figure 23.** Internal validation of the metastatic classifier.

**Supplementary Figure 24.** External validation of the metastatic classifier using data from TRACERx.

**Supplementary Figure 25.** Distant metastasis of a primary tumor-derived PDX.

**Supplementary Figure 26.** Cellular origins of RCC subtypes.

**Supplementary Figure 27.** Macrophage gene program analysis.

**Supplementary Figure 28.** Macrophage gene programs differentially enriched between metastatic and primary ccRCC

**Supplementary Figure 29.** Spatial analysis of macrophage gene programs.

**Supplementary Table 1.** List of source data files.

**Supplementary Data Table 1.** Specimens from the RCC cohort.

**Supplementary Data Table 2.** Transcriptomic reclassification of RCC tumors from TCGA.

**Supplementary Data Table 3.** Top 50 genes per program across 59 malignant gene programs.

**Supplementary Data Table 4.** Differentially expressed genes between metastatic and primary ccRCC.

**Supplementary Data Table 5.** Top 50 genes per program across 39 macrophage gene programs.

**Supplementary Data Table 6.** Marker genes used for cell type annotation.

**Supplementary Data Table 7.** Gene set used to minimize the impact of stroma or immune-specific gene expression in unsupervised clustering of TCGA bulk RNA-seq samples.

**Supplementary Data Table 8.** Marker genes defining RCC subtype signatures.

