## Supplementary Figures for "Single-cell gene programs define subtype identity and metastatic trajectories in renal cell carcinoma"

### Supplementary Figures and Tables

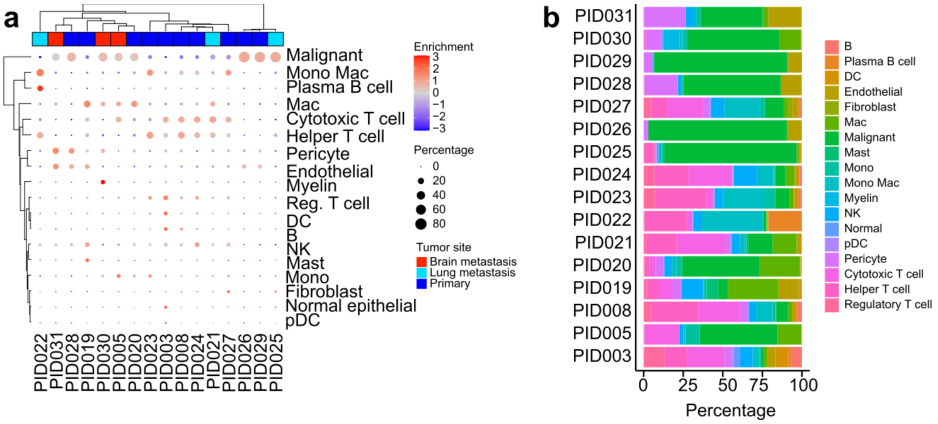

**Supplementary Figure 1.** Overall cellular composition across RCC tumors

(**a**) Dot plot depicting the overall cellular composition of major cell types across RCC tumors. Dot size corresponds to the percentage of each cell type within a given donor, and dot color indicates the relative enrichment of that cell type. For four samples (PID026, PID028, PID029 and PID031) $CD45^+$ cells were depleted during tumor processing prior to generating the single-cell library. Top color bars indicate the tumor site for each sample. (**b**) Bar plots showing the cellular composition of each tumor, with colors representing distinct cell types.

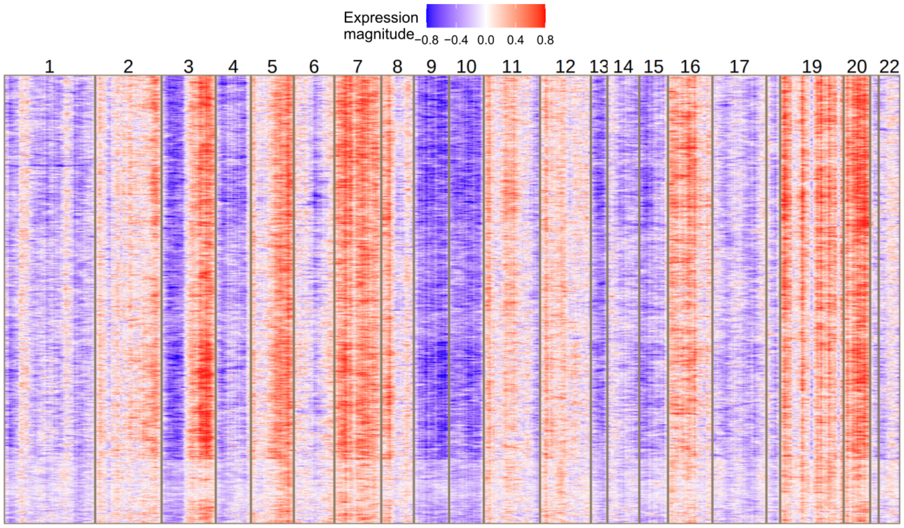

**Supplementary Figure 2.** CNV patterns in a ccRCC sample

Heatmap showing single-cell smoothed expression profiles for malignant cells from sample PID001. The plot reveals a pronounced downregulation of genes located on chr3p.

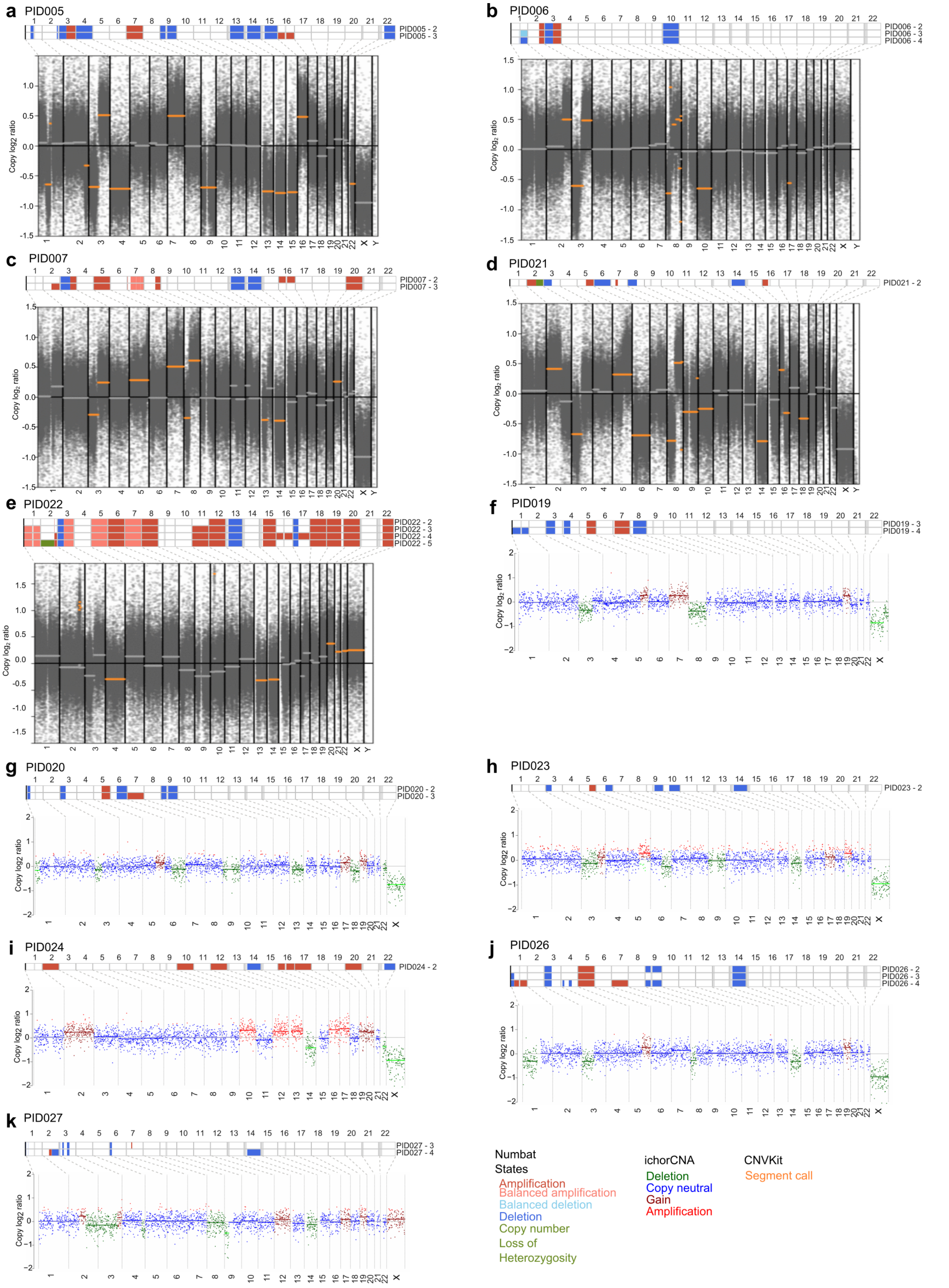

**Supplementary Figure 3.** Comparison between CNV patterns from bulk genomic analysis and single-cell inferred CNV profiles (Figure caption continued on the next page).

CNV patterns derived from whole-exome sequencing or an RCC-specific 24 gene panel[1] sequencing are compared to single-cell inferred CNV patterns from Numbat. For each sample, the top heatmap shows the Numbat analysis, where each row represents a subclone, with colors denoting CNV states identified by Numbat[2]. Gray vertical bars indicate gap regions. The bottom heatmap shows the inferred $log_2$ ratio status from bulk genomic analysis, either from whole-exome sequencing (a-e), inferred with CNVKit[3], or from the RCC-specific gene panel (f-k), inferred with ichorCNA[4]. Dotted lines link corresponding chromosomes between the two methods. Samples shown: (**a**) PID005, (**b**) PID006, (**c**) PID007, (**d**) PID021, (**e**) PID022, (**f**) PID019, (**g**) PID020, (**h**) PID023, (**i**) PID024, (**j**) PID026, (**k**) PID027.

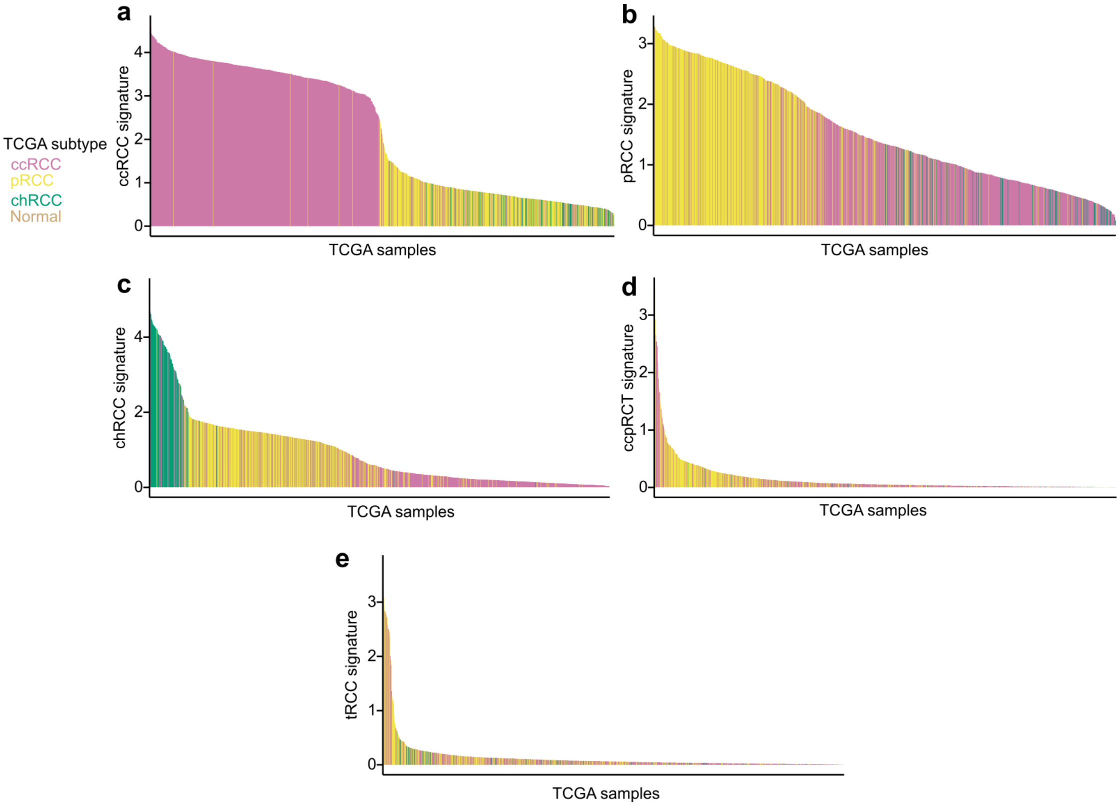

**Supplementary Figure 4.** Subtype gene-signature activities across pathological subtypes in TCGA

Bar plots showing single-cell derived subtype gene-signature activities in TCGA samples (ccRCC, pRCC, chRCC and normal). Each bar represents one bulk RNA-seq sample, with colors indicating the original pathological subtype annotated in TCGA. Gene signatures shown are: (**a**) ccRCC, (**b**) pRCC, (**c**) chRCC, (**d**) ccpRCT and (**e**) tRCC.

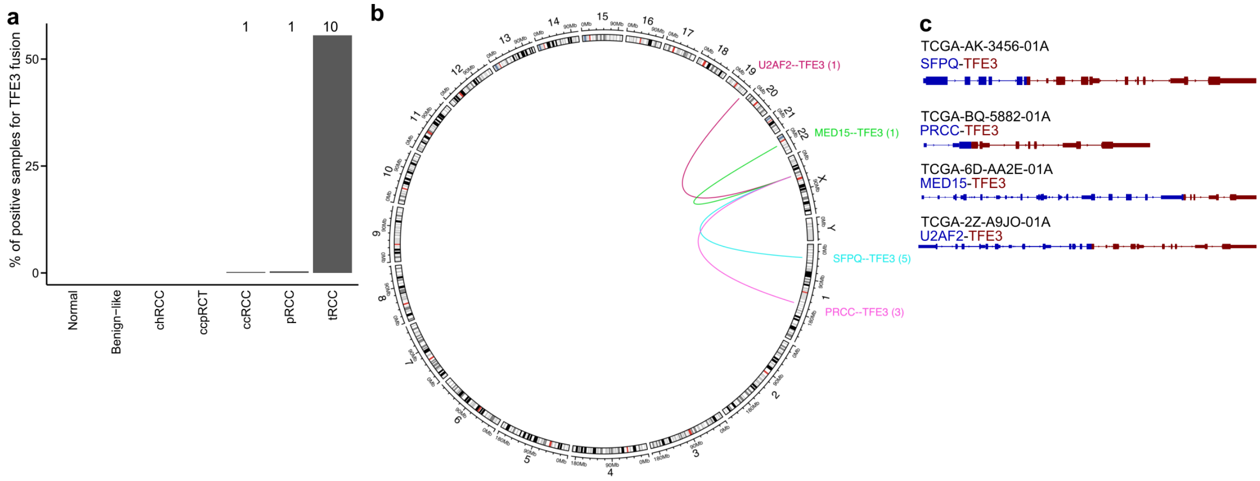

**Supplementary Figure 5.** *TFE3* fusion events detected in TCGA samples

(**a**) Bar plot showing the percentage of samples positive for a *TFE3* fusion event across TCGA molecular subtypes. The number of samples in each subtype is indicated above bar. Fusion events were detected using StarFusion (see **Methods**). Only in-frame predicted fusions are included in this analysis. (**b**) Circos plot depicting *TFE3*-associated fusion events. For each fusion pair, the number of samples harboring that event is indicated. (**c**) Representative examples of *TFE3* fusion events. The corresponding TCGA sample identifiers are shown.

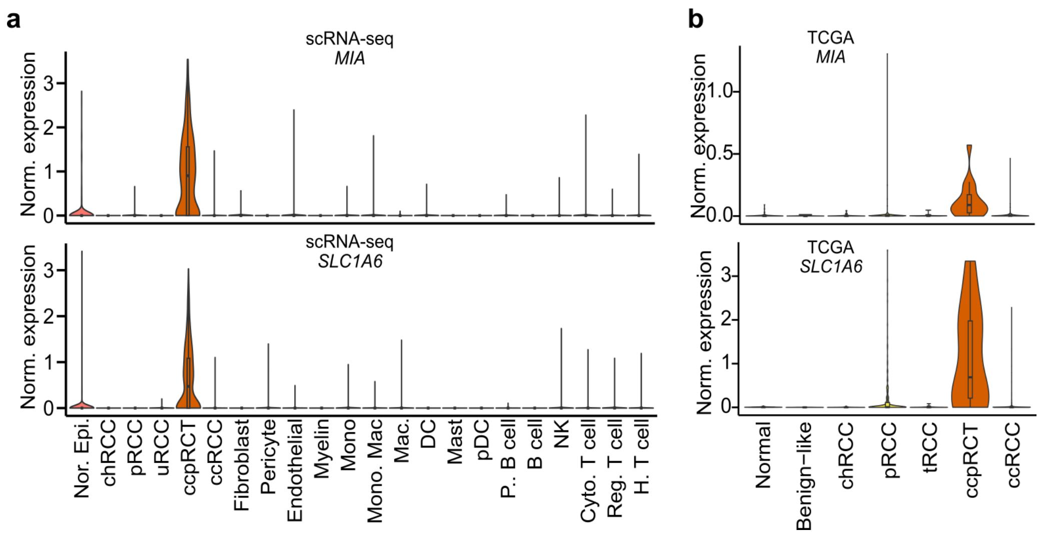

**Supplementary Figure 6.** ccpRCT subtype markers

Violin plots showing normalized expression of *MIA* and *SLC1A6*, which, together with *CASP14* (see Fig. 3e,f), serve as markers of the ccpRCT subtype. (**a**) Violin plots showing expression across cell types in RCC tumors. (**b**) Violin plots showing expression across molecular subtypes derived from bulk RNA-seq profiles of RCC tumors from TCGA.

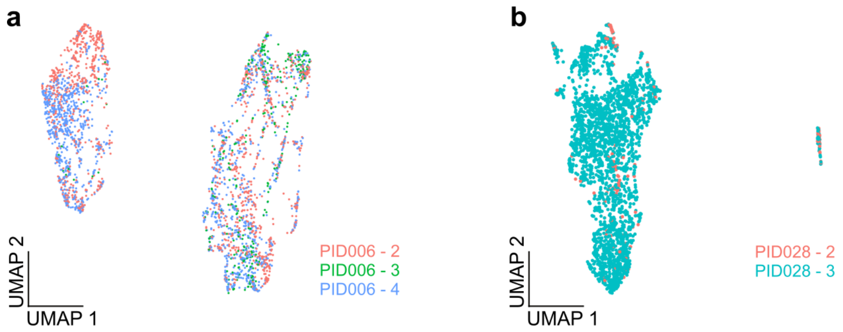

**Supplementary Figure 7.** Additional examples comparing intra-tumor clustering and clonal heterogeneity

(**a-b**) UMAP embeddings computed from the log-normalized expression counts for each sample, with colors indicating clonal identity. (**a**) PID006. (**b**) PID028.

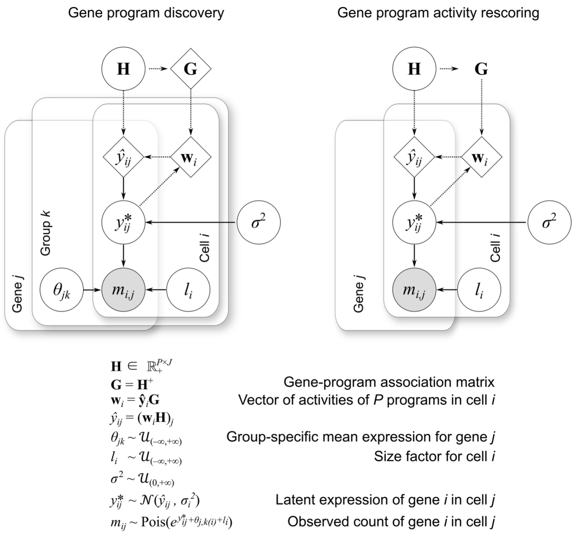

**Supplementary Figure 8.** Schematic of the scConvexNMF model

Diagram of the probabilistic generative model implemented in scConvexNMF. The left panel illustrates how the model is fitted while accounting for group-specific mean effects $\boldsymbol{\theta_{j,k}}$ during gene program discovery. The right panel shows the estimation of gene program activities for a new dataset, where G is held fixed and only overall gene-specific means $\boldsymbol{\theta_j}$ are removed. Circles and diamonds represent random and deterministic variables, respectively; grey shading indicates observed variables.

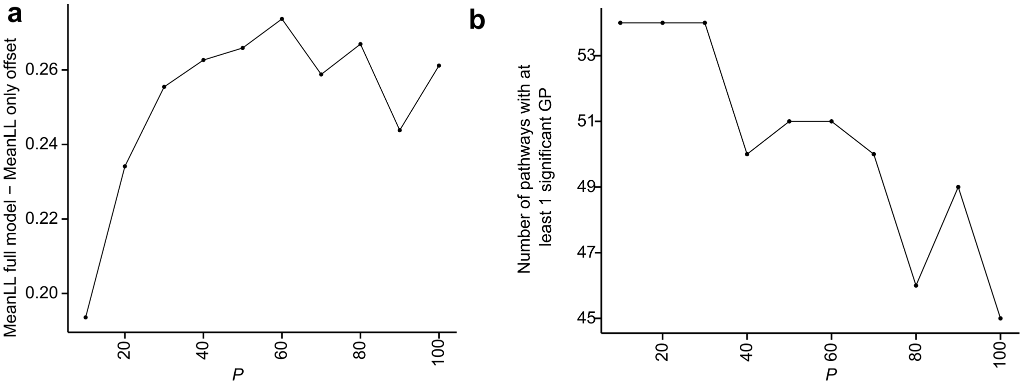

**Supplementary Figure 9.** Metrics used to select the rank *P* for the scConvexNMF model

(**a**) Added explanatory power of the gene-program structure, quantified by comparing the Poisson log-likelihood of the full model on held-out entries (see **Methods**) to that of an offset-only model containing only cell-specific and gene-specific intercept terms. (**b**) Interpretability of the model, measured by counting the number of pathways for which at least one gene program is significantly enriched (see **Methods**). The pathway matrix included MSigDB Hallmark gene sets[5], a HIF core gene signature[6], a HIF2α signature[7], HIF-derived gene signatures and cancer cell states gene sets[8]. Based on these two metrics, we selected the rank *P*=60.

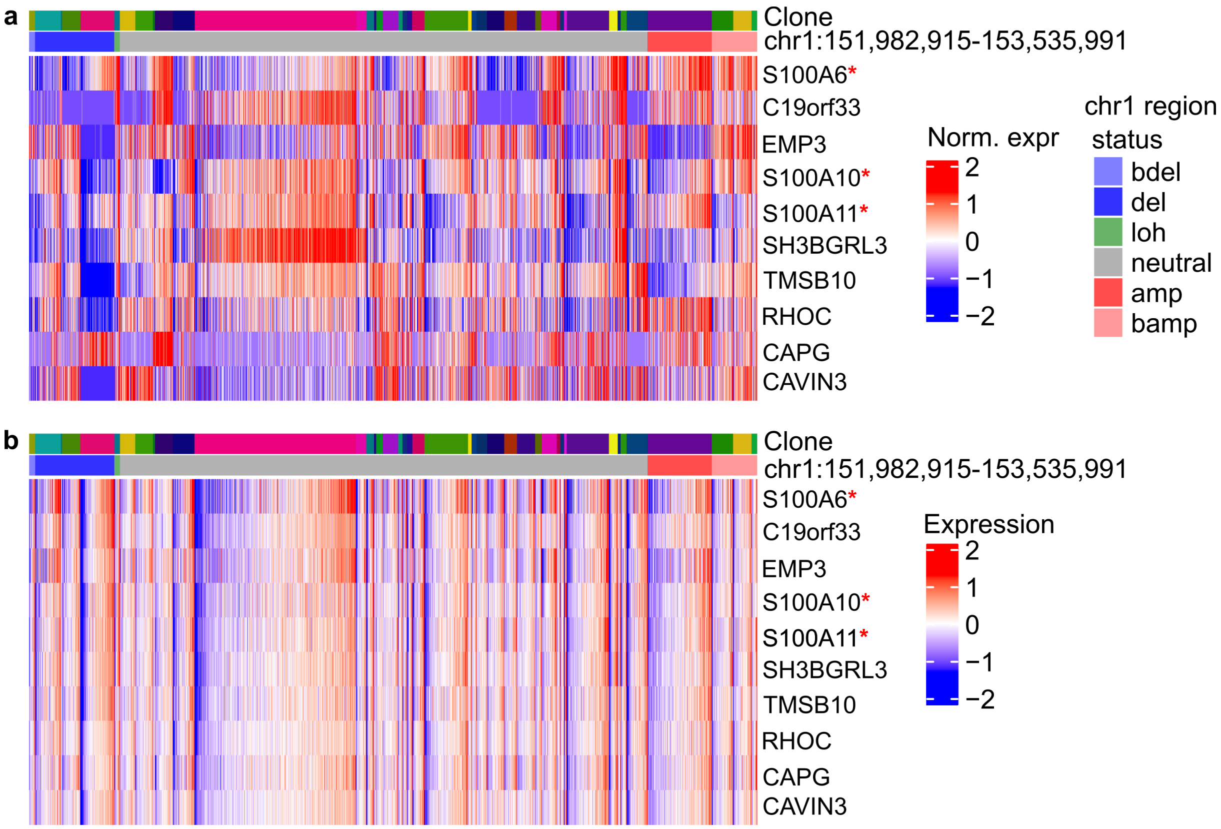

**Supplementary Figure 10.** Representative example of gene program identification in samples harboring CNVs

(**a**) Heatmap showing the normalized latent-log expression from the scConvexNMF framework, obtained after fitting the model to single-cell data and subsequently rescoring it. In this version of the model, only overall gene-specific mean effects were removed, thereby preserving sample-specific signals that reflect CNV patterns. The heatmap displays the top 10 member genes of GP59; among these, *S100A6*, *S100A10* and *S100A11* lie in the indicated chr1 region. The top annotation bar indicates clonal identity and chr1 region status inferred by Numbat[2]. (**b**) Same as in (a) but showing latent-log expression from the scConvexNMF framework after the initial fitting step, incorporating group specific information, thereby removing both intra-clone and inter-sample specific effects.

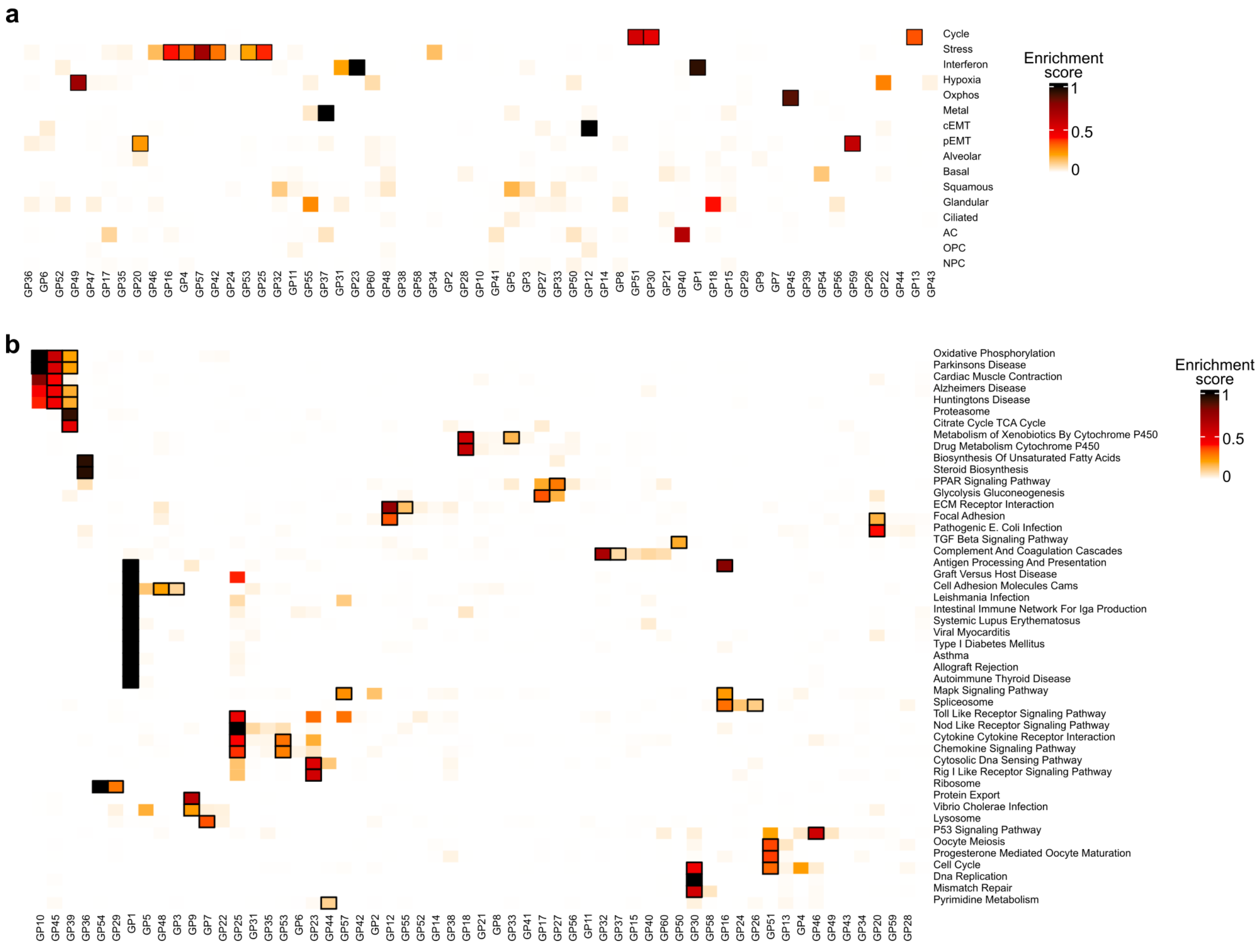

**Supplementary Figure 11.** Additional associations of malignant gene programs with biological gene sets

(**a**) Association between gene programs (columns) and recurrent cancer-cell states[8] gene sets (rows). Heatmap shows enrichment scores; significant associations (FDR < 0.05) are outlined with a black box. (**b**) Same as in (a) but showing association between gene programs (columns) and KEGG[9] legacy gene sets (rows). Due to the large number of gene sets in this collection, only those with a significant overlap (FDR < 0.05) with at least one gene program are shown.

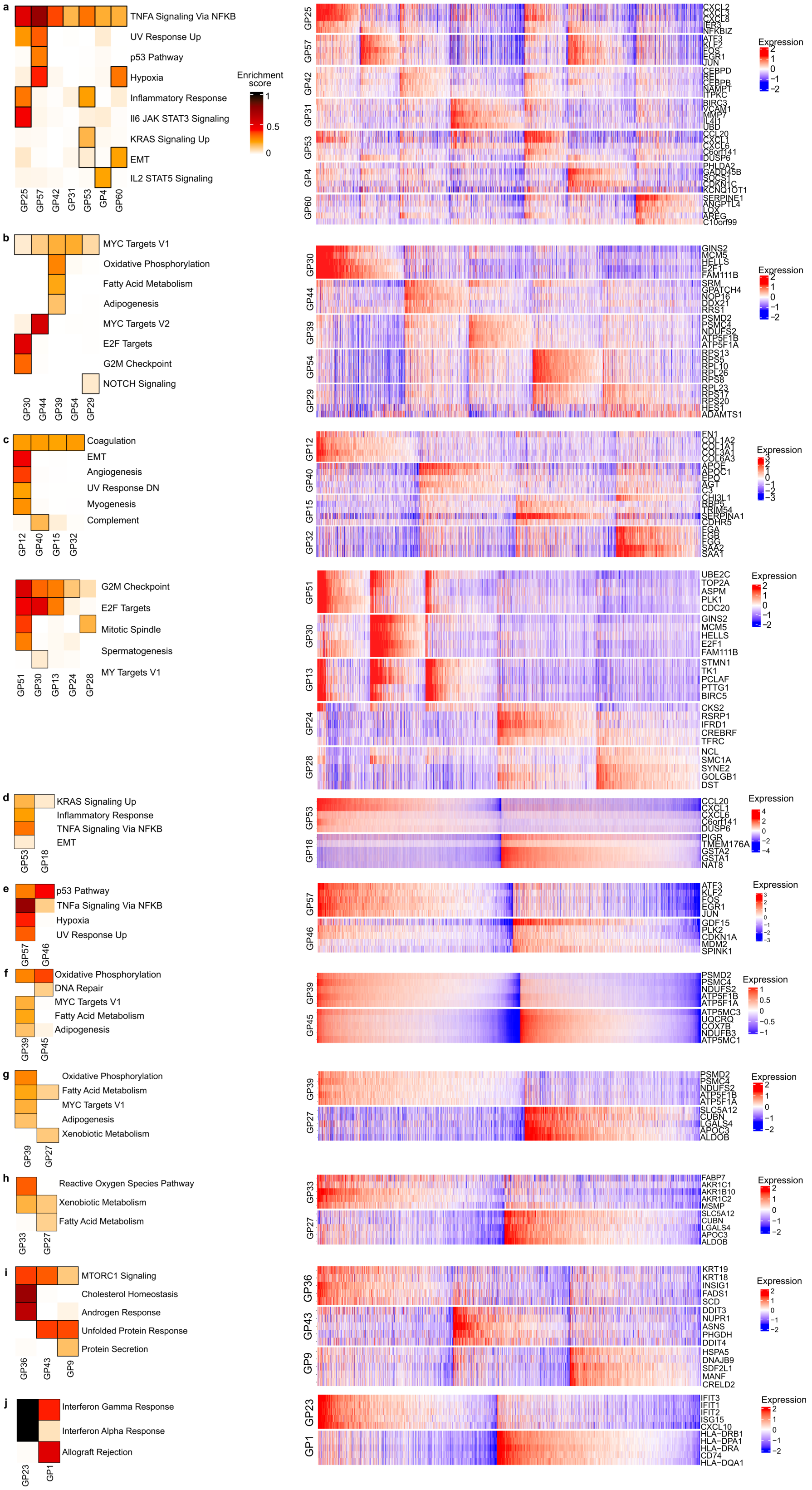

**Supplementary Figure 12.** Additional examples of pathways from the Hallmark MsigDB collection deconvolved into submodules (Figure caption continued on the next page).

For each pathway, the left panel shows a heatmap of associations between gene programs (columns) and Hallmark pathways (rows). Heatmap shows enrichment scores; significant associations (FDR < 0.05) are outlined with a black box. The right panel shows a heatmap of normalized latent-log expression from the scConvexNMF framework, displaying the top five member genes for each gene program across individual cells. The Hallmark pathways shown are: (**a**) TNFa signaling via NFKB, (**b**) Myc targets, (**c**) Coagulation, (**d**) Cell cycle, (**e**) p53 pathway, (**f**) Oxidative phosphorylation, (**g**) Fatty acid metabolism, (**h**) Xenobiotic metabolism, (**i**) mTORC1 signaling, (**j**) Interferon response.

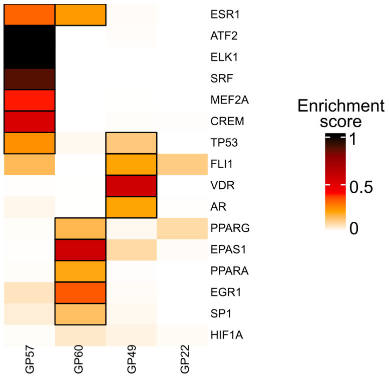

**Supplementary Figure 13.** Association of hypoxia gene programs with regulons from DoRothEA

Association between gene programs (columns) and DoRothEA[10] regulons (rows). DoRothEA regulons were filtered to retain only the highest-confidence TF-target interactions (confidence levels A,B,C). Heatmap shows enrichment scores; significant associations (FDR < 0.1) are outlined with a black box.

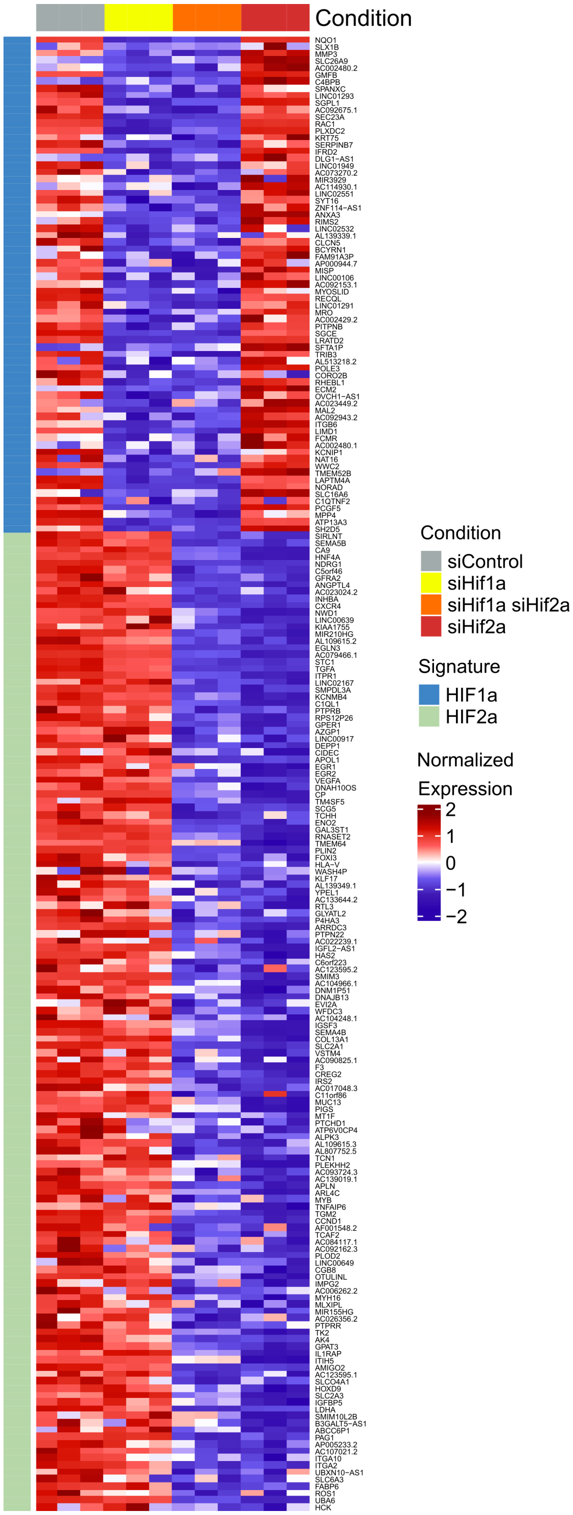

**Supplementary Figure 14.** HIF-specific siRNA-derived gene signatures

Activity signatures for HIF1A and HIF2A were derived by performing siRNA-mediated knockdown of *HIF1A* and *EPAS1* (see **Methods**). RNA-seq libraries were generated and sequenced across four experimental conditions: control, HIF1A-KD, EPAS1-KD, and double-KD, each with three biological replicates. The heatmap displays row-wise z-scores calculated from $\log_2$ normalized gene expression counts, showing the HIF1A- and HIF2A-derived gene set signatures in the left bar. The top annotation bar indicates the experimental condition for each sample.

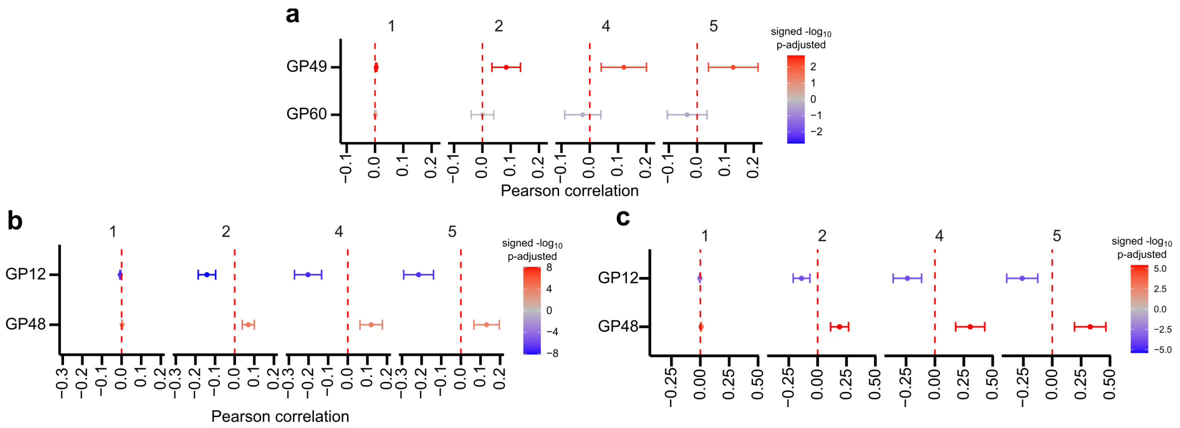

**Supplementary Figure 15.** Additional spatial analysis of malignant cells

(**a**) Meta-analysis of weighted correlation between local malignant cell density and activities of GP49 and GP60 in a single-cell spatial transcriptomic dataset[11] of ccRCC tumors. Pearson correlation coefficients and 95% confidence intervals were derived via meta-analysis across FOVs. Color represents signed -$\log_{10}$ FDR values. Each panel displays a different neighborhood-step *k* (see Figure 4j for *k* = 3). (**b**) Same as in (a), but assessing the relationship between local malignant cell density and activities of GP12 and GP48. (**c**) Same as in (b), but performing weighted correlation analysis using a single-cell resolution spatial transcriptomic map of an MTSCRCC tumor.

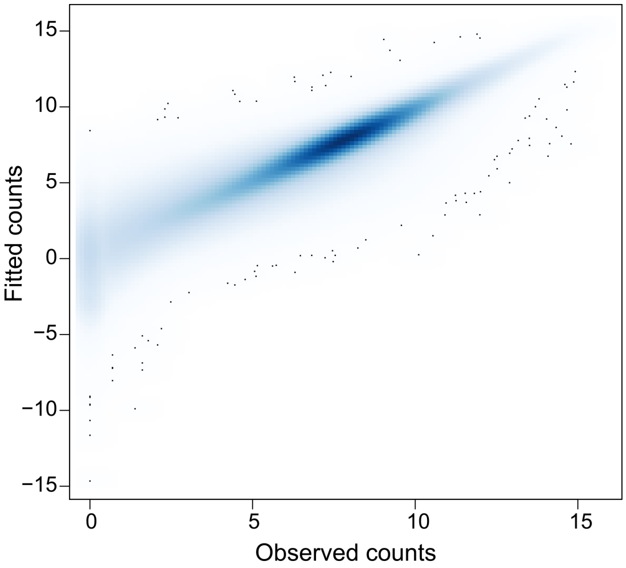

**Supplementary Figure 16.** Correlation between predicted gene expression based on program activity and observed gene expression counts from TCGA

Scatterplot comparing the fitted counts from the scConvexNMF model to original observed gene expression counts from TCGA. Pearson correlation *r* is indicated.

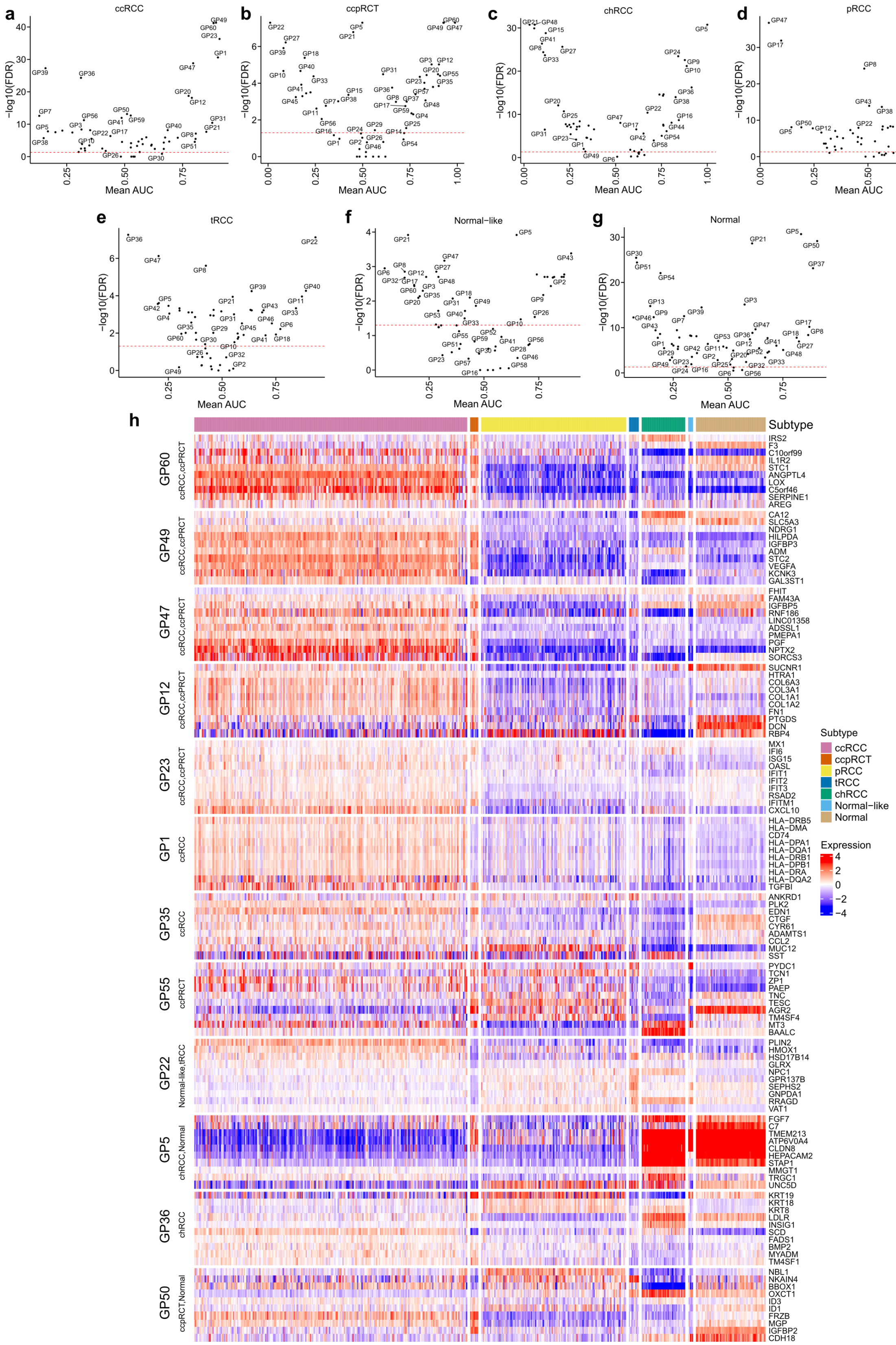

**Supplementary Figure 17.** Subtype-specific gene programs across molecular subtypes in TCGA (Figure caption continued on the next page).

(**a**-**g**) Volcano plots showing differentially active programs across RCC molecular subtypes and normal samples. For each group (either molecular subtype or normal samples), group-specific gene programs were identified using marker analysis. Mean AUC statistics were computed using the scoreMarkers function from scran[12], while p-values and FDR values were obtained with findMarkers. The red dotted line indicates FDR = 0.05. Molecular subtypes shown are: (**a**) ccRCC, (**b**) ccpRCT, (**c**) chRCC, (**d**) pRCC, (**e**) tRCC, (**f**) Normal-like and (**g**) normal. (**h**) Subtype-specific programs in TCGA. Heatmap shows normalized latent-log expression values derived using the scConvexNMF framework. Only programs with a mean AUC > 0.9 are shown. Rows display the top 10 member genes for each program; columns represents individual RNA-seq samples. Top bar indicates molecular subtype. Subtype labels for gene programs were determined by identifying subtypes in which the mean AUC was greater than 0.7.

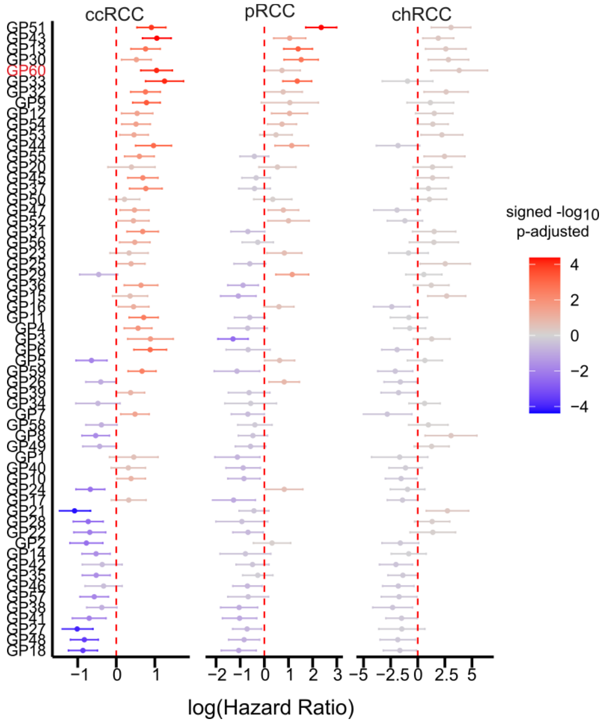

**Supplementary Figure 18.** Disease-free survival analysis of gene programs in TCGA

Disease-free survival analysis of RCC gene programs across the three major RCC subtypes in TCGA. Plots show log hazard ratios with 95% confidence intervals from a Cox-regression model that included purity and sex as covariates. Color represents signed -$\log_{10}$ FDR values.

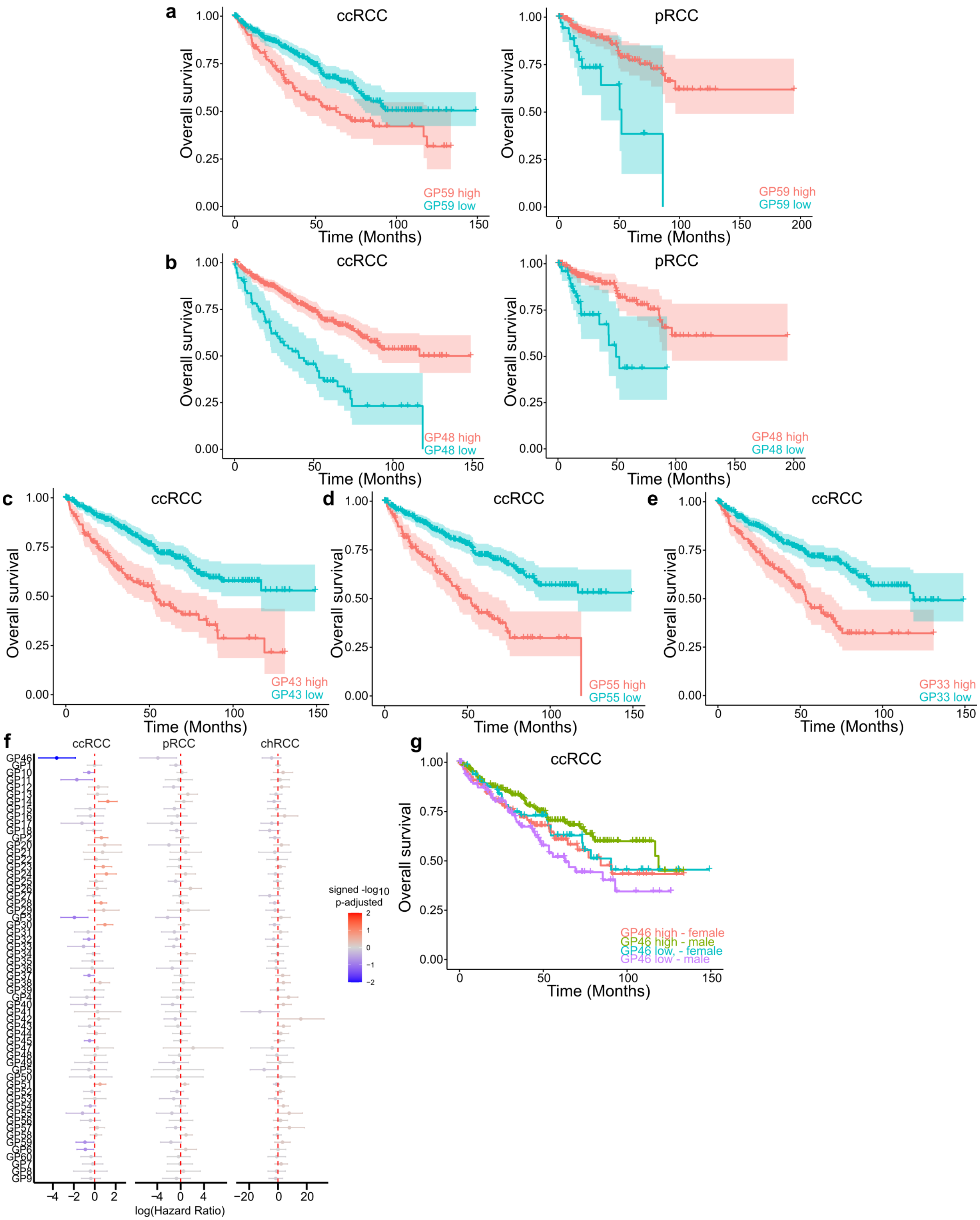

**Supplementary Figure 19.** Additional examples of gene programs associated with survival outcomes in TCGA

(**a**) Left: Kaplan-Meier overall survival curve comparing ccRCC tumors with high versus low GP59 activity in TCGA. Patients were stratified into two groups based on the optimal cutoff determined during testing (see **Methods**). Right: Kaplan-Meier overall survival curve for pRCC tumors. (**b**) Left: Kaplan-Meier overall survival curve comparing ccRCC tumors with high versus low GP48 activity. Right: Kaplan-Meier overall survival curve for pRCC tumors (**c**-**e**) Kaplan-Meier survival curves for gene programs strongly associated with worse overall survival in ccRCC tumors, including (**c**) GP43, (**d**) GP55 and (**e**) GP33. (**f**) Overall survival analysis of RCC gene programs whose association with outcome depends on sex across the three major RCC subtypes in TCGA. Cox-regression models included an interaction term between each gene program stratified variable and sex, with purity and sex as covariates. Plots display log hazard ratios with 95% confidence intervals. Color represents signed $-\log_{10}$ FDR values. (**g**) Kaplan-Meier survival curve comparing overall survival of ccRCC tumors with high versus low GP46 activity stratified by sex.

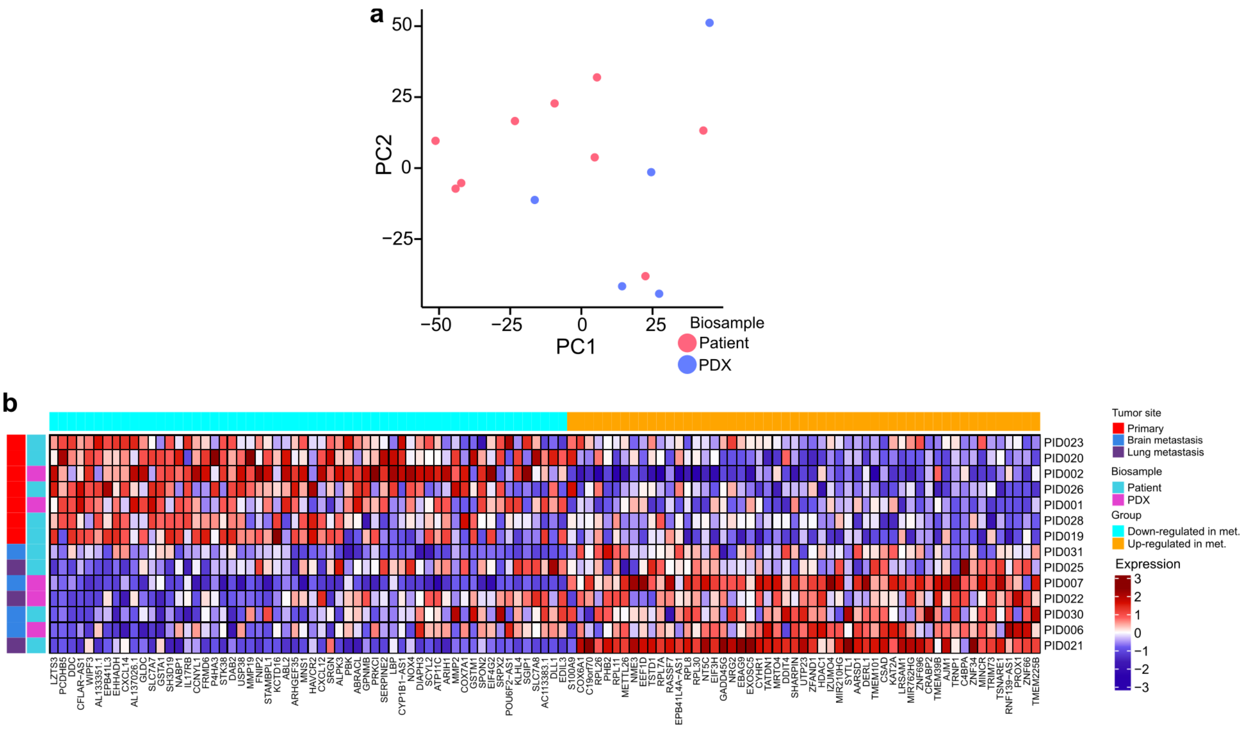

**Supplementary Figure 20.** Differential expression analysis between primary and metastatic ccRCCs

(**a**) PCA of pseudo-bulk mRNA profiles from malignant ccRCC cells. Each dot represents one sample; color indicates biosample type. (**b**) Heatmap of differentially expressed genes between primary and metastatic ccRCC samples. Values represent column-wise z-scores of $log_2$-normalized pseudo-bulk RNA-seq expression profiles for 111 genes (FDR < 0.05). Left annotation bar indicates metastatic status and bio type for each sample; top annotation bar indicates whether the gene was up- or downregulated.

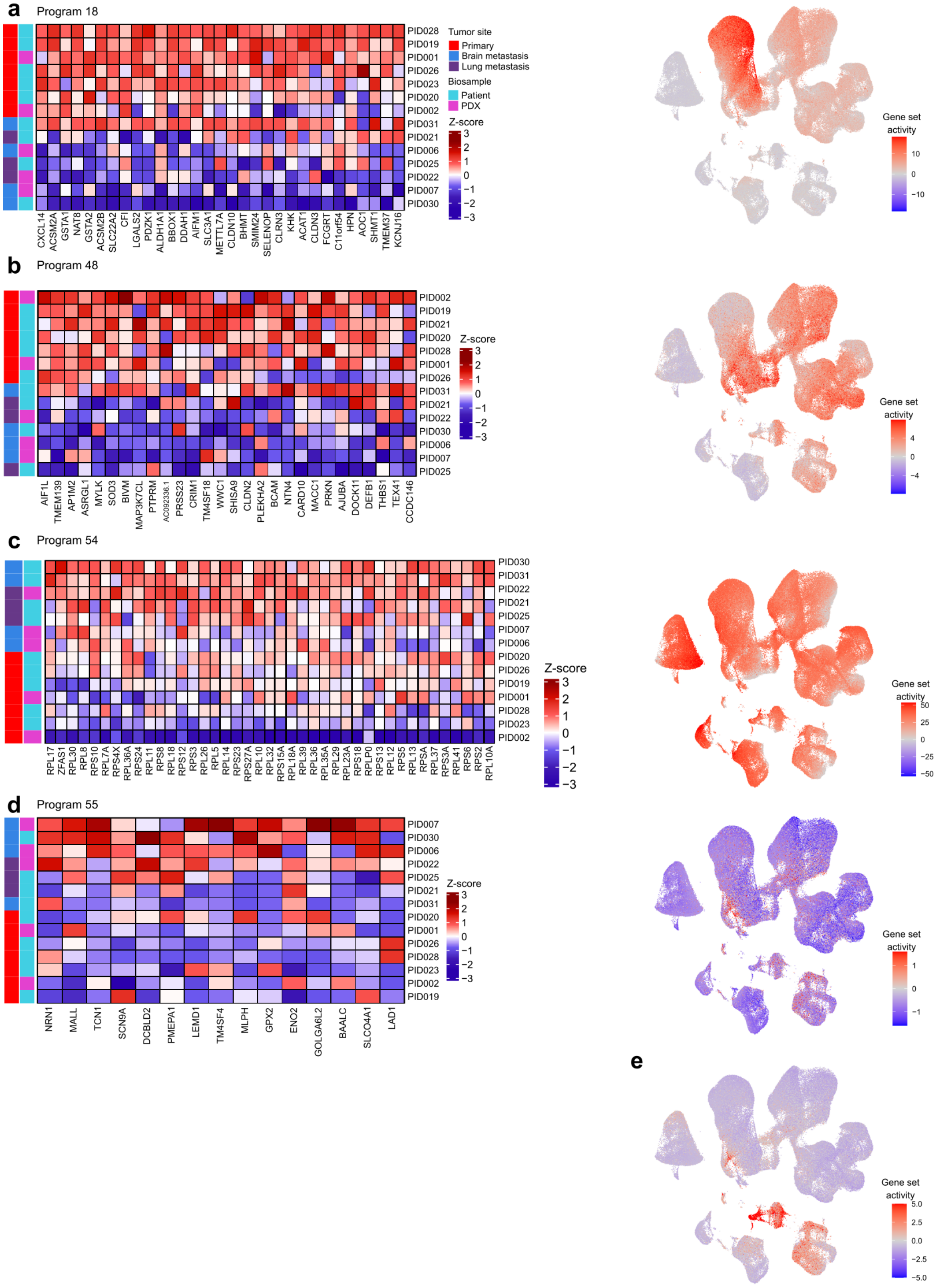

**Supplementary Figure 21.** Additional gene programs differentially active between metastatic or primary ccRCC

For each gene program, the left panel shows a heatmap of leading-edge genes, displaying column-wise z-scores of $\log_2$-normalized pseudo-bulk RNA-seq expression profiles from ccRCC samples. The left annotation bar indicates metastatic status and biosample type for each sample. The right panel shows the activity of leading-edge genes for each gene program in an atlas of single-cell expression profiles of kidney cells from the Kidney Precision Medicine Project[13]. UMAP embedding shows gene-set activity estimated using the ulm method from decoupleR[14]. The gene programs shown are: (**a**) GP18, (**b**) GP48, (**c**) GP54, (**d**) GP55 and (**e**) GP12.

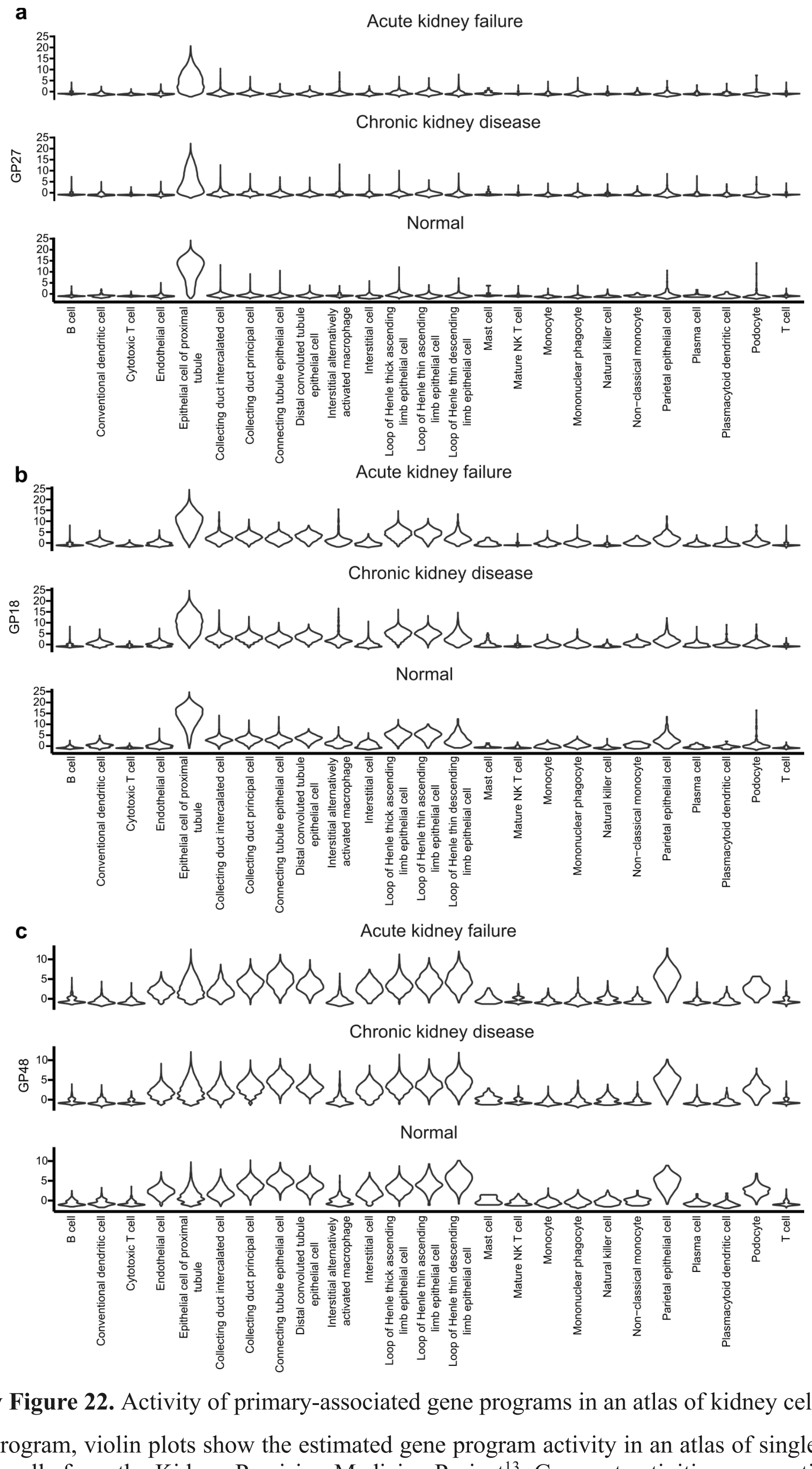

**Supplementary Figure 22.** Activity of primary-associated gene programs in an atlas of kidney cells

For each gene program, violin plots show the estimated gene program activity in an atlas of single-cell expression profiles of kidney cells from the Kidney Precision Medicine Project[13]. Gene-set activities were estimated using the ulm method from decoupleR[14]. Plots are stratified by condition group, including normal healthy donors, donors with acute kidney failure and donors with chronic kidney disease. The gene program shown are: (**a**) GP27, (**b**) GP18 and (**c**) GP48.

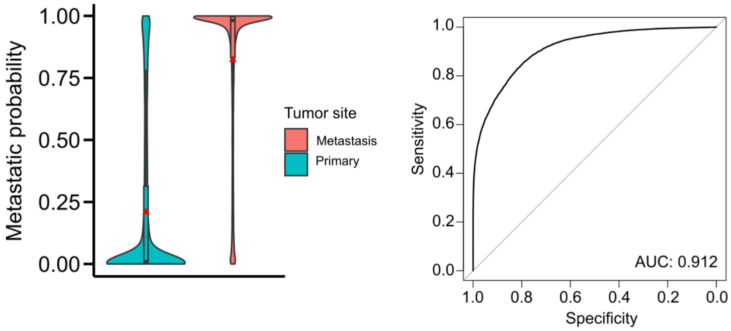

**Supplementary Figure 23.** Internal validation of the metastatic classifier

(**a**) Distribution of metastasis signature scores in primary and metastatic ccRCC cells. Scores represent predicted probabilities from an elastic net logistic regression model evaluated using leave-one-out sample cross-validation (see **Methods**). Violin plots and boxplots summarize the distribution across malignant ccRCC cells; red triangle: mean, center line: median; box limits: upper and lower quartiles; whiskers: 1.5x the interquartile range. (**b**) Receiver operating characteristic (ROC) curve assessing classification performance in distinguishing metastatic from primary tumor cells.

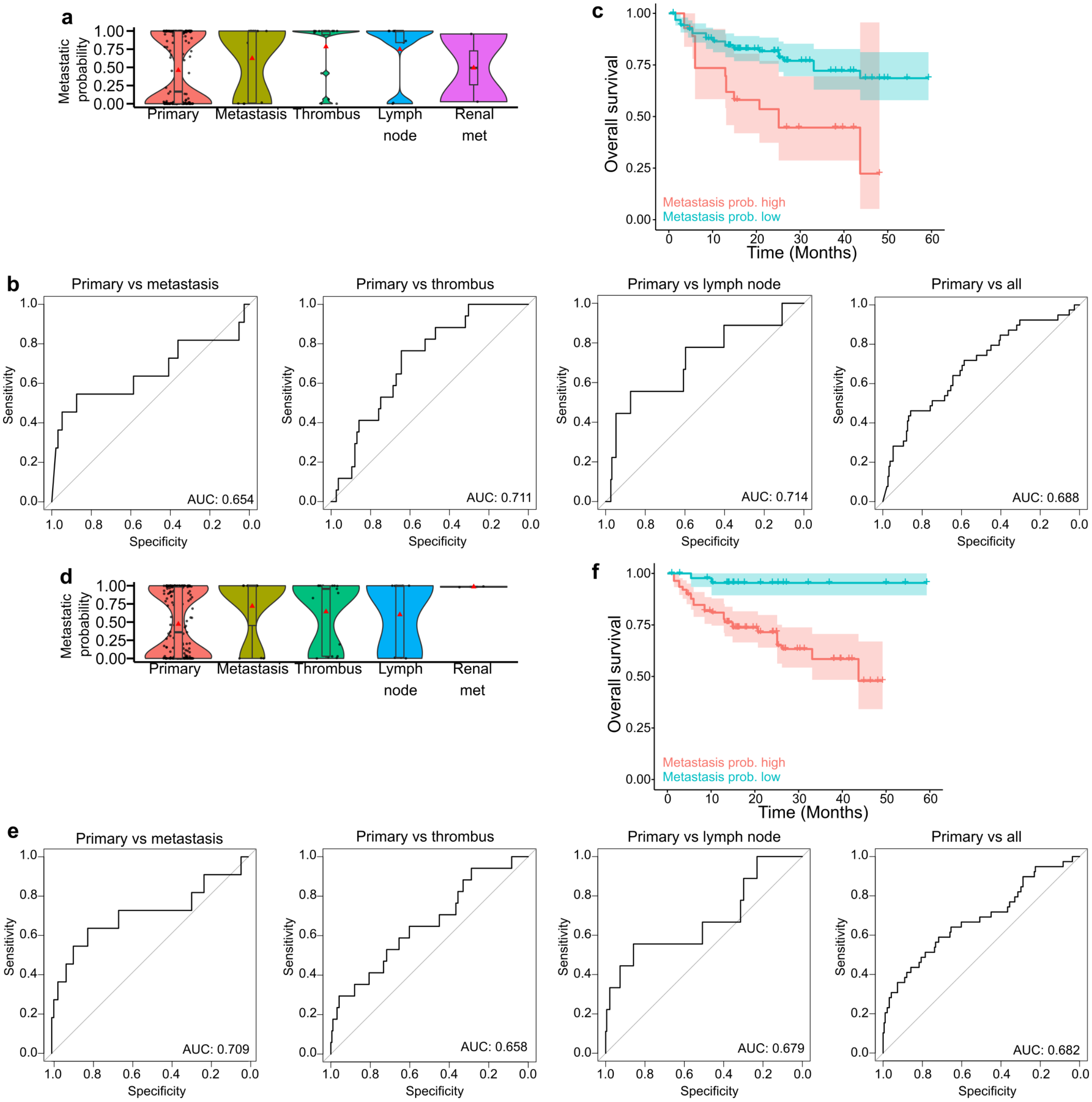

**Supplementary Figure 24.** External validation of the metastatic classifier using data from TRACERx

(**a**-**c**) Validation of the full 111-gene metastatic signature. (**a**) Distribution of metastasis signature scores in ccRCC tumors from the TRACERx[15] cohort. Scores represent predicted probabilities from an elastic net logistic regression model. Violin plots and boxplots summarize the distribution across ccRCC tumors; red triangle: mean, center line: median; box limits: upper and lower quartiles; whiskers: 1.5x the interquartile range. (**b**) Receiver operating characteristic (ROC) curve assessing classification performance in distinguishing: primary versus metastasis, primary versus tumor thrombus, primary versus lymph nodes and primary versus all non-primary tumors. (**c**) Kaplan-Meier survival curves comparing overall survival of ccRCC tumors with high versus low full metastasis signature scores in the TRACERx cohort. Patients were stratified into high and low-score groups based on the optimal cutoff determined during testing (see **Methods**). (**d**-**f**) Validation of the compact 16-gene metastatic signature. Panels follow the same structure as in (a-c) but using the compact metastatic signature.

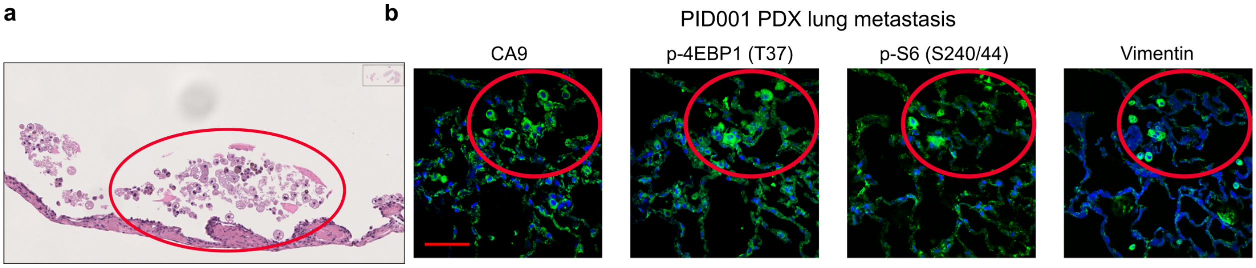

**Supplementary Figure 25.** Distant metastasis of a primary tumor-derived PDX

(**a**) Hematoxylin and eosin staining of lung tissue from the PID001 ccRCC PDX. (**b**) Immunofluorescence staining intensities of CA9, p-4EBP1 (T37), p-S6 (S240/244) and vimentin in metastatic lung nodules of the PID001 ccRCC PDX, a sample with high metastatic probability score. Red circles highlight the regions with CA9-positive metastatic cancer cells on each consecutive slide. Scale bar = 50 µm.

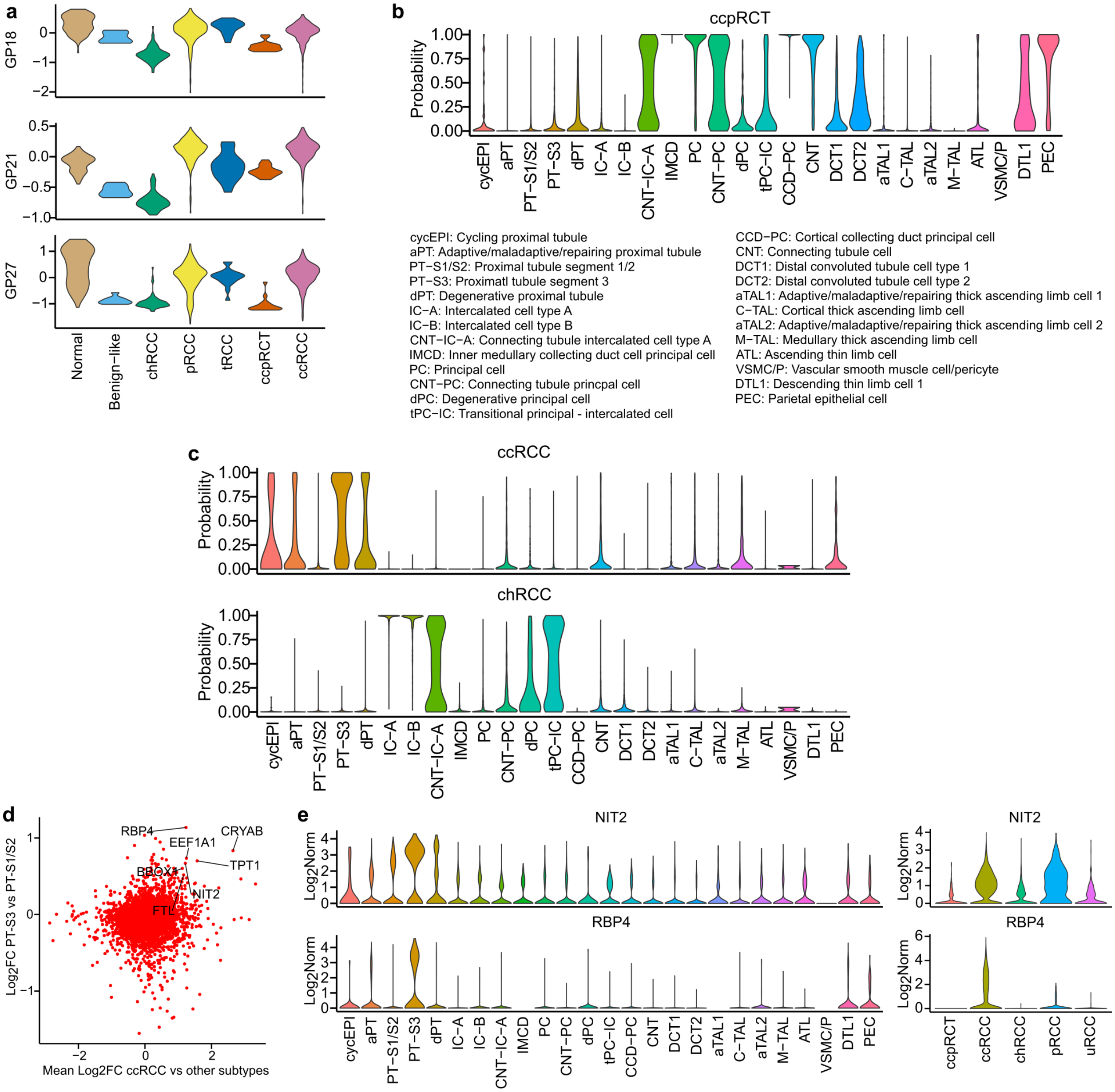

**Supplementary Figure 26.** Cellular origins of RCC subtypes

(**a**) Violin plots showing the distribution of proximal tubule-related gene program activities across RCC subtypes and normal samples from TCGA. (**b**) Violin plots showing predicted probabilities of the ccpRCT subtype for its predicted cell of origin across normal kidney cell types from the Kidney Precision Medicine Project[13] (KPMP) single-cell atlas. To generate these predictions, epithelial single-cells from KPMP and malignant RCC cells from tumors were integrated into a shared latent space using GEDI[16]. Elastic net logistic regression models were then trained on the integrated cell embeddings of malignant RCC cells to classify subtype identity and subsequently applied to the embeddings of normal epithelial cells from KPMP. (**c**) Same as (b) but showing predicted probabilities for ccRCC and chRCC subtypes. (**d**) Scatterplot of $\log_2$ fold-changes highlighting gene markers shared between ccRCC and PT-S3 cells. Differential expression was performed using the scoreMarkers function. For the x-axis, ccRCC cells were compared with other RCC subtypes (excluding pRCC), while for the y-axis PT-S3 cells were compared with PT-S1/S2 cells. (**e**) $\text{Log}_2$ normalized gene expression values of selected marker genes across kidney epithelial cell types from KPMP single cell atlas and malignant RCC cells.

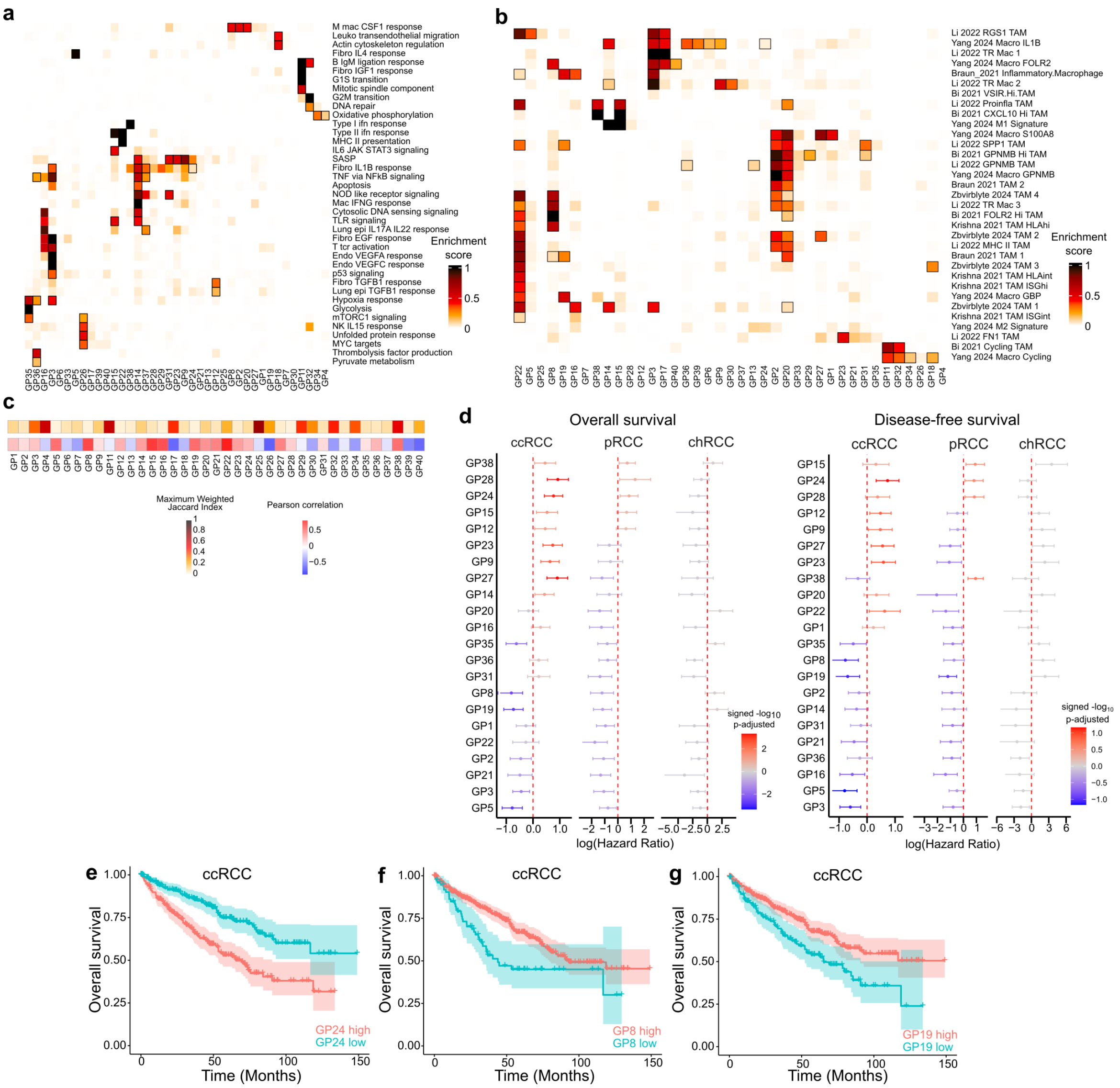

**Supplementary Figure 27.** Macrophage gene program analysis

(**a**) Association between 39 TAM gene programs (columns) and Cytopus immunology knowledge base pathways[17] (rows). Heatmap shows enrichment scores, with 58 significant associations (FDR < 0.05) outlined with a black box. (**b**) Same as (a) but showing association between TAM gene programs (columns) and curated gene sets (rows), summarizing previously described macrophage populations in RCC. The gene sets were curated from the following sources: Li 2022[18], Yang 2024[19], Braun 2021[20], Bi 2021[21]. (**c**) TAM gene programs selected for survival analysis. Heatmap shows Pearson correlation values between GP activity and inferred macrophage cell type proportion in TCGA, as well as comparison between the macrophage and malignant gene programs. Only programs positively correlated with macrophage cell type proportion were retained for survival analysis. (**d**) Overall and disease-free survival analysis of TAM gene programs using TCGA data across the three major RCC subtypes. Plots show log hazard ratios with 95% confidence intervals from a Cox regression model adjusted for estimated macrophage proportion and sex. Color represents signed -log10 FDR values. Thirteen TAM gene programs were significantly associated with patient outcomes (FDR < 0.05) in at least one RCC subtype. (**e-g**) Representative examples of macrophage gene programs associated with survival. Kaplan-Meier survival curves comparing overall survival of ccRCC tumors with high versus low TAM programs: (**e**) TAM GP24, characterized by high expression of *CCL20, G0S2* and *IL6*, markers of inflammatory cytokine-enriched TAMs[22]; (**f**) TAM GP8, representing a *FOLR2*+ TRM-like state; and (**g**) TAM GP19, representing a *CX3CR1*+ TRM-like state. Macrophage gene programs were projected onto bulk-RNA seq profiles from TCGA primary tumors.

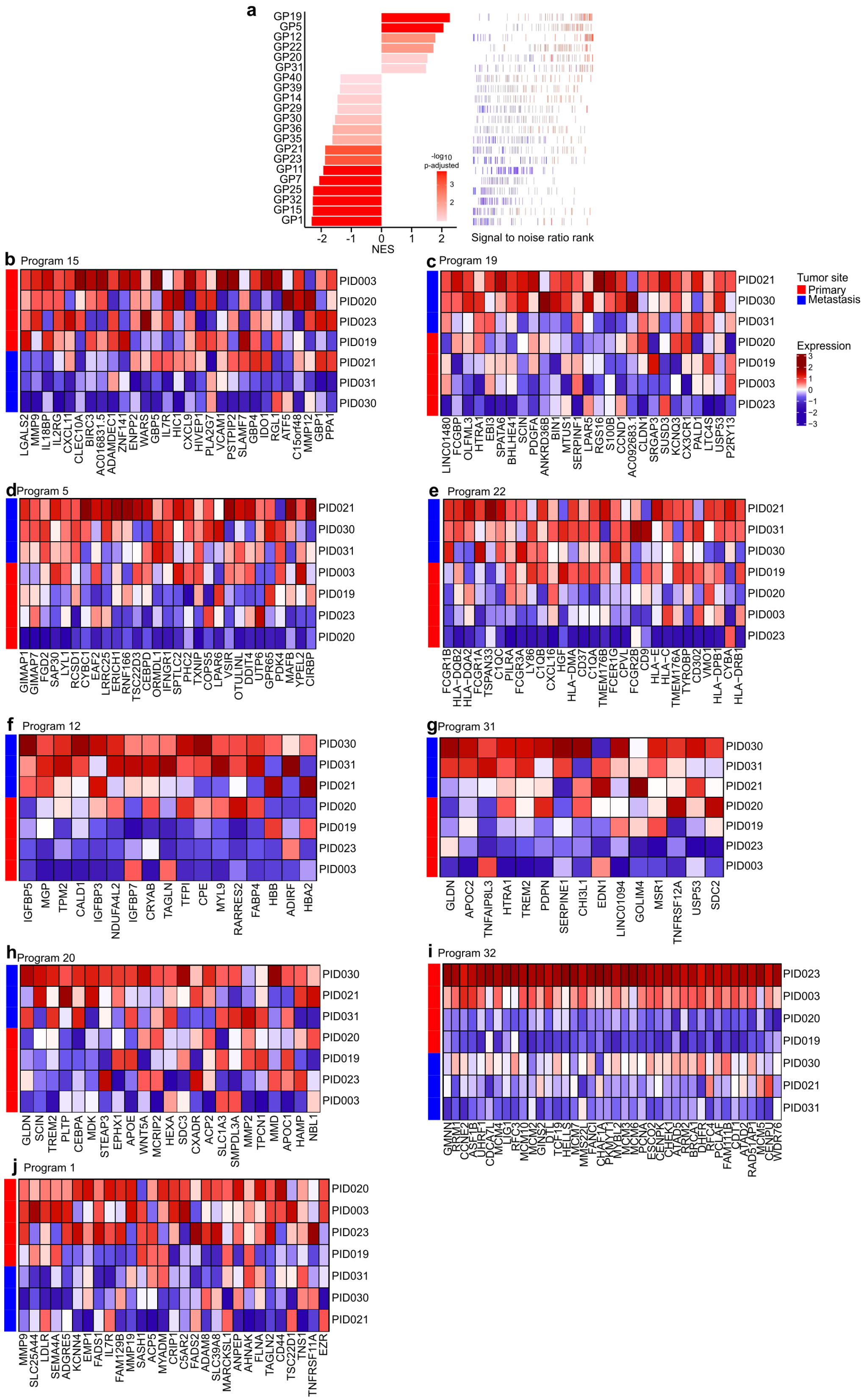

**Supplementary Figure 28.** Macrophage gene programs differentially enriched between metastatic and primary ccRCC (Figure caption continued on the next page).

(**a**) Metastasis-associated macrophage gene expression programs. Left: bar plot showing normalized enrichment scores from GSEA for 21 programs with significant enrichment (FDR < 0.1). Right: signal-to noise-ratio ranking for genes within each program. For each gene program, member genes are shown as bars colored by log2 fold-change from differential expression analysis (**b-j**) Programs differentially enriched between metastatic and primary ccRCC. For each gene program, a heatmap of leading-edge genes is shown, displaying column-wise z-scores of $log_2$-normalized pseudo-bulk RNA-seq expression profiles from macrophages in ccRCC samples. The left annotation bar indicates metastatic status. Programs shown are: (**b**) GP15, (**c**) GP19, (**d**) GP5, (**e**) GP22, (**f**) GP12, (**g**) GP31, (**h**) GP20, (**i**) GP32 and (**j**) GP1. GP15 leading genes overlap with M1 signatures, including *CXCL9* and *CXCL11,* markers of interferon-primed TAMs[22], as well as *LGALS2* which has been suggested to produce a pro-inflammatory phenotype in macrophages[23]. GP22 is an antigen-presentation program enriched for *HLA* and complement-related genes (*C1QC, C1QB*). GP20 is a lipid-associated macrophage program (*APOC1*, *APOE*). GP31 is characterized by the expression of *SPP1*, *CCL2* and *CCL8*, which are involved in tumor-promoting mechanisms[24-26]. GP12 is associated with genes involved in extracellular matrix remodelling.

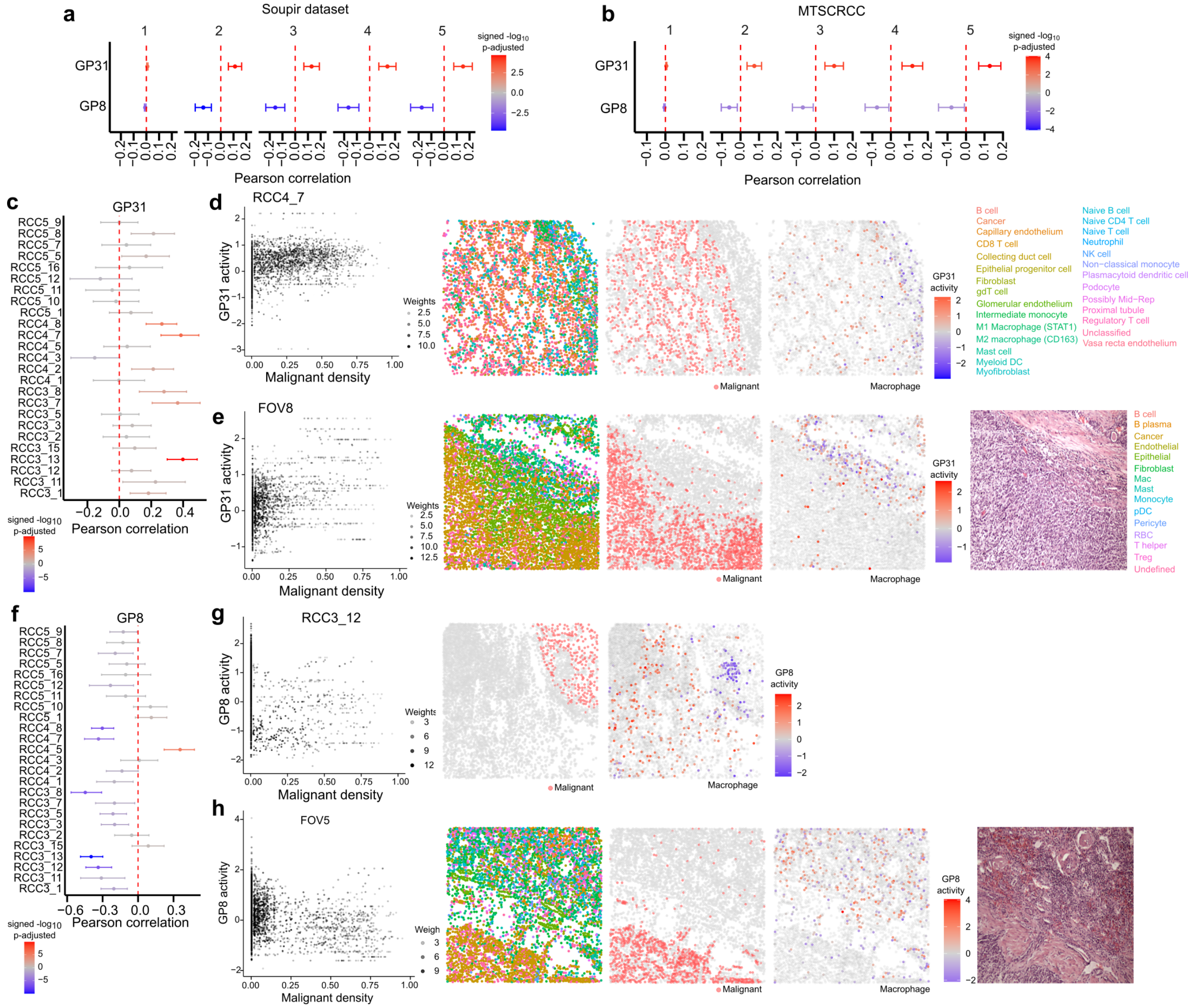

**Supplementary Figure 29.** Spatial analysis of macrophage gene programs

(**a**) Meta-analysis of weighted correlation between local malignant cell density and activities of TAM GP31 and TAM GP8 in a single-cell spatial transcriptomic dataset[11] of ccRCC tumors. Pearson correlation coefficients and 95% confidence intervals were derived via meta-analysis across FOVs. Color represents signed -log$_{10}$ FDR values. Each panel displays a different neighborhood-step *k*. (**b**) Same as in (a), but performing weighted correlation analysis using a single-cell resolution spatial transcriptomic map of an MTSCRCC tumor. (**c**) Weighted correlation analysis between malignant density and TAM GP31 activity across individual FOVs in a ccRCC spatial dataset[11]. (**d**) Example of positive association between malignant density and TAM GP31 activity. Left: scatterplot comparing TAM GP31 activity and malignant density; dot intensity reflects weights representing local neighborhood support for both signals. Middle left: spatial coordinates of cell centroids colored by cell type annotation from the original authors[11]. Middle right: spatial coordinates indicating malignant cells. Right: spatial coordinates showing TAM GP31 activity in macrophages; non-macrophage cells are transparent. (**e**) Example of positive association between malignant density and TAM GP31 activity in a MTSCRCC tumor. Panel format is the same as (d), except that right panel shows hematoxylin and eosin staining. (**f**) Weighted correlation analysis between malignant density and TAM GP8 activity across individual FOVs in a ccRCC spatial dataset[11]. (**g**) Example of negative association between malignant density and TAM GP8 activity in a ccRCC tumor. Panel format is the same as (d), except that the middle-left panel with cell type labels can be found in Fig.6l. (**h**) Example of negative association between malignant density and TAM GP8 activity in a MTSCRCC tumor. Panel format is the same as (e).

**Supplementary Table 1**. List of source data files.

The Zenodo records are available at:

https://zenodo.org/doi/10.5281/zenodo.20058906,

https://zenodo.org/doi/10.5281/zenodo.20086236

| |
|---|
| **File name:** gedi_model_malignant.rds (related to Figure 1)<br><br>**Description:** GEDI model for malignant RCC cells.<br><br>**Download URL:** https://zenodo.org/records/20058907/files/gedi_model_malignant.rds?download=1 |
| **File name:** gedi_model_tumor.rds (related to Figure 1)<br><br>**Description:** GEDI model for integration of tumor cells.<br><br>**Download URL:** https://zenodo.org/records/20058907/files/gedi_model_tumor.rds?download=1 |
| **File name:** numbat_call_per_clone.rds (related to Figure 1)<br><br>**Description:** Dataframe with inferred CNV results from Numbat per clone.<br><br>**Download URL:** https://zenodo.org/records/20058907/files/numbat_call_per_clone.rds?download=1 |
| **File name:** lis_tcga.rds (related to Figure 2).<br><br>**Description:** List with objects for the TCGA data. List contains: 'meta': metadata, 'rc': raw counts of the expression data, 'norm_data': normalized expression data.<br><br>**Download URL:** https://zenodo.org/records/20058907/files/lis_tcga.rds?download=1 |
| **File name:** gedi_model_rcc_tcga.rds (related to Figure 2 and 3).<br><br>**Description:** GEDI model for integration of RCC cells with TCGA RCC samples.<br><br>**Download URL:** https://zenodo.org/records/20058907/files/gedi_model_rcc_tcga.rds?download=1 |
| **File name:** scConvexNMF_model_malignant.rds (related to Figure 4).<br><br>**Description:** scConvexNMF model for malignant gene program discovery using malignant cells from the RCC dataset.<br><br>**Download URL:** https://zenodo.org/records/20086237/files/scConvexNMF_model_malignant.rds?download=1<br>https://zenodo.org/record/8222698/files/COVID19_gedi_model_cohort2.rds?download=1 |
| **File name:** scConvexNMF_rescoringModel_malignant.rds (related to Figure 4).<br><br>**Description:** scConvexNMF rescoring model of malignant gene programs using malignant cells from the RCC dataset.<br><br>**Download URL:** https://zenodo.org/records/20086237/files/scConvexNMF_rescoringModel_malignant.rds?download=1<br>https://zenodo.org/record/8222698/files/COVID19_list_DE.rds?download=1 |
| **File name:** lis_spatial_Soupir.rds (related to Figure 4).<br><br>**Description:** List with objects for spatial analysis of the Soupir dataset. List contains: 'meta': metadata, 'df_coords': Spatial coordinates, 'res_malignant': Colocalization results for the malignant cells, 'res_macrophage': Colocalization results for the macrophage cells.<br><br>**Download URL:** https://zenodo.org/records/20058907/files/lis_spatial_Soupir.rds?download=1 |
| **File name:** scConvex_rescoringModel_Soupir_malignant.rds (related to Figure 4).<br><br>**Description:** scConvexNMF rescoring model of malignant gene programs using malignant cells from the Soupir dataset.<br><br>**Download URL:** https://zenodo.org/records/20058907/files/scConvex_rescoringModel_Soupir_malignant.rds?download=1 |
| **File name:** scConvex_rescoringModel_TCGA_malignant.rds (related to Figure 5). |

| |
|---|
| **Description:** scConvexNMF rescoring model of malignant gene programs using bulk RNA-seq samples from the TCGA dataset.<br><br>**Download URL:** https://zenodo.org/records/20058907/files/scConvex_rescoringModel_TCGA_malignant.rds?download=1 |
| **File name:** lis_tcga_stats.rds (related to Figure 5).<br><br>**Description:** List with objects for the survival analysis in TCGA data. List contains: 'malignant_os': Overall survival results for the malignant RCC cells, 'malignant_dfs': Disease-free survival results for the malignant RCC cells, 'mac_os': Overall survival results for the macrophage cells, 'mac_dfs': Disease-free survival results for the macrophage cells.<br><br>**Download URL:** https://zenodo.org/records/20058907/files/lis_tcga_stats.rds?download=1 |
| **File name:** DESeq2_dds_malignant.rds (related to Figure 6).<br><br>**Description:** DESeq2 object for pseudo-bulk malignant ccRCC samples.<br><br>**Download URL:** https://zenodo.org/records/20086237/files/DESeq2_dds_malignant.rds?download=1<br>https://zenodo.org/records/11164777/files/CFDE_miloDE.rds?download=1 |
| **File name:** lis_gsea.rds (related to Figure 6).<br><br>**Description:** List with objects of GSEA results. List contains: 'malignant': list with GSEA results for malignant gene programs, 'mac': list with GSEA results for macrophage gene programs.<br><br>**Download URL:** https://zenodo.org/records/20086237/files/lis_gsea.rds?download=1 |
| **File name:** lis_spatial_PID037.rds (related to Figure 6)<br><br>**Description:** List with spatial results for the PID037 sample. 'res_malignant': Colocalization results for the malignant cells, 'res_macrophage': Colocalization results for the macrophage cells.<br><br>**Download URL:** https://zenodo.org/records/20086237/files/lis_spatial_PID037.rds?download=1 |
| **File name:** scConvex_rescoringModel_spatial_PID037_malignant.rds (related to Figure 6).<br><br>**Description:** scConvexNMF rescoring model of malignant gene programs using malignant cells from PID037.<br><br>**Download URL:**<br>https://zenodo.org/records/20086237/files/scConvex_rescoringModel_spatial_PID037_malignant.rds?download=1 |
| **File name:** RCC_metastasis_score.rds (related to Figure 7).<br><br>**Description:** Dataframe with metastatic scores for ccRCC malignant cells.<br><br>**Download URL:** https://zenodo.org/records/20086237/files/RCC_metastasis_score.rds?download=1&preview=1 |
